# Outcomes of planned caesarean birth compared with planned or actual vaginal birth: an update and expansion of the NICE Caesarean Birth Guideline systematic review NG192

**DOI:** 10.64898/2026.05.28.26354321

**Authors:** Mairead Black, Clare Robertson, Moira Cruickshank, Aniebiet Ekong, Paul Manson, Ogechi Kemakolam, Olivia Steel, Clare Richards, Pavithra Harshani, Abi Merriel, Declan Devane, Siladitya Bhattacharya, Denitza Williams, Miriam Brazzelli

## Abstract

**Background:** Planned caesarean birth (CB) is an increasingly utilised intervention, observed in almost 1 in 6 first-time mothers giving birth in the UK in 2023-24. Outcomes of planned (or actual) CB have been compared with planned (or actual) vaginal birth (VB) in a UK national guideline, but the scope of the comparison does not fully reflect the range of outcomes of interest to stakeholders. This review provides a comprehensive synthesis of outcomes of planned or actual CB with planned or actual VB to shape information resources which support informed birth planning.

**Methods:** The UK NICE Caesarean Birth Guideline NG192 evidence review of outcomes associated with planned CB (or actual CB where no planned CB data was available) was updated and expanded to incorporate additional outcomes prioritised by stakeholders.

**Results:** A total of 33 new study reports were combined with 32 reports previously included in NG192. All new reports were observational cohort studies or systematic reviews at low risk of bias. Only 3 studies reported outcomes of planned CB compared with planned VB (regardless of actual mode of birth), whereas all remaining studies reported actual VB outcomes.

Planned CB was followed by more maternal infection (wound infection, mastitis, endometritis and urinary tract), venous thrombosis and lower neonatal unit admission rates than a planned VB. In the long-term, CB was linked to one or more sexual problems (insufficient lubrication and dyspareunia) being more common, future pregnancy being less common, and infertility being more frequent than after VB. For offspring, infant urinary tract infection after any CB, gastrointestinal tract infections and autism after planned CB were more common compared with VB. New findings highlight conflicting reports on childhood asthma and type 1 diabetes risk after planned CB, suggesting that prior positive associations may be explained by confounding.

Existing evidence in NG192 suggests that cardiac arrest, maternal death and hysterectomy are more common after planned CB, but arise from studies at high risk of bias. NG192 also reports that placenta accreta and uterine rupture in a future pregnancy are more common after any CB. No new evidence was identified on these outcomes.

**Conclusion:** This review provides stakeholder-relevant information to populate decision-support materials on outcomes of planned (and actual) CB compared with planned (and actual) VB. The existing evidence base lacks data on long-term outcomes of planned (rather than actual) VB.

## Introduction

Caesarean birth (CB) is increasingly utilised for relative indications or maternal choice,^1–3^ with overall CB rates rising from 4% in England in 1980 to 42% in 2023-2024.^4^ In 2023-24, first births to UK mothers comprised: 13% planned CB; 18% vaginal births (VB) assisted by forceps or ventouse; 31% unplanned CB; and 36% unassisted VB.^4, 5^ This change reflects a shift in attitudes, governance, policy, and the legal stance that increasingly support the mode of birth (MOB) choice.^6–9^ With growing acceptance of CB as a reasonable option, the need for evidence-based information on its relative risks and benefits has increased. In the UK, CB for maternal choice was first supported by the National Institute for Health and Care Excellence (NICE) CB guidance in 2011, which recommended that women who request CB be given information and counselling on birth options, and that an informed CB request be honoured.^10^

The NICE CB guideline (NG192) includes an evidence review assessing the benefits and risks of planned CB compared with planned VB, although the scope of the review is not exhaustive. Maternal risks of planned CB include increased infection, maternal cardiac arrest and death, and future life-threatening pregnancy complications, including scar rupture and placenta accreta. CB has also been linked to an increased risk of asthma in children. Planned VB carries a substantial risk of perineal tears and a higher risk of faecal and urinary incontinence compared with CB. With only one third of planned VBs in a first pregnancy ending in unassisted VB, many women have unplanned operative birth, which is linked to less favourable birth experiences and more post-traumatic stress-type symptoms, though these outcomes were not addressed by NG192.^11, 12^

The objective of this study was to update and expand the NG192 evidence review of the long-and short-term maternal and neonatal outcomes when comparing planned CB and planned VB to inform birth planning. The findings will be used to shape decision support material for pregnant women to help them plan their birth in routine NHS care.^13^

## Methods

An update and expansion of the evidence review of short- and long-term outcomes of planned CB compared with planned VB, which informed the NICE NG192, was undertaken.^7^ The review was conducted according to current methodological standards and reported in adherence to the Preferred Reporting Items for Systematic Reviews and Meta-Analyses (PRISMA) 2020 statement for reporting systematic reviews and meta-analyses.^14–16^ This review is part of a wider mixed methods study aimed at developing a decision aid to support planned MOB conversations in routine antenatal care in the UK NHS and comparable settings (the Plan-A study, researchregistry8238). The study protocol has been made available at: https://www.abdn.ac.uk/acwhr/research/plan-a-193.php#panel207 since 2022.

### Search methods for the identification of studies

The literature search for the NICE guideline on caesarean birth (NG192) was largely replicated for this review. The CRD Health Technology Assessment database, which was searched for NG192, has not been updated since 2015, so it was replaced by the International HTA Database. The other databases searched were Ovid MEDLINE, Ovid Embase, the Cochrane Database of Systematic Reviews, and CENTRAL. The ScHARR Low and Middle Income Countries filter was applied to MEDLINE and Embase to limit the results to high-income countries.^17^ The initial search covered the period from the final search for NG192 (July 2019) through December 2022; it was updated in May 2025. All results were exported to EndNote for recording and deduplication. The searches are available in supplementary file S1.

### Inclusion and exclusion criteria

The key eligibility criteria are based on the criteria of NG192 and are outlined below.

#### Types of studies

Eligible study designs included randomised and non-randomised comparative studies, cohort studies, case-control studies, cross-sectional studies and population-based registry studies. Systematic reviews, with or without meta-analyses, of randomised controlled trials or observational studies were also eligible for inclusion. Studies conducted in low/middle-income countries and studies that did not report data that had been adjusted for any confounding factors were excluded.

#### Types of population (participants)

Primiparous and multiparous pregnant women with singleton pregnancies were included without age restriction. For short-term outcomes, women with pregnancies at lower obstetric/medical risk (no absolute medical/obstetric indication for CB) were included if data were analysed according to planned MOB. In keeping with the methods outlined in the NG192 review, for long-term outcomes, studies of women with any indication for CB were included (see exposure details below). This is due to the expected sparsity of long-term data reported by planned MOB.

In keeping with the NG192 review, studies including only pregnant women with relative or absolute clinical indications for CB, such as breech presentations, multi-foetal pregnancies, preterm births, babies who were small for gestational age, placenta praevia, and maternal infections, were excluded.

#### Types of interventions

For studies reporting short-term outcomes: Planned CB

For studies reporting long-term outcomes: Any CB (planned or unplanned). Studies that reported data for planned CB were prioritised over those which reported data for planned and unplanned CB as one group.

#### Types of comparators

For studies reporting short-term outcomes: Planned VB (including spontaneous and assisted VB, and unplanned CB)

For studies reporting long-term outcomes: Planned or actual VB. Studies that reported data for planned VB were prioritised over those which reported data for actual VB as one group. Studies were included if they reported data on all VBs, while those that reported only spontaneous VBs were included only if no studies reported planned or all VBs.

#### Types of outcomes

Outcomes eligible for inclusion from the original search were those considered in the NG192 evidence review covering maternal short-term (up to 6 weeks), maternal long-term (beyond 6 weeks), infant short-term (up to 7 days of age) and children long-term (from 7 days to 18 years of age). Additionally, outcomes identified as important to stakeholders involved in MOB planning were included in an updated search, as determined through a Delphi process comprising a 2-round survey and a consensus meeting. Delphi participants included healthcare professionals, women who were currently pregnant, and women who had previously given birth, either by VB or CB. The outcomes reported in the main results section include those previously reported in the NG192 as well as those prioritised through the Delphi process. Clinical outcomes identified during the review that did not meet NG192 or Delphi inclusion criteria but were judged by the authors to be of material clinical significance are reported for completeness as supplementary material.

### Short-term outcomes

*Eligible maternal short-term outcomes included:* haemorrhage; hysterectomy; bladder injury; bowel injury; ureteric injury; thromboembolic disease; pain during/after birth; infection (any postpartum infection); blood transfusion; infection; intensive care admission; maternal death; length of hospital stay.

*Eligible infant short-term outcomes included:* perinatal mortality; neonatal unit admission; neonatal respiratory morbidity; neonatal infectious morbidity; hypoxic ischaemic encephalopathy; intracranial haemorrhage; nerve injury.

### Long-term outcomes

*Eligible maternal long-term outcomes included:* satisfaction with birth experience; quality of life; post-traumatic stress disorder due to childbirth; urinary incontinence 1 year or more after birth; faecal incontinence 1 year or more after birth; pelvic organ prolapse; postnatal depression; sexual dysfunction. Future pregnancy complications: miscarriage, placenta accreta spectrum, uterine rupture, stillbirth.

*Eligible child long-term outcomes included*: neurodevelopmental disorder; cognition/communication/psychomotor disorder; cerebral palsy; childhood obesity; asthma; type 1 diabetes; autism; infant/child mortality.

### Additional outcomes

Outcomes identified as important during the Delphi consensus process included *maternal outcomes* (actual MOB; placental abruption; anaesthetic type; anaesthetic complications; postpartum recovery; postpartum anxiety), *outcome related to future pregnancies* (preterm birth; fertility; MOB; caesarean scar pregnancy) and *infant outcomes* (meconium aspiration syndrome; skin-to-skin contact; bruising).

#### Study selection

Three reviewers (OK, OS and CRi) screened the titles and abstracts from the initial search results to identify potentially eligible articles. A second reviewer (MBl, MBr) independently screened a random 20% sample of all citations to assess consistency. For the updated search, titles and abstracts were double screened by two of three reviewers (MBl, AE, PH) within Covidence™ software.

#### Appraisal of methodological quality, Data extraction and Statistical analysis

Appraisal of methodological quality was performed by single reviewers (OK, OS, CRi) using the Newcastle-Ottawa scale for primary studies^18^ and the ROBIS tool for systematic reviews.^19^

Single data extraction was conducted by four reviewers (OK, OS, CR, MC) for outcomes relevant to planned or actual CB and VB, depending on the eligibility criteria for short- and long-term outcomes. Adjusted effect measure data from the newly included studies and the study data reported in NG192 were collated for each outcome. Where similar outcomes data were identified from either primary studies or systematic reviews, meta-analyses were conducted using the inverse-variance method with fixed effects models. Random effects models were used when heterogeneity was identified. For the meta-analyses, CB was defined as the target MOB and VB as the reference mode. Analyses were performed in RevMan Web using adjusted effect measures reported by the study authors (odds ratio, hazard ratio, or relative risk). Standard errors were calculated from the reported 95% CI data using the RevMan Web calculator. Where meta-analysis was not possible due to an insufficient number of comparable studies reporting equivalent measures of effect, findings were summarised descriptively. Absolute effect estimates reported in NG192 were extracted for the purposes of this review. Absolute effects for newly included outcomes were calculated following the recommendations of the Cochrane Handbook for Systematic Reviews of Interventions.^20^

## Results

The literature searches identified 9775 citations. After screening titles and abstracts, 242 full-text records were assessed for eligibility; 33 reports representing 32 studies met the inclusion criteria. The previous NICE evidence review included a total of 32 studies^7^ therefore, the updated review incorporates evidence from 64 studies reported across 65 reports. The study selection process is illustrated in Figure 1. Of those which met inclusion criteria, two studies were excluded following full-text assessment. The study by Gonzalez-Timoneda et al. (2025)^21^ was excluded because the reported subgroup numerators did not align with the total group numerators, and the direction of effect estimates was inconsistent with the proportions presented, preventing reliable interpretation of the results. The study by Yang et al. (2022)^22^ was excluded due to concerns about the reliability of the results and the lack of a response to author correspondence seeking clarification.

**Figure 1.**
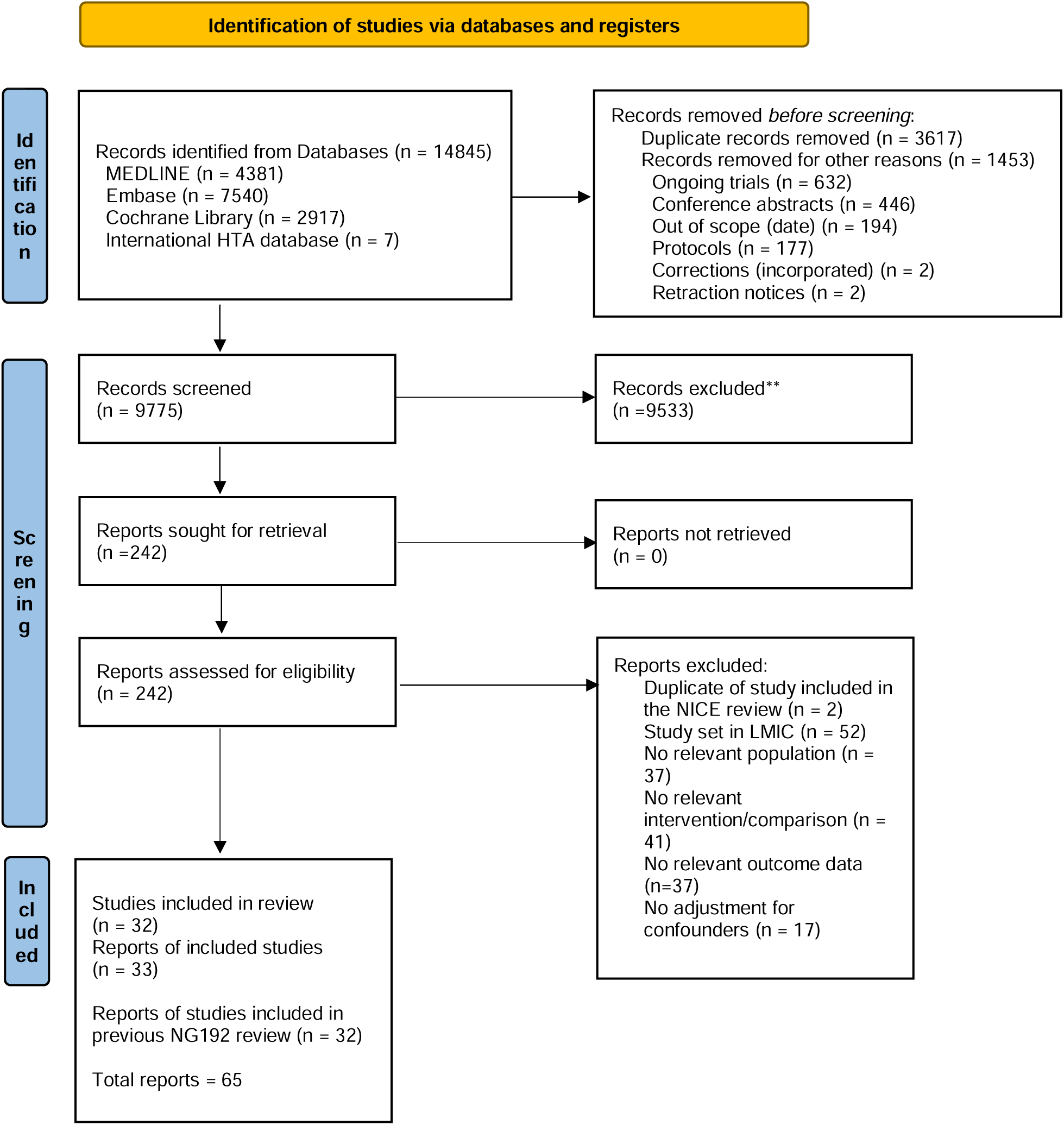
PRISMA 2020 flow diagram for new systematic reviews which included searches of databases and registers only^1^.

The characteristics of the 32 studies included in the NICE evidence review are detailed in the published NICE guideline NG192.^7^ Of the 32 additional studies identified in this update, seven were systematic reviews,^23–29^ with the remainder being cohort studies. Fourteen studies were conducted in Europe, including two in the UK,^30, 31^ four in Sweden,^32–35^ three in Denmark,^36–38^ two in Switzerland,^39, 40^ two in Norway,^41, 42^ and one in Finland.^43^ Two studies were conducted across Europe and Oceania: one in the UK and New Zealand^44^ and one in Denmark, Scotland, England, and Australia.^45^ Two additional studies were conducted in Canada,^46, 47^ three in the USA,^48–50^ two in Japan,^51, 52^ one in Australia,^53^ and one in Taiwan.^54^ The seven systematic reviews included studies conducted across multiple countries and continents, including some in low- and middle-income countries (LMICs). However, many studies within these systematic reviews (at least 70%) were conducted in high-income countries. From the 32 additional studies identified in this review, only two (three reports) directly compared planned caesarean birth with planned vaginal birth,^34, 47, 55^ with remaining studies either comparing planned caesarean birth with any vaginal birth, or any caesarean birth with any vaginal birth.

In this update, short-term outcomes data were provided by three reports from two studies: Dahlquist et al (2022),^34^ Guo et al (2021),^47^ and Guo et al (2024).^55^ The publications by Guo et al (2021)^47^ and Guo et al (2024)^55^ were considered to represent reports from the same underlying study; for the purposes of summarising birth numbers, Guo et al (2021) was treated as the primary study report. Across the short-term outcome studies, data were available for a total of 24,682 planned CBs and 1,111,854 planned VBs. In the study by Dahlquist et al (2022),^34^ of the 691,471 women who planned a VB, 30,623 (4.4%) subsequently had a CB. Similarly, in the Guo et al (2021) study,^47^ 44,706 of 420,383 women (10.6%) who planned a VB subsequently had a CB.

Long-term outcomes data were provided by 23 primary studies (reporting 1,064,187 CB and 4,694,242 VBs)^30–33, 35–46, 48–54^ and seven systematic reviews^23–29^ (including 53 studies; the number of CBs and VBs was not consistently reported).

Main characteristics of these studies are reported in Supplementary file S2.

### Appraisal of Methodological Quality

The study level summary of the quality assessment of the included studies is presented in Supplementary File S3. All primary studies were judged to be of good methodological quality and all systematic reviews were considered to have low risk of bias.

### Clinical outcomes

Outcomes that were reported in NG192, along with those prioritised for inclusion through the Delphi process, are presented below.^13^ Additional outcomes are presented in Supplementary file S4. Short- and long-term maternal and offspring outcomes are presented separately. Outcomes from studies reported in Appendix N of NG192 but not included in the main text of NG192, as “*their reported effect estimates did not substantially alter the overall estimate of included systematic reviews assessing the same outcome*”, are considered in this review alongside data from newly identified studies. Tables 1 and 2, respectively, present summaries of short-term and long-term outcomes reported in NG192 and those identified in this update.

**Table 1.**
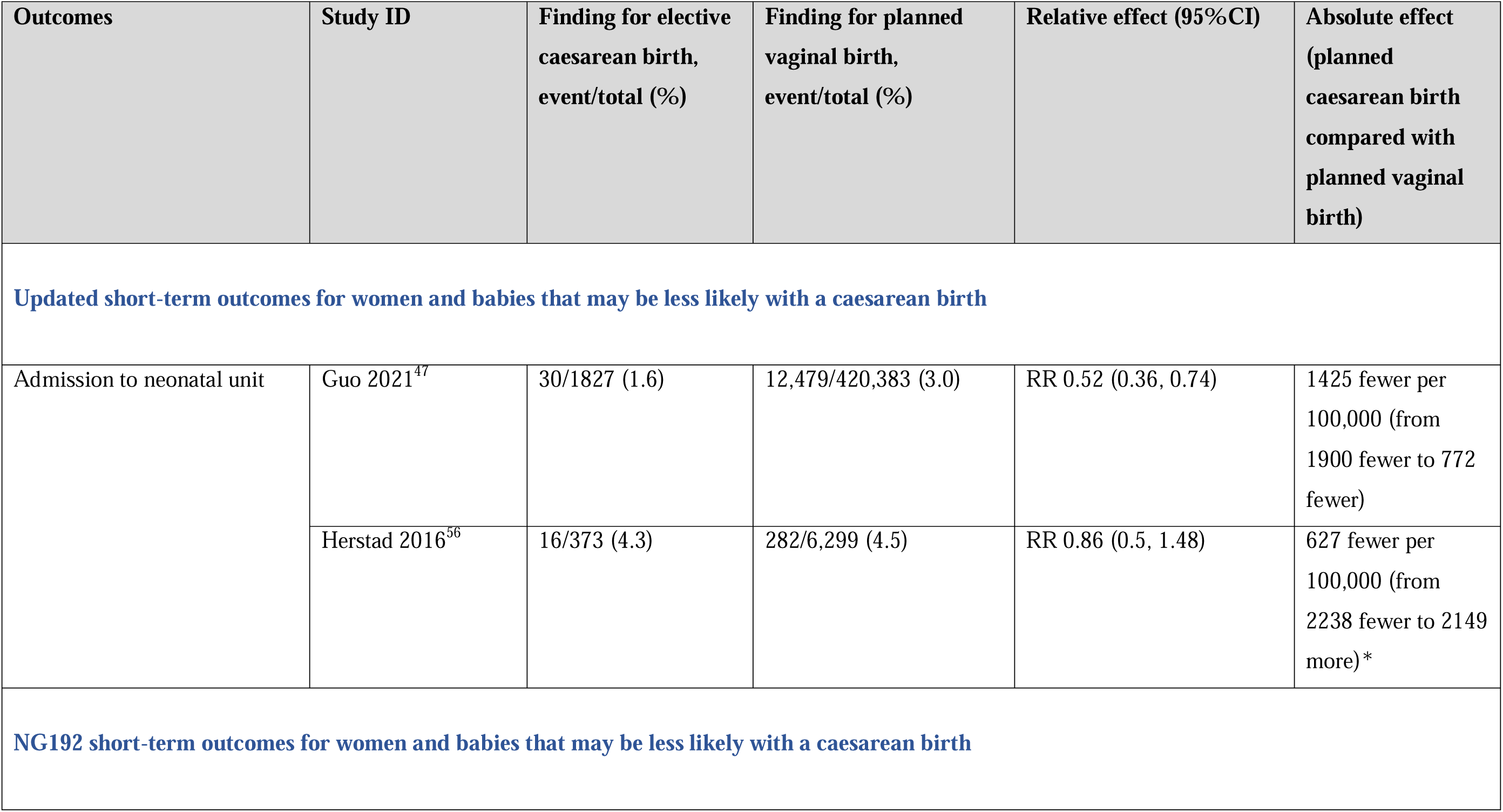
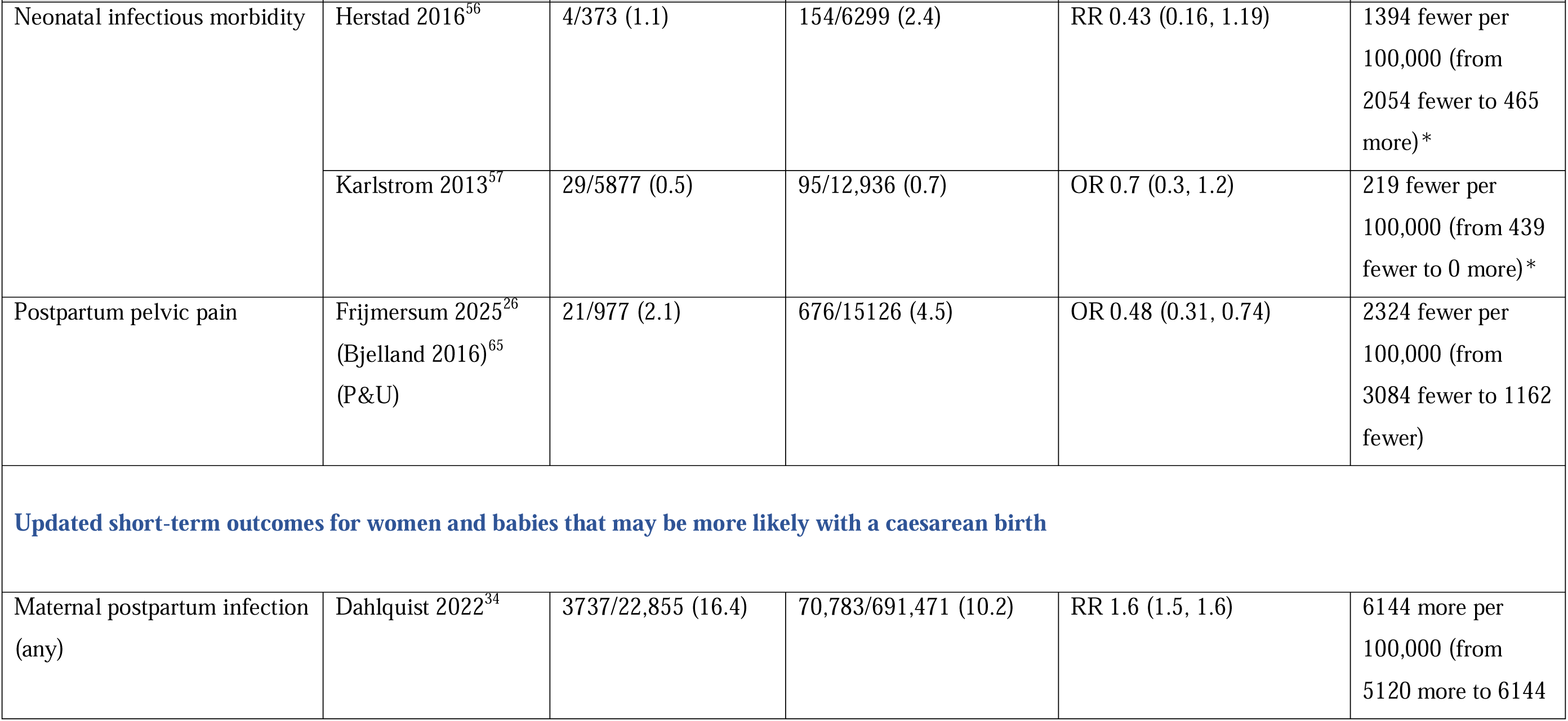
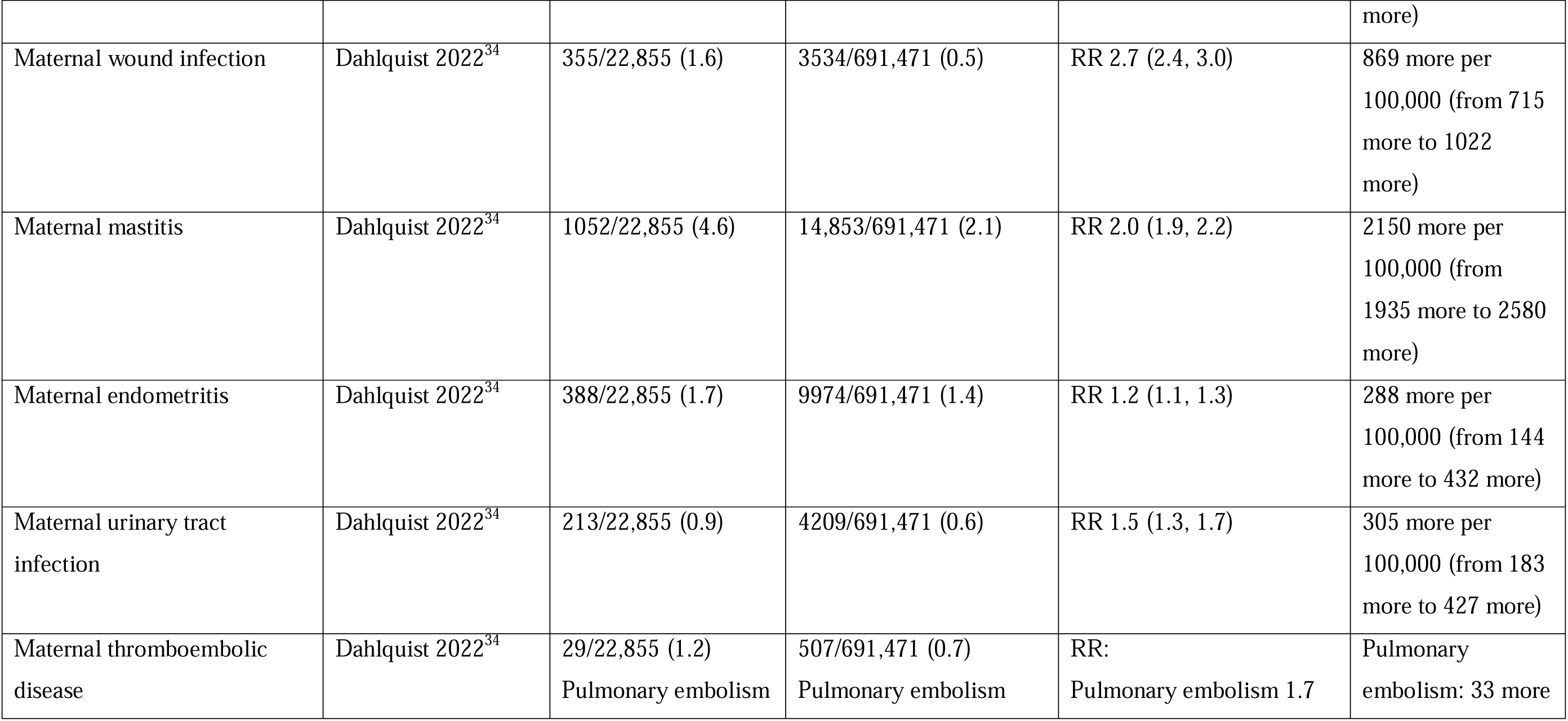
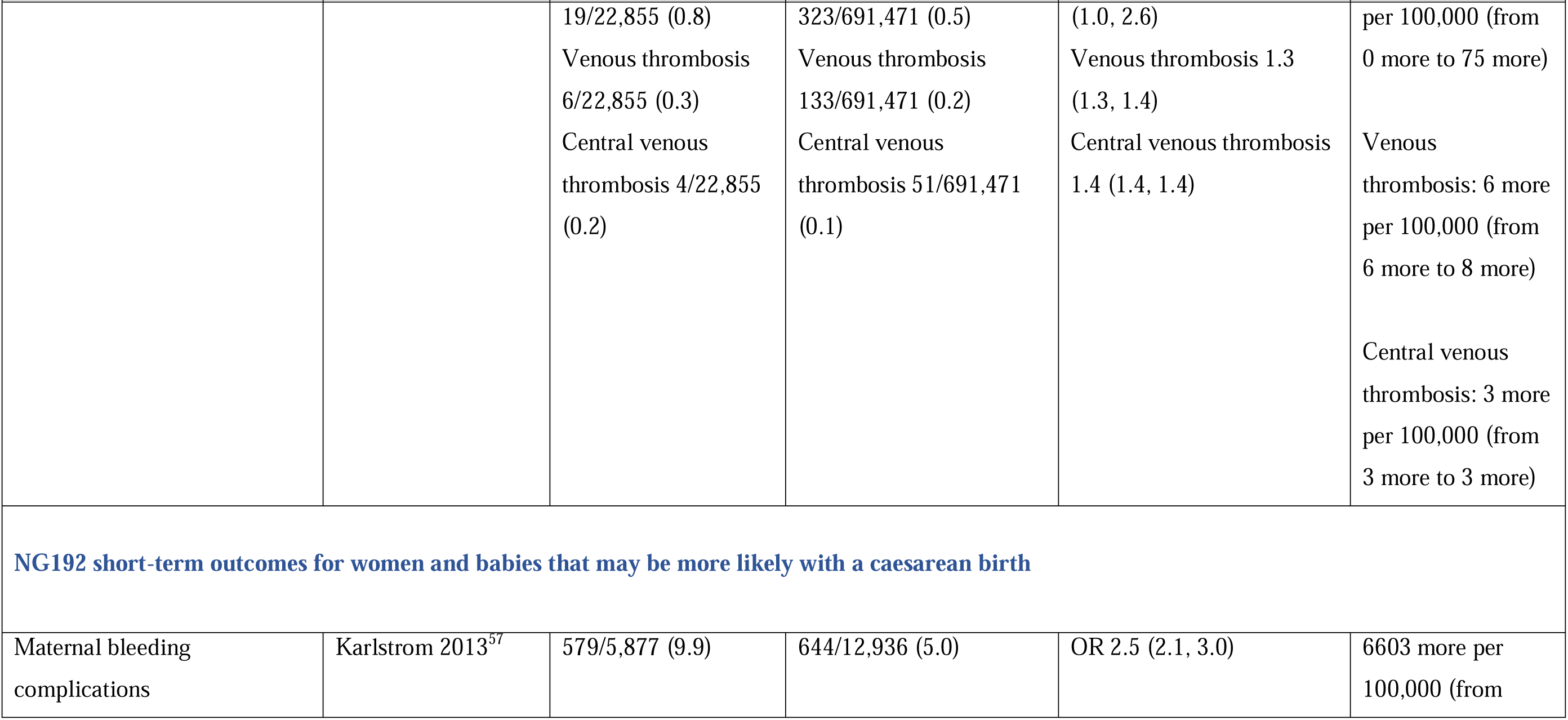
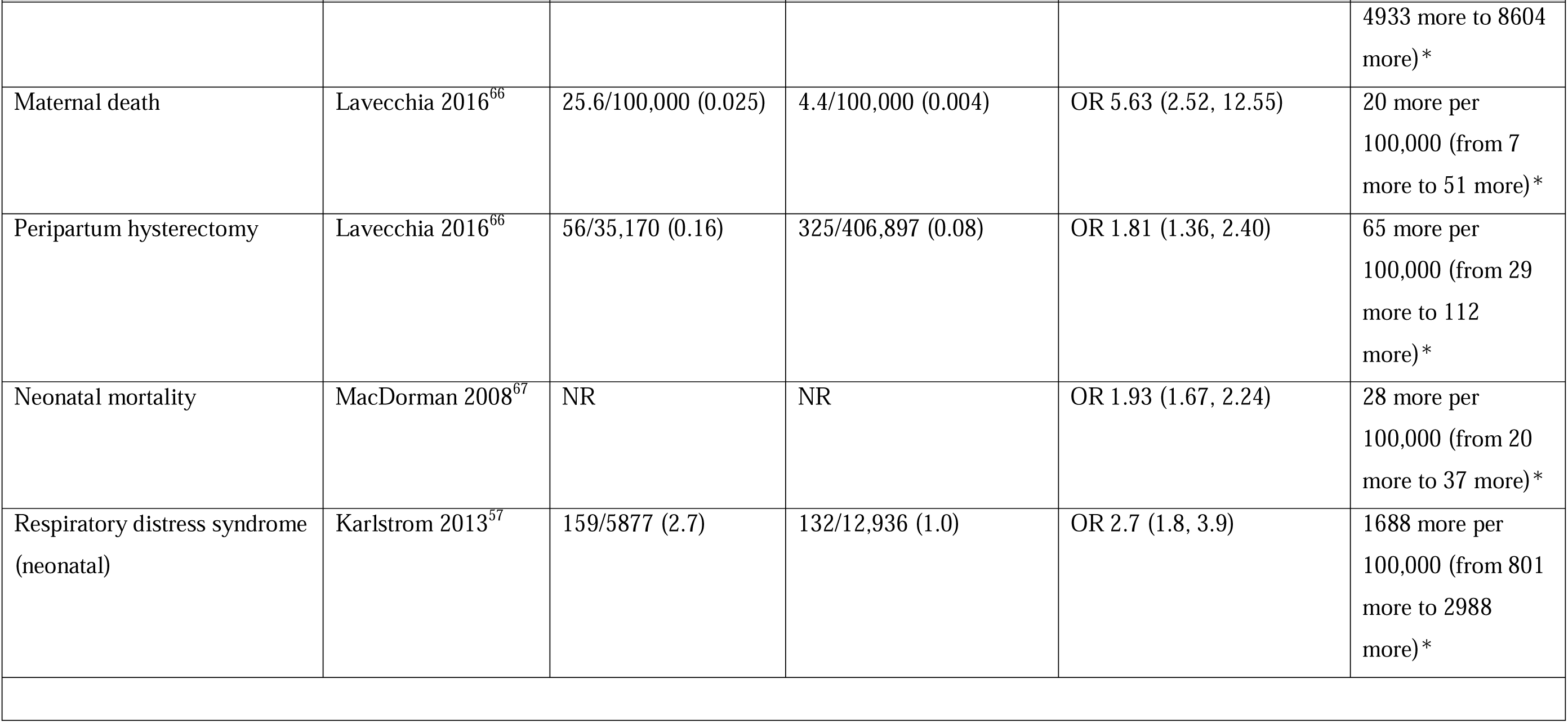
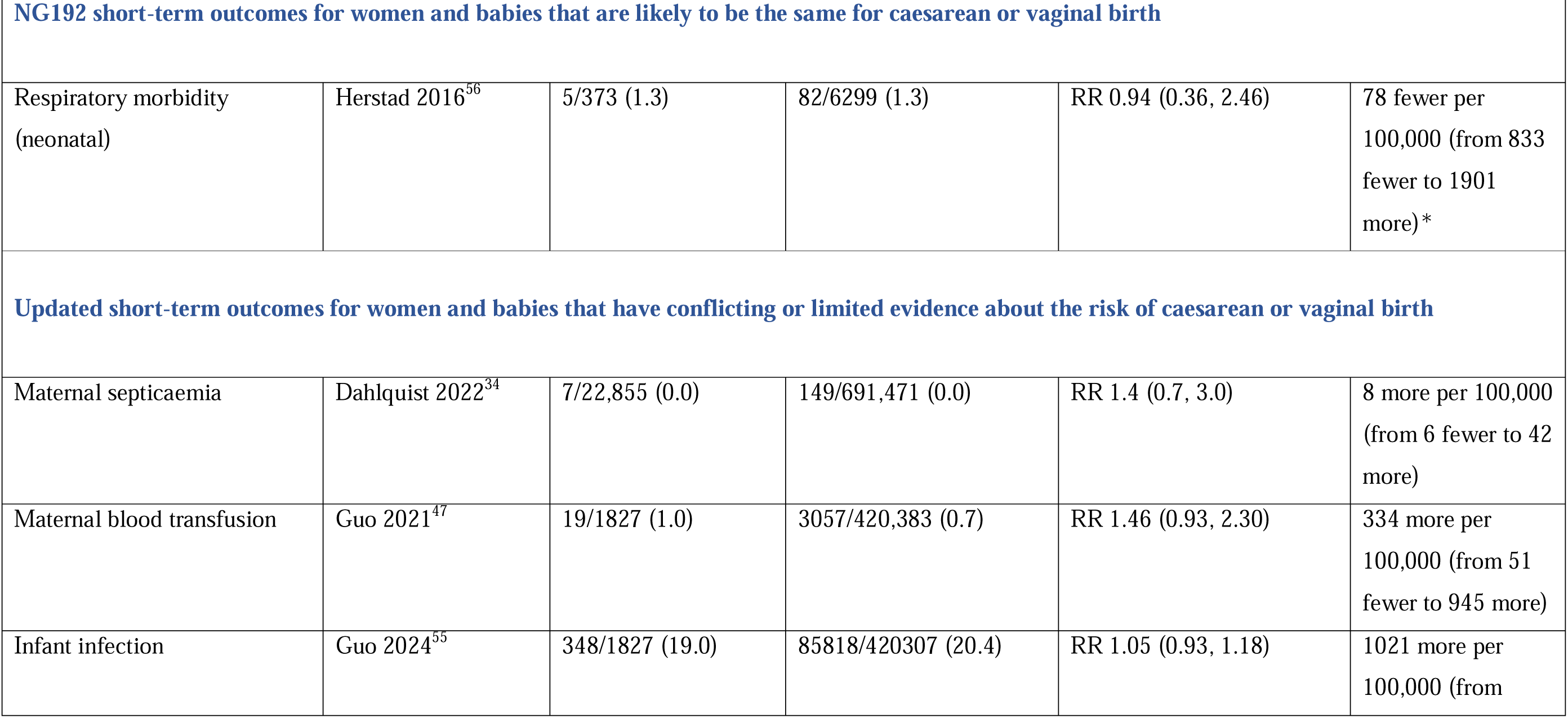
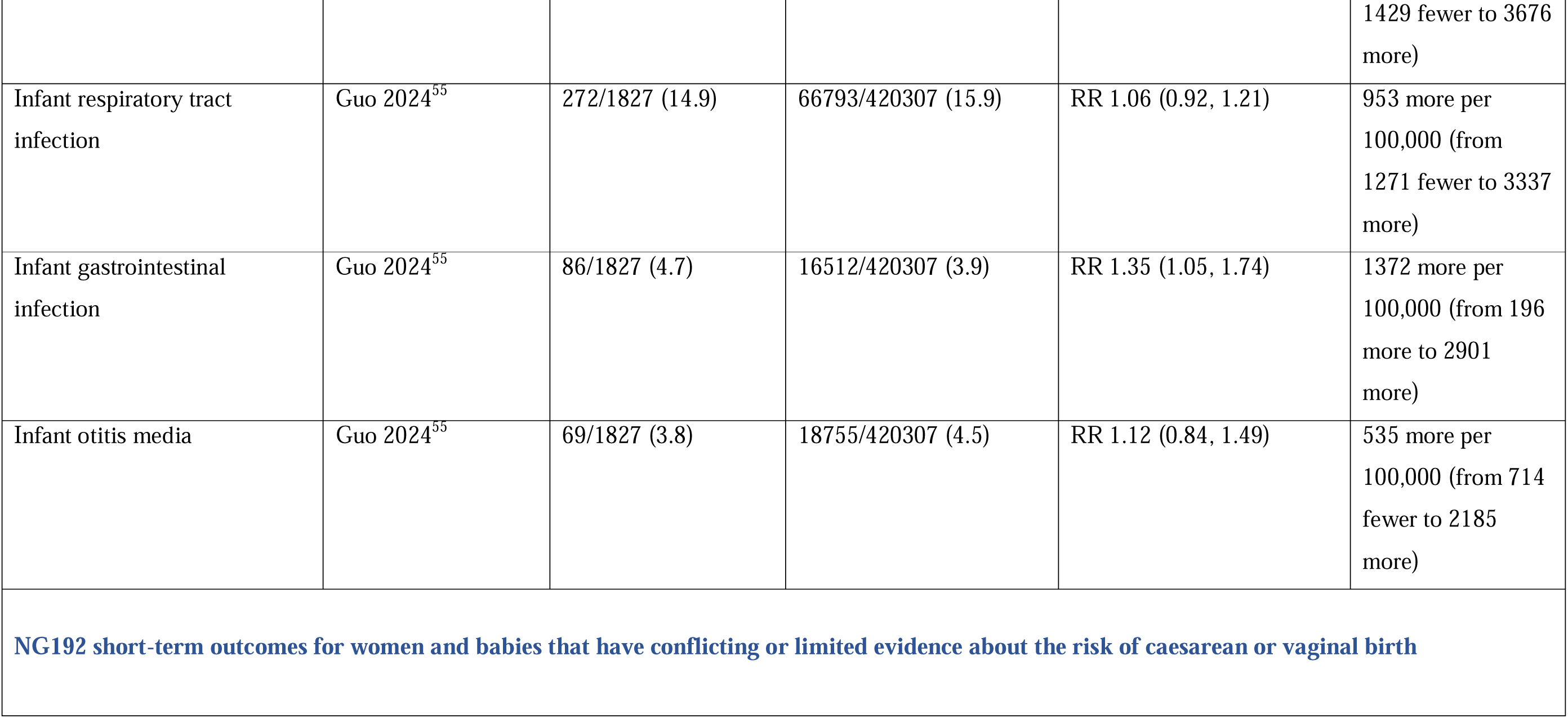
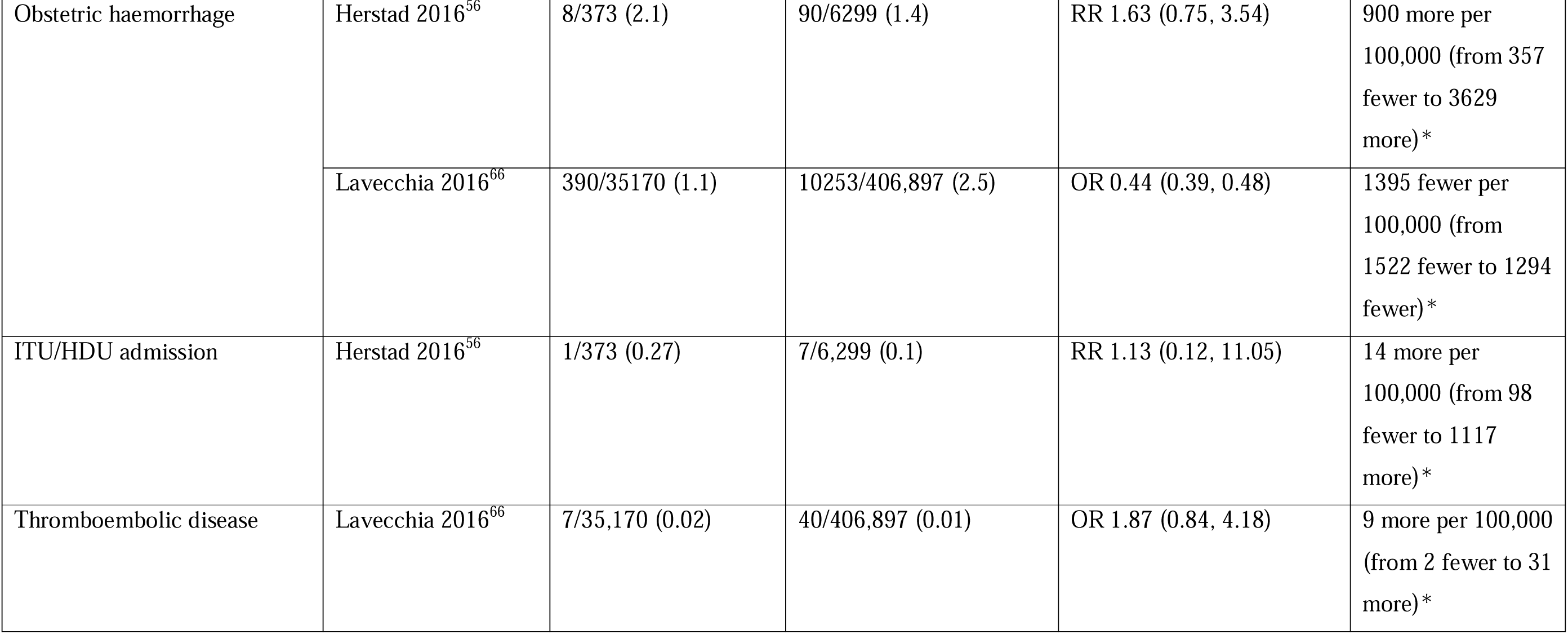
Summary of short-term outcomes of planned caesarean birth reported in NG192 and those identified in the update.

**Table 2.**
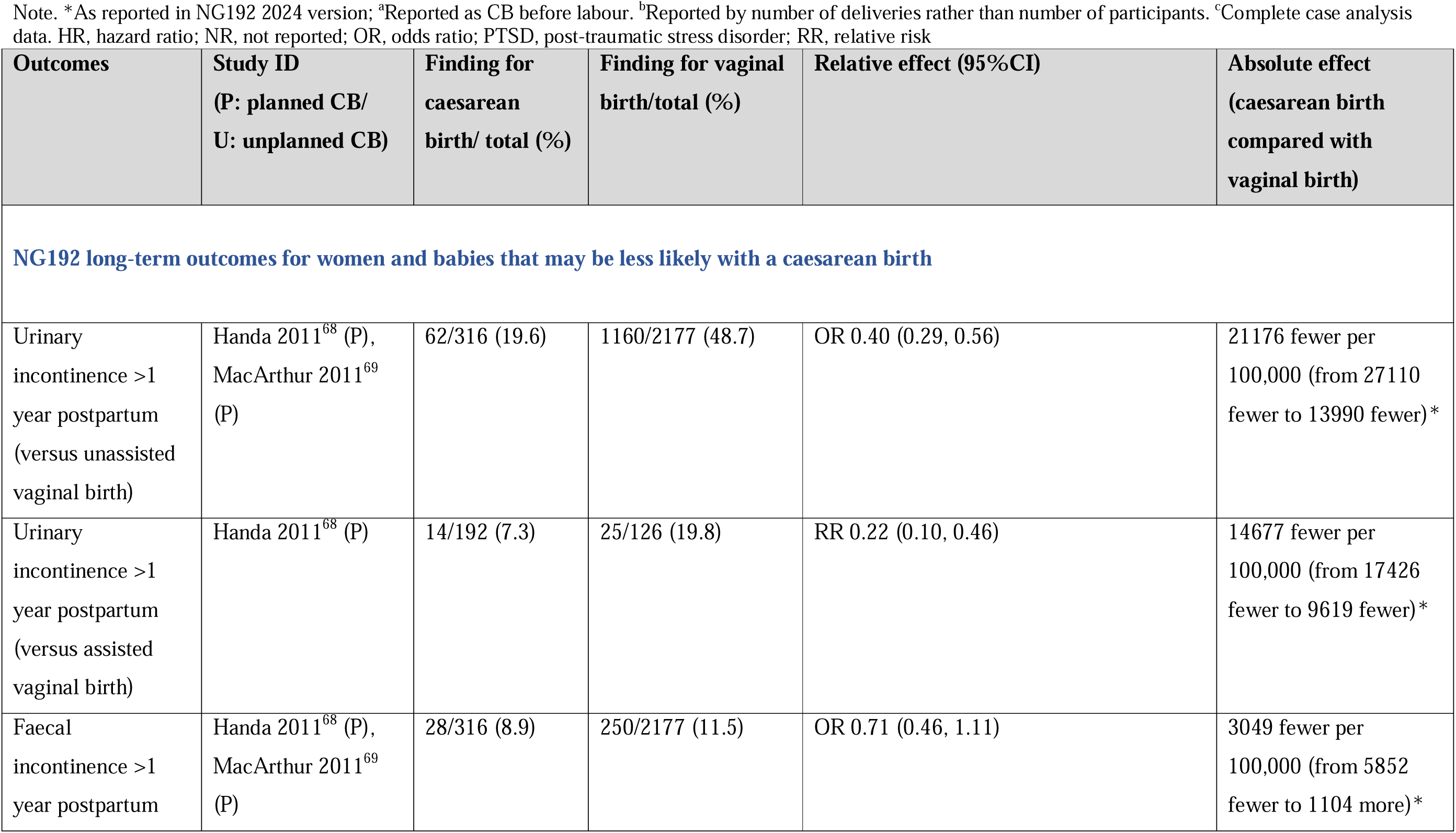
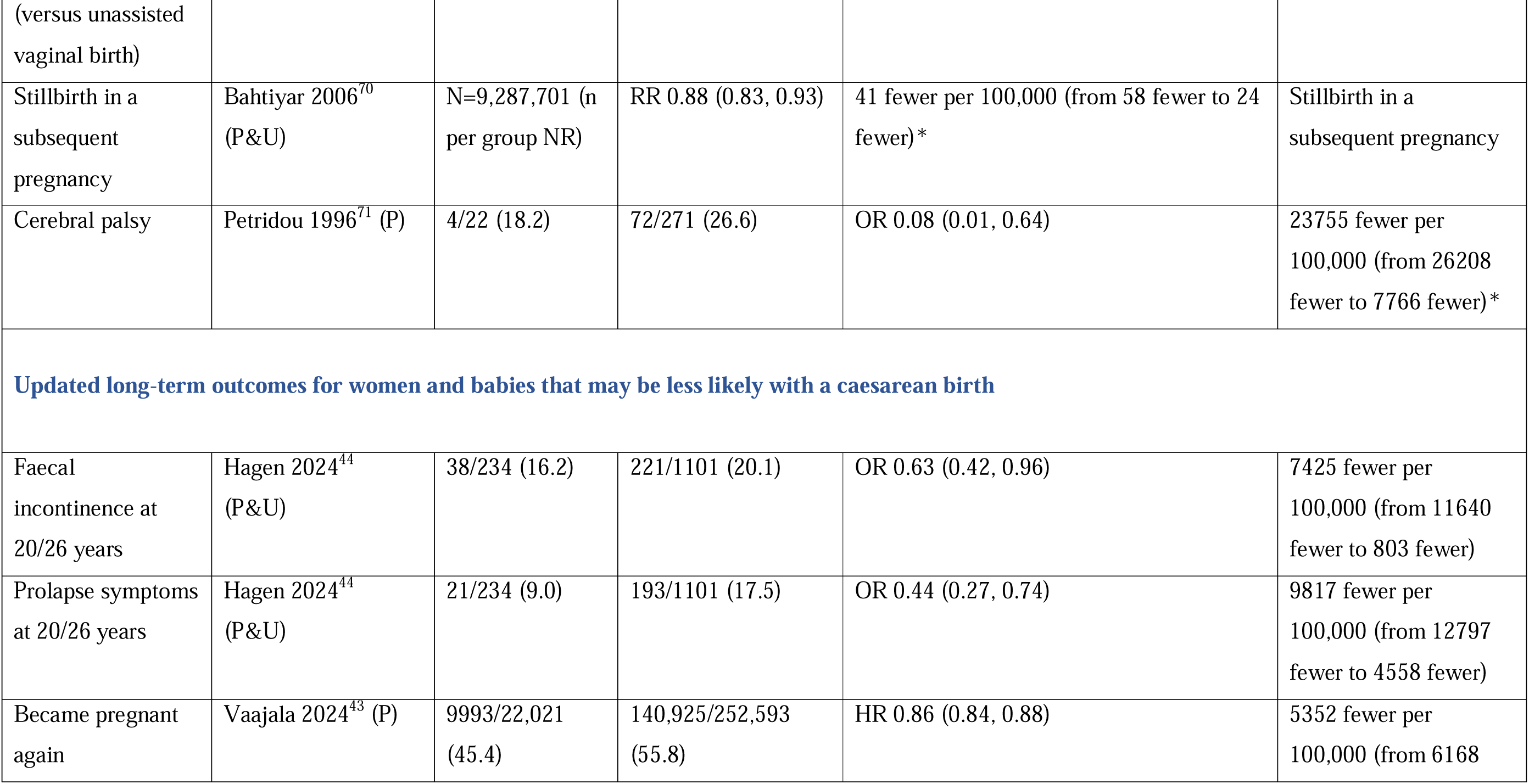
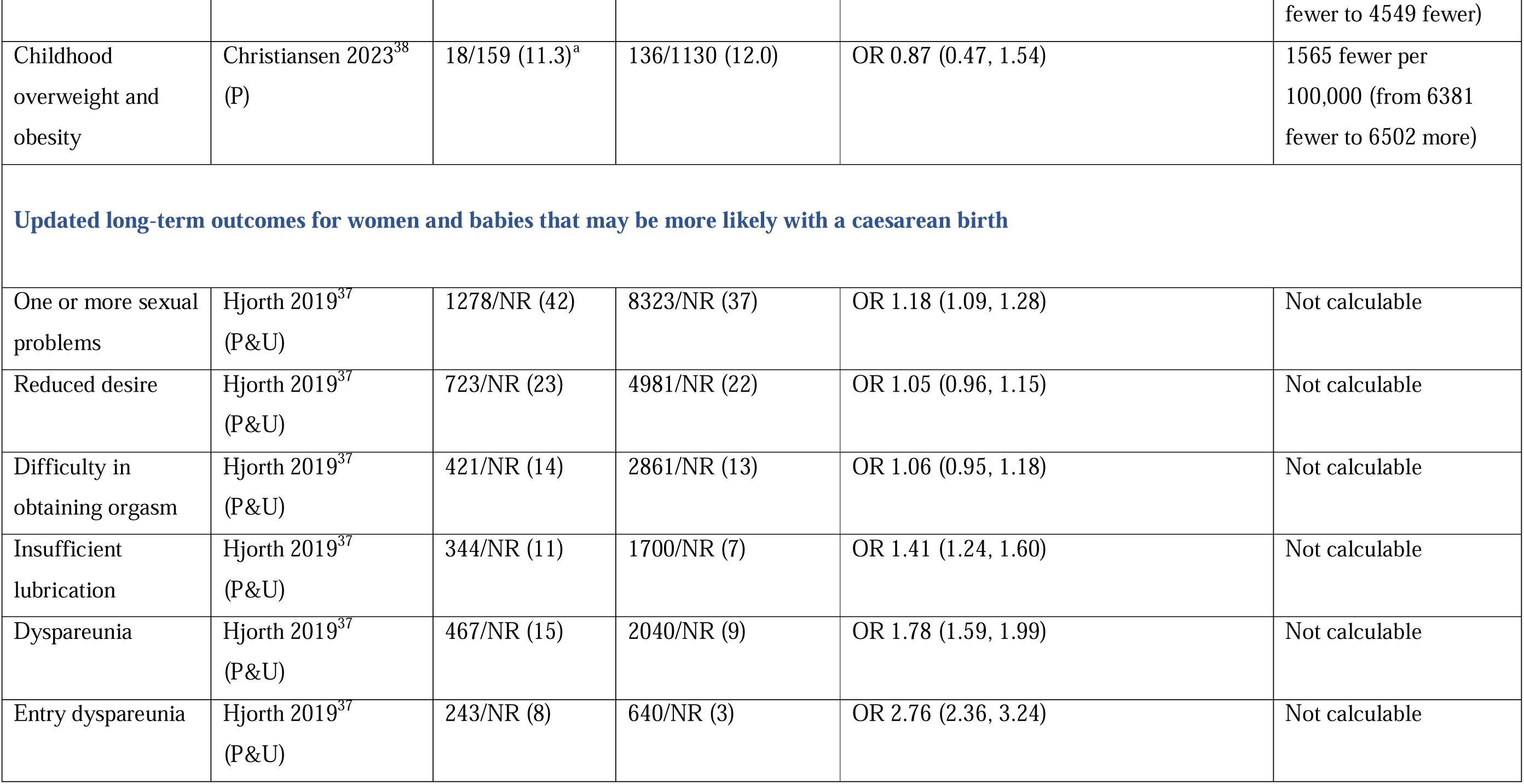
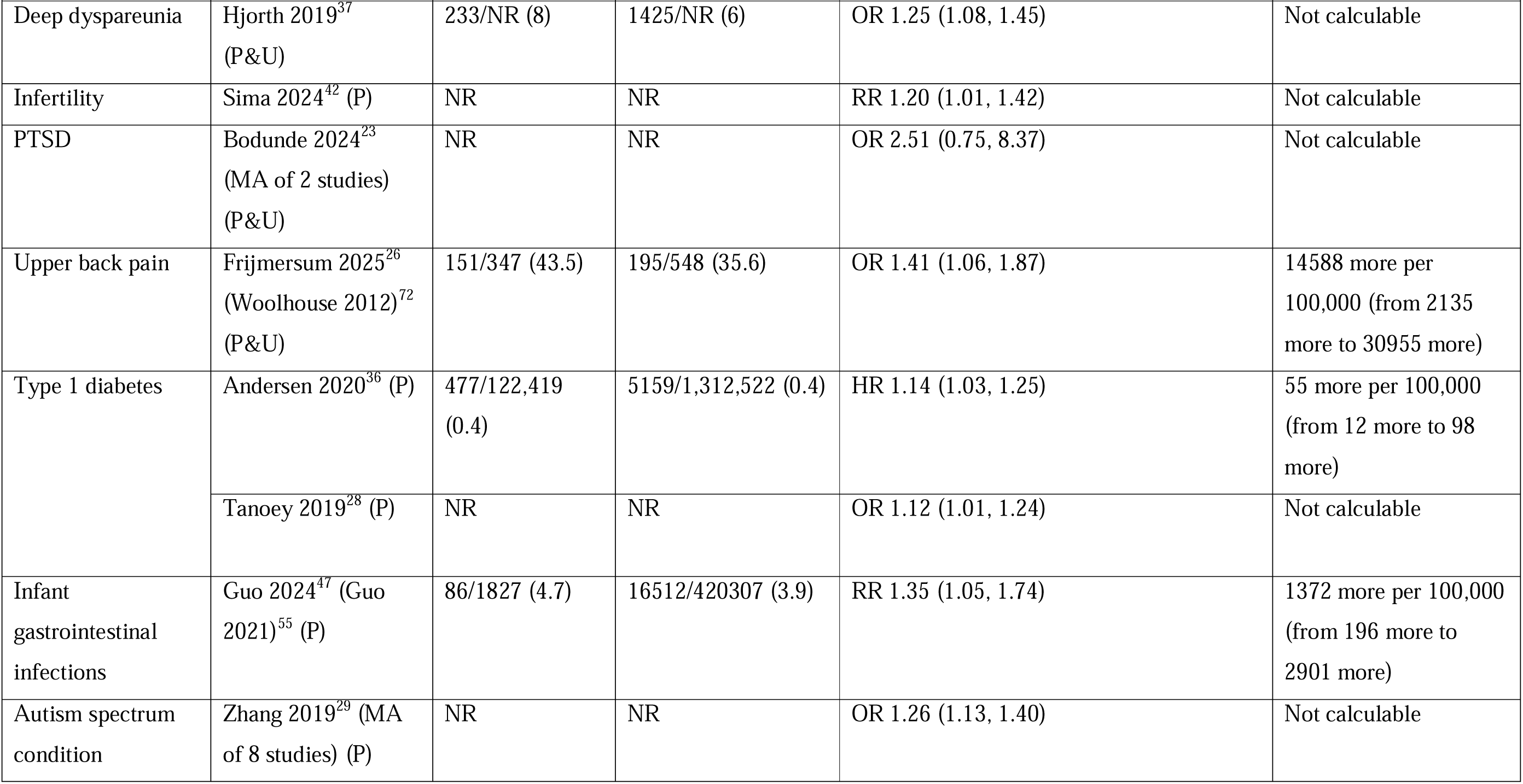
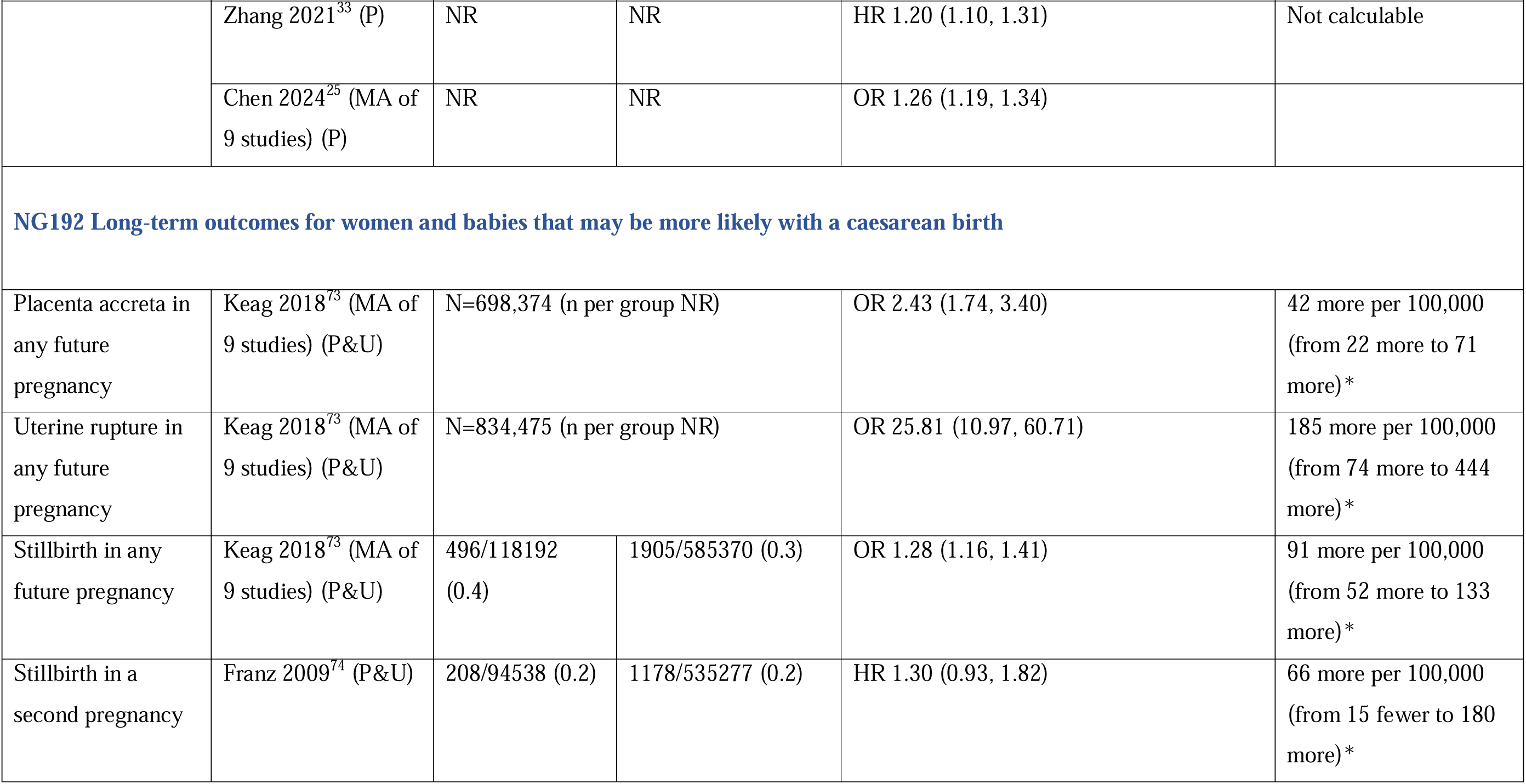
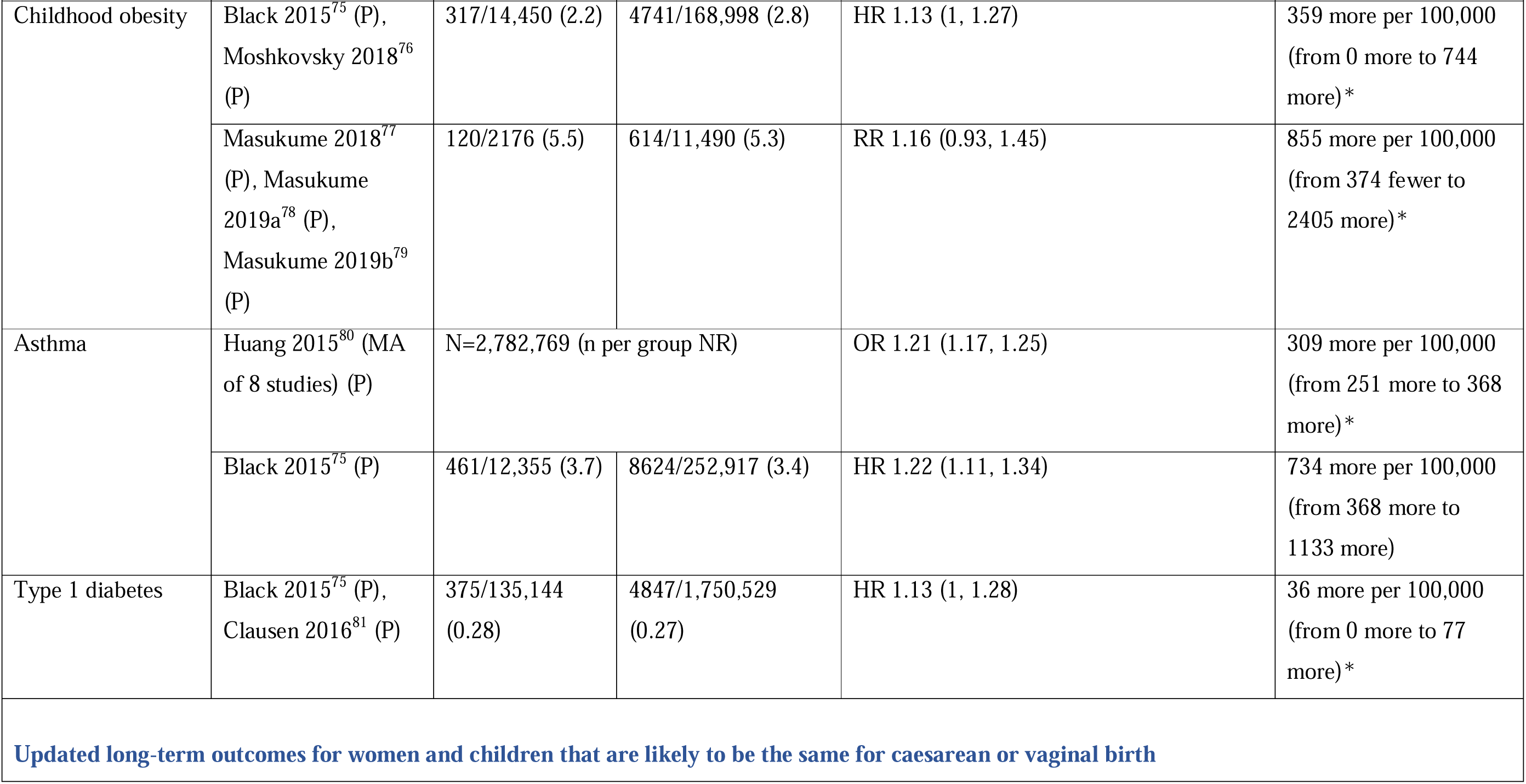
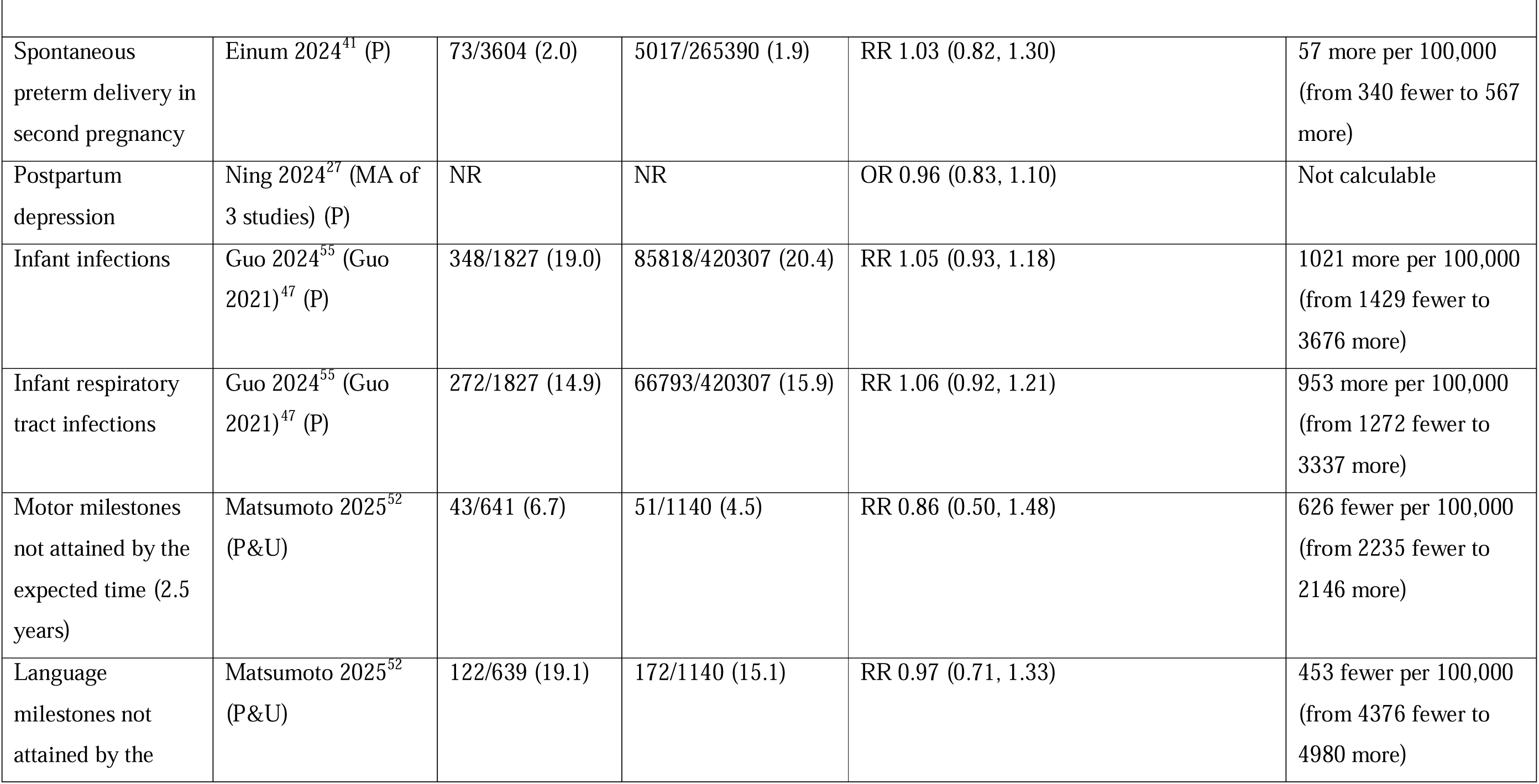
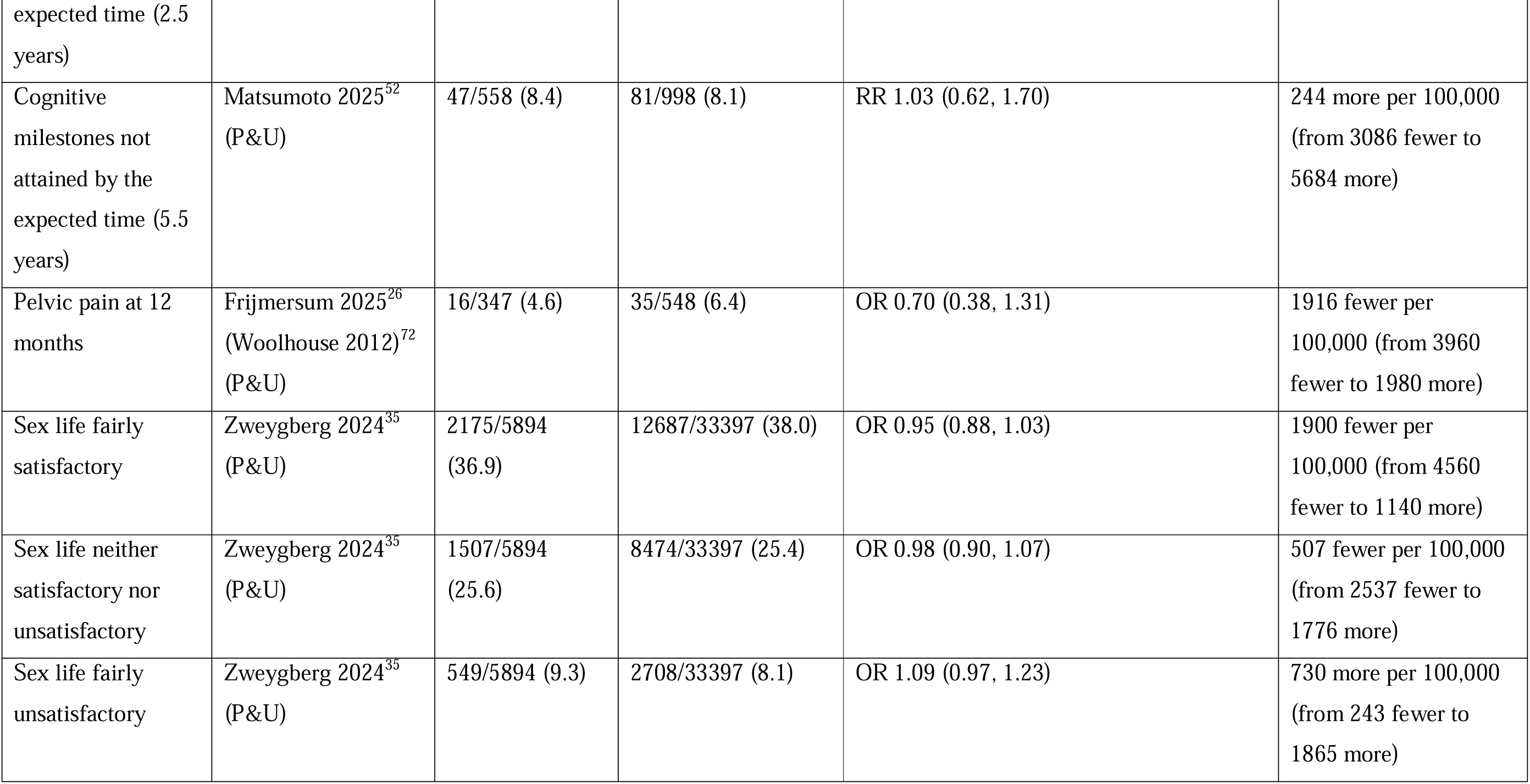
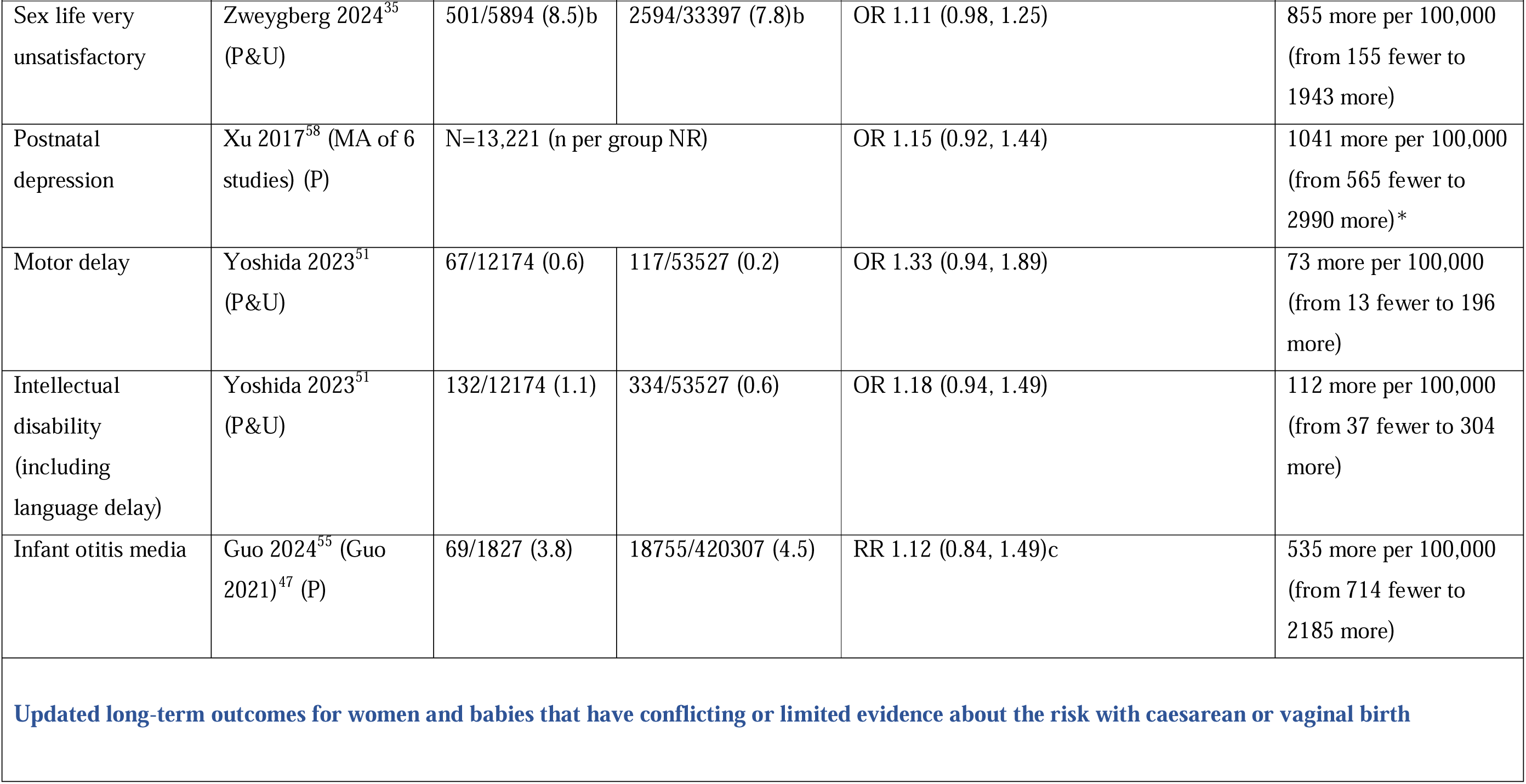
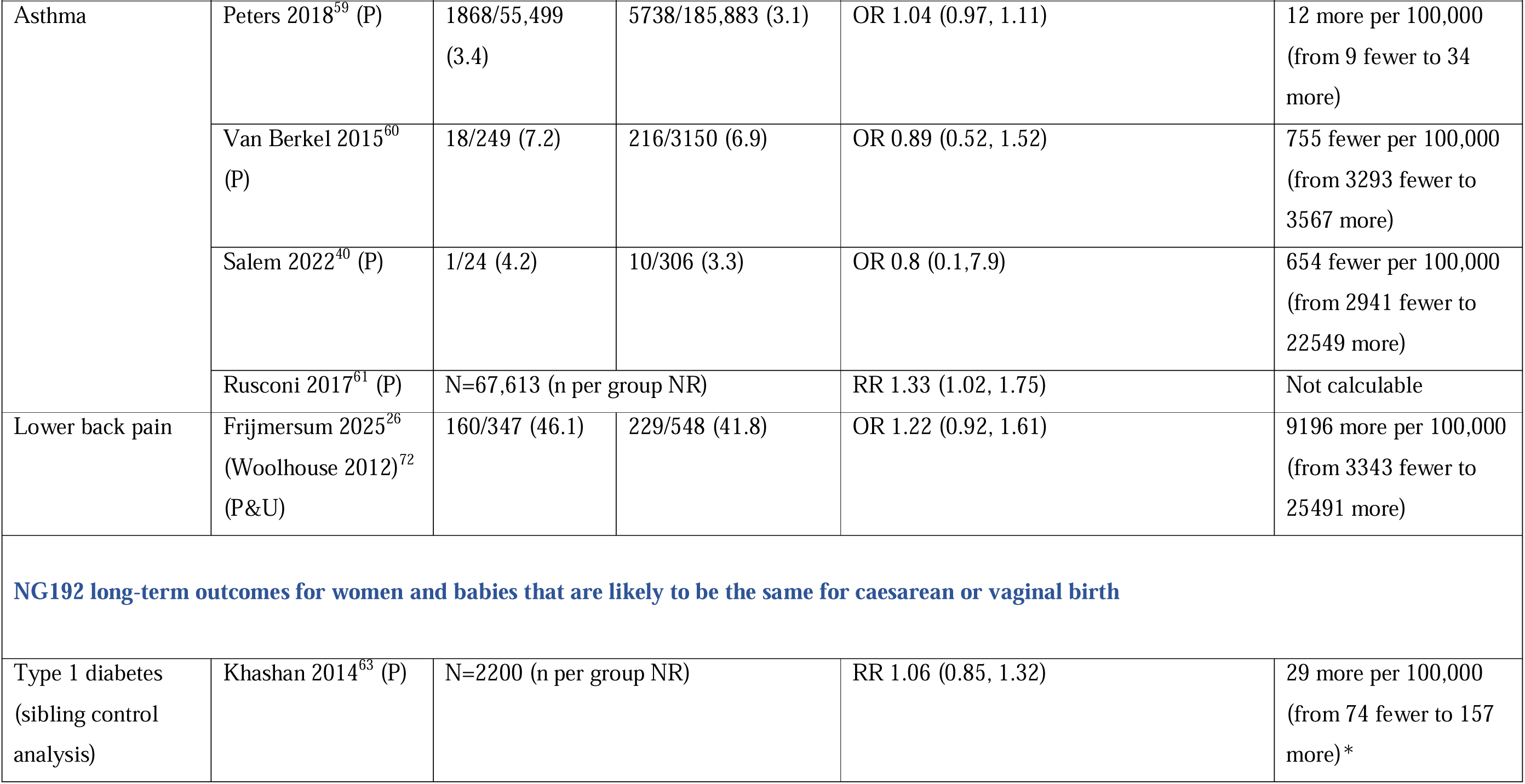
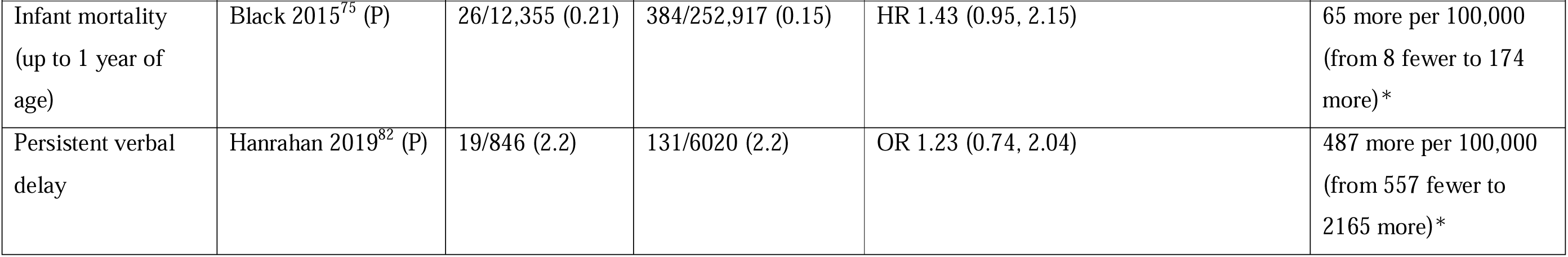
Summary of long-term outcomes of planned (or actual) caesarean birth reported in NG192 2024 and those identified in the update.

### Short-term outcomes for women

New evidence identified in this update indicates that maternal postpartum infection (16.4% vs 10.2%, RR 1.6, 95% CI 1.5-1.6) was more common following planned CB than planned VB. Higher risks were observed for specific infections, including wound infection (1.6% vs 0.5%, RR 2.7, 95% CI 2.4-3.0), mastitis (4.6% vs 2.1%, RR 2.0, 95% CI 1.9-2.2), endometritis (1.7% vs 1.4%, RR 1.2, 95% CI 1.1-1.3) and urinary tract infection (0.9% vs 0.6%, RR 1.5, 95% CI 1.3-1.7).^34^ Maternal thromboembolic disease was also more frequent following planned CB birth compared with planned VB (1.2% vs 0.7%). Increased risks were reported for pulmonary embolism (RR 1.7, 95% CI 1.0, 2.6), venous thrombosis (RR 1.3, 95% CI 1.3, 1.4), central venous thrombosis (RR 1.4, 95% CI 1.4, 1.4).^34^

New evidence also indicates that postpartum pelvic pain was less common after any CB compared with VB (2.1% vs 4.5%, OR 0.48, 95% CI 0.31-0.74).^26^ Limited new evidence suggests no statistically significant difference in the risk of septicaemia or blood transfusion between planned CB and planned VB.

Existing evidence from NG192 shows that bleeding complications (9.9% vs 5.0%, OR 2.5, 95% CI 2.1-3.0), maternal death (0.025% vs 0.004%, OR 5.63, 95% CI 2.52-12.55) and peripartum hysterectomy (0.16% vs 0.08%, OR 1.81, 95% CI 1.36, 2.40) are more common after planned CB than planned VB. NG192 reports conflicting evidence for haemorrhage-related maternal outcomes: postpartum haemorrhage was less common following planned CB (1.1% vs 2.5%, RR 0.44, 95% CI 0.39-0.48), whereas obstetric haemorrhage did not significantly differ between groups (2.1% vs 1.4%, RR 1.63, 95% CI 0.75, 3.54), nor did intensive care or high dependency care admission (0.27% vs 1.0%, RR 1.13 (0.12, 11.05).

### Short-term outcomes for babies

New evidence from this update indicates that admission to a neonatal unit is less likely (1.6% vs 3.0%, RR 0.52, 95% CI 0.36-0.74) following a plan for CB compared with a planned VB in one study.^47^ In contrast, NG192 reported no significant difference (4.3% vs 4.5%) in neonatal unit admission in a second, smaller study.^56^ Pooling of these data shows a reduced risk after planned CB with moderate heterogeneity (RR 0.61, 95% CI 0.45-0.82, I^2^ 56%).

**Figure 2.**
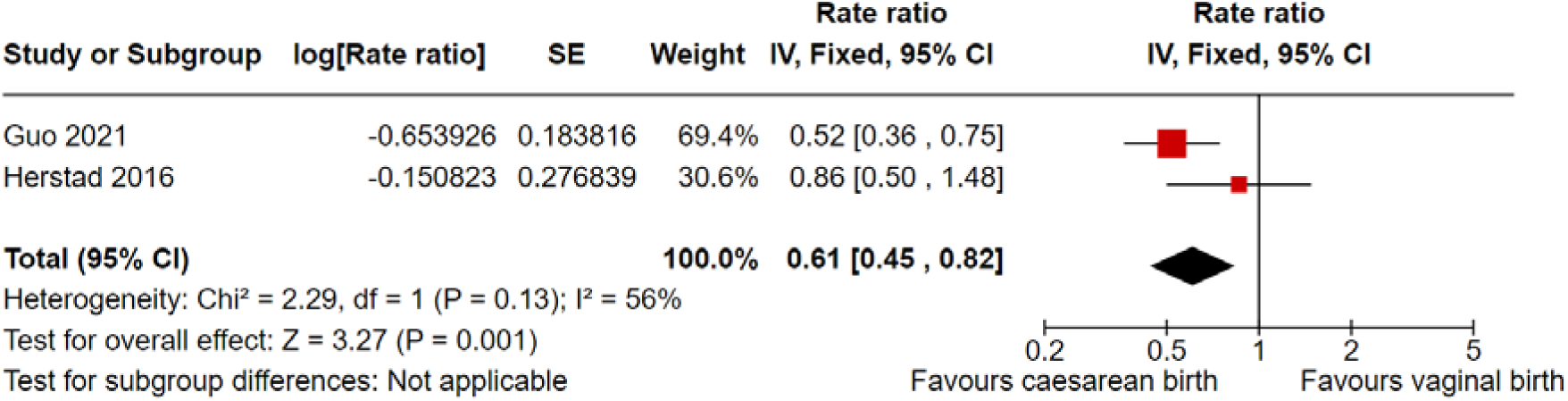
Meta-analysis of admission to a neonatal unit.

New evidence from this update shows conflicting findings regarding the likelihood of infant infection after planned CB compared with planned VB, including overall infection and specific infections of the respiratory tract, gastrointestinal tract, and otitis media. Existing findings from the NG192 evidence review indicate that neonatal infectious morbidity does not differ by planned MOB, with reported incidence ranging from 0.5 to 2.4%.^56, 57^ NG192 also reports that respiratory distress in infants is more common following planned CB than planned VB (2.7% vs 1.0% OR 2.7, 95% CI 1.8-3.9). Neonatal respiratory morbidity is reported at similar rates in both groups (1.3%).

### Long-term outcomes for women

New evidence from this update indicates that postnatal depression occurs at similar rates following planned CB and VB (OR 1.15, 95% CI 0.92-1.44 and OR 0.96, 95% CI 0.83-1.10, respectively).^27, 58^ Similarly, posttraumatic stress disorder was not found to be significantly more common following CB (any CB) compared with VB (OR 2.51, 95% CI 0.75-8.37).^23^ Musculoskeletal outcomes showed mixed findings. Lower back pain was reported at similar frequencies after CB (any CB) and VB (46.1% vs 41.8%, OR 1.22, 95% CI 0.92-1.61), whereas upper back pain was more common after CB (43.5% vs 35.6%, OR 1.41, 95% CI 1.06-1.87). Pelvic pain at 12 months did not differ significantly between birth modes (4.6% vs 6.4%, OR 0.7, 95% CI 0.38-1.31).^26^

Evidence on pelvic floor outcomes suggests potential long-term differences. Prolapse symptoms were less common after 20-26 years after CB compared with VB (9% vs 17.5%, 0.44, 95% CI 0.27-0.74).^44^ Consistent with NG192, urinary incontinence more than 1 year postpartum was less common following planned CB (range 7.3% to 19.6%) than VB (range 19.8% to 48.7%), with reported odds ratios ranging from 0.4 (95% CI 0.29-0.56) compared with unassisted VB to 0.22 (95% CI 0.1, 0.46) compared with assisted VB. NG192 also reported no significant difference in faecal incontinence at ≥1 year postpartum (8.9% vs 11.5%, OR 0.71, 95% CI 0.46-1.11). However, new evidence from this update suggests that faecal incontinence was less common 20-26 years after CB (any CB) compared with VB (16.2% vs 20.1%; 0.63, 95% CI 0.42-0.96).

Sexual health outcomes were mixed. A large study of over 39,000 women found that being ‘fairly satisfied’ with sex life was reported in similar proportions following CB (any CB) and VB (36.9% vs 38%, OR 0.95, 95% CI 0.88-1.03).^35^ It also reported sex life to be ‘fairly’ or ‘very unsatisfactory’ in similar proportions after CB and VB (8.5% vs 7.8% OR 1.11 (0.98, 1.25).^35^ A separate study of over 37,000 women found that one or more sexual problems was more common after (any) CB than after VB (42% vs 37%, OR 1.18, 95% CI 1.09-1.28)).^37^ This study reported the specific sexual problems after CB and VB respectively as reduced desire (23% vs 22%), difficulty obtaining orgasm (14% vs 13%), insufficient lubrication (11% vs 7%), dyspareunia (15% vs 9%), entry dyspareunia (8% vs 3%) and deep dyspareunia (8% vs 6%).

### Future pregnancy outcomes

New evidence identified in this update indicates that women were less likely to become pregnant again after planned CB than after VB (45.4% vs 55.8%, HR 0.86, 95% CI 0.84-0.88).^43^ Women also reported infertility more often after CB than VB (RR 1.20, 95% CI 1.01-1.42).^42^ The NG192 guideline reported mixed evidence regarding stillbirth in a future pregnancy. One study found stillbirth to be less likely after CB than after VB (RR 0.83, 95% CI 0.83-0.93), whereas another reported a higher risk following CB (OR 1.28, 95% CI 1.16-1.41). When restricted to the second pregnancy, no statistically significant difference was observed (HR 1.30, 95% CI 0.93-1.82). NG192 also reported that placenta accreta was more likely following CB (any CB) compared with VB (OR 2.43, 95% CI 1.74- 3.40) and that uterine rupture was more common after CB (OR 25.81, 95% CI 10.97, 60.71).

### Long-term outcomes for babies

New evidence identified in this update suggests that the risk of childhood asthma following CB is generally comparable to that following VB. In three of the four newly included studies, there was no significant difference in asthma risk between planned CB and VB, and all studies reported odds ratios (ORs). In the fourth study, asthma was reported to be significantly more common after planned CB.^40, 59–61^ Existing evidence reported in NG192 indicated a modestly increased risk of asthma in children born by planned CB compared with VB (3.7% vs 3.4%, HR 1.22, 95% CI 1.11-1.34), with a pooled OR estimate of 1.21 (95% CI 1.17-1.25). However, the updated meta-analysis (Figure 3) showed a pooled OR of 1.04 (95% CI 0.97-1.11, I^2^ = 0%) with low heterogeneity, indicating no statistically significant difference in asthma risk between planned CB and VB.

**Figure 3.**
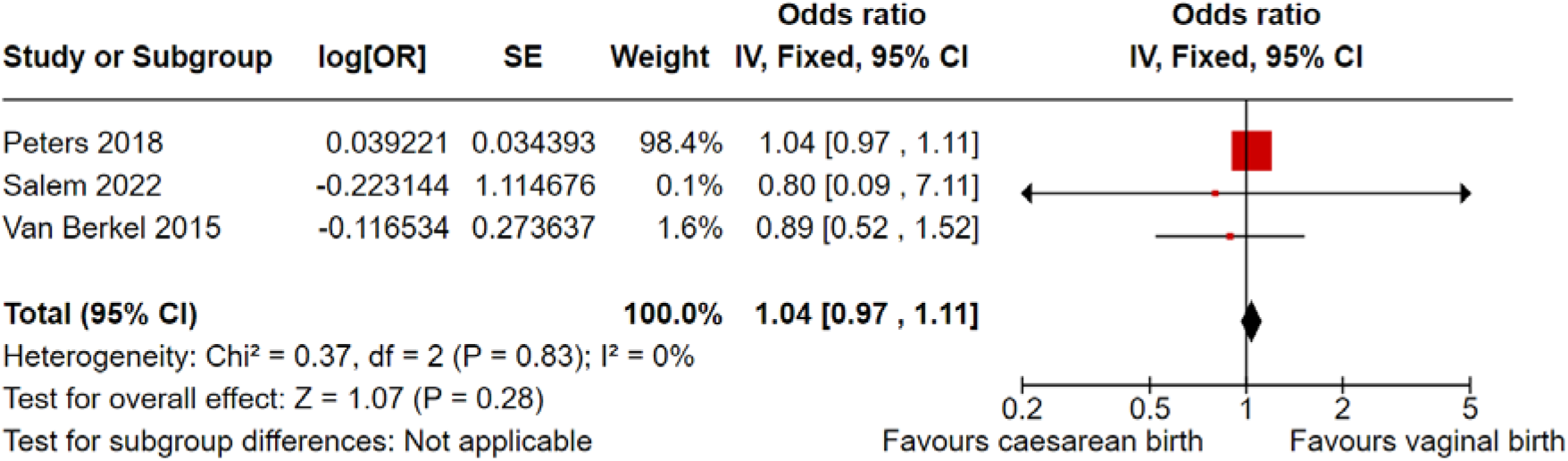
Meta-analysis of asthma (OR data)

New evidence identified in this update found that childhood overweight and obesity were no more likely after planned CB than after VB (11.3% vs 12% OR 0.87, 95% CI 0.47-1.54).^38^ The NG192 evidence review reported mixed results regarding childhood obesity, with some evidence suggesting a higher risk after planned CB than after VB (2.2% vs 2.8%, HR 1.13, 95% CI 1.0-1.27), while other evidence showed no difference between modes of birth (5.5% vs 5.3%, RR 1.16, 95% CI 0.93-1.45). NG192 also reported no statistically significant difference in the risk of infant mortality (0.21% vs 0.15% HR 1.43, 95% CI 0.95-2.15) and cerebral palsy being less likely after planned CB than VB (OR 0.08, 95% CI 0.01-0.64).

### Neurocognitive outcomes

New evidence from this update found that children’s attainment of motor milestones at the expected age was neither more nor less likely after any CB compared with VB (6.7% vs 4.5%, RR 0.86, 95% CI 0.50-1.48). The same study reported no difference in the attainment of language milestones (19.1% vs 15.1%, RR 0.97, 95% CI 0.71-1.33) or cognitive milestones (8.4% vs 8.1%, RR 1.03, 95% CI 0.62-1.70).^52^ In addition, evidence from this update indicated that motor delay (0.6% vs 0.2%) and intellectual disability (1.1% vs 0.6%) were neither more nor less likely after any CB compared with VB.^51^

### Infection

New evidence identified in this update found that overall infections during the first year of life were neither more nor less likely to occur after any CB compared with VB (RR 0.93, 95% CI 0.86-1.01 in one study^62^ and 19.0% vs 20.4% RR 1.05, 95% CI 0.93-1.18 in another study).^47^ However, urinary tract infections in the first year of life were reported to be more common (OR 3.6, 95% CI 1.1-11.1).^62^ Infant gastrointestinal infections were also more common after planned CB (4.7% vs 3.9%; RR 1.35, 95% CI 1.05-1.74) than after VB.^47^ In contrast, infant otitis media was neither more nor less common after CB compared with VB (3.8% vs 4.5%, RR 1.12, 95% CI 0.84-1.49).^47^

### Type 1 diabetes

New evidence identified in this update indicates that type 1 diabetes was more likely after planned CB than VB in two studies (0.4% in both groups, HR 1.14, 95% CI 1.03-1.25^36^ and OR 1.12, 95% CI 1.01-1.24, respectively).^28^ In contrast, a sibling- control study found no significant difference between planned CB and VB (RR 1.06, 95% CI 0.85-1.32).^63^ NG192 reported Type 1 diabetes to be more common after planned CB than after VB (0.28% vs 0.27% HR 1.13, 95% CI 1-1.28). An updated meta-analysis (Figure 4) of three studies reporting HRs found a similar pooled estimate with low heterogeneity (HR 1.13, 95% CI 1.05-1.22, I^2^ 0%).

**Figure 4.**
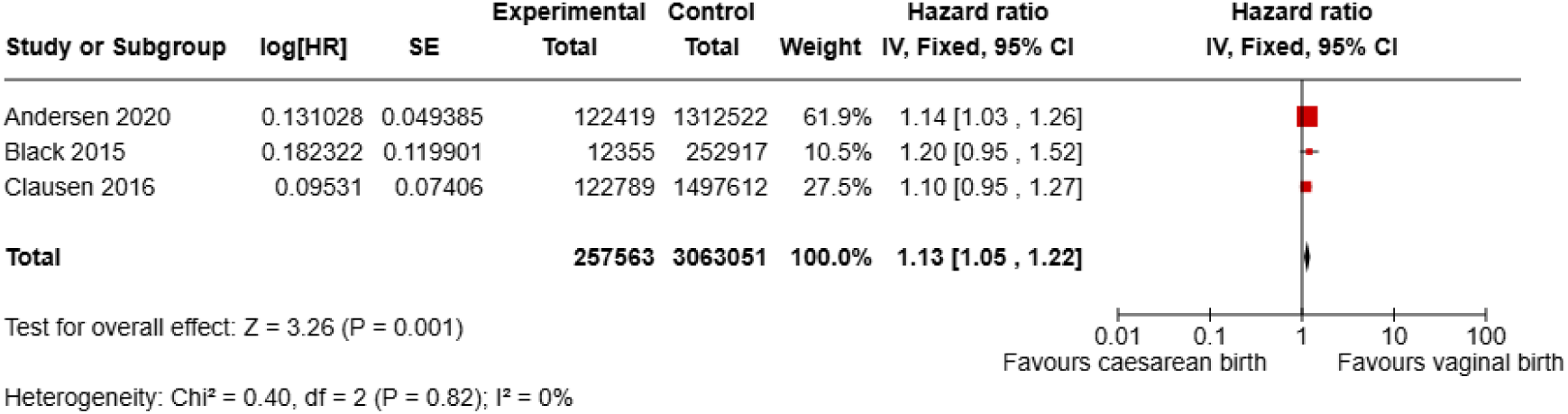
Meta-analysis of type 1 diabetes (HR data)

### Autism

New evidence identified in this update indicates that autism spectrum disorder is more common after planned CB than after VB, as reported in two studies (HR 1.20, 95% CI 1.10-1.31; OR 1.26, 95% CI 1.19-1.34).^33, 54^ A meta-analysis of two previously included studies and one newly identified study (Figure 5) found that children born after CB had an increased risk of autism spectrum disorder diagnosis over time compared with those born vaginally (pooled HR 1.15, 95% CI 1.05-1.27, I^2^ 69%).

**Figure 5.**
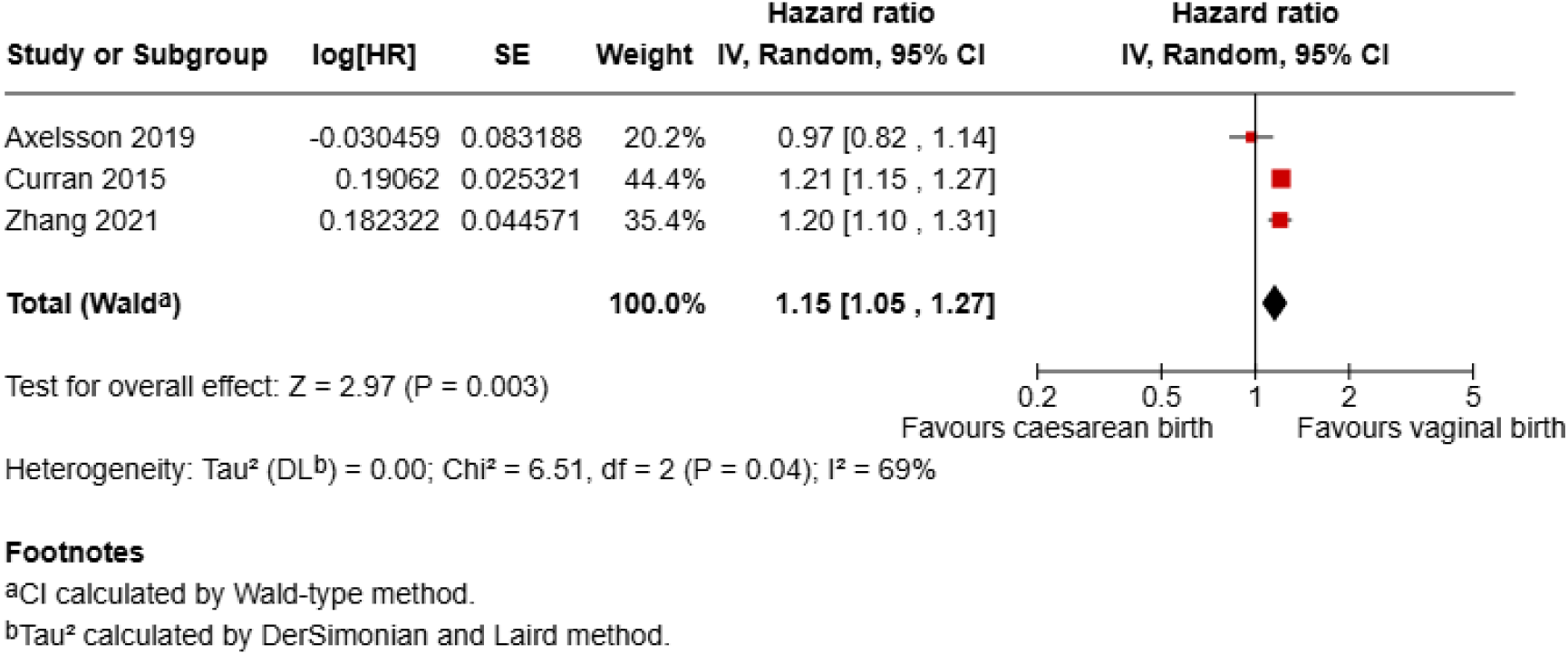
Meta-analysis of autism spectrum disorder (HR data)

## Discussion

This systematic review updates and extends the evidence underpinning the NG192 guideline on outcomes of planned CB compared with planned and actual VB.^7^ Drawing on 65 reports from 58 primary observational studies and 7 systematic reviews, reporting evidence for 36 key outcomes, it provides contemporary data to inform decision-support materials for women, clinicians and policymakers. All newly identified studies were at low risk of bias. Critically, only three new studies reported outcomes of planned CB compared to planned VB.

Among the three new studies directly comparing planned CB with planned VB, several important findings emerged. Planned CB was associated with a higher risk of maternal postpartum infection, including wound infection, mastitis, endometritis and urinary tract infection,^34^ alongside a marginally increased risk of venous thrombosis. Future pregnancy was less common, and infertility more frequent, after planned CB and any CB compared, respectively, compared with VB.^42, 43^ In contrast, planned CB was associated with lower rates of admission to the neonatal unit than planned VB.

Maternal sexual problems were more frequently reported after CB than VB, although overall satisfaction with sex life appeared similar between modes of birth.^26, 35, 37^ New evidence also suggested higher rates of infant gastrointestinal infections after planned CB compared with planned VB.^55^ For long-term outcomes, new evidence highlights conflicting findings regarding childhood asthma and type 1 diabetes following planned CB, suggesting that previously observed positive associations may be at least partly explained by confounding, supported by the sibling study design assessing type 1 diabetes risk where maternal/environmental confounding is accounted for. Findings related to autism were consistent with NG192, with both existing and new evidence indicating a higher incidence after planned CB than after VB. However, potential for substantial confounding remains and no sibling analysis has been identified to date, thus the role of parental factors in both the indication for CB and the autism diagnosis have not been accounted for.

No new evidence was identified for several rare but serious maternal outcomes reported in NG192. Existing low-quality evidence continues to suggest that cardiac arrest, maternal death and hysterectomy are more common after planned CB, and that placenta accreta and uterine rupture in subsequent pregnancy are more common after any CB.

### Strengths and limitations

This review represents the most comprehensive synthesis to date of outcomes associated with planned CB, compared with planned or actual VB, that are relevant to women, families, and healthcare professionals. Its findings will underpin information provision and shared decision-making regarding MOB from 2026 onwards. However, important evidence gaps persist. High-quality data remain scarce for outcomes of particular importance to women and health professionals, including maternal death, hysterectomy, length of hospital stay, and long-term maternal well-being. Although three recent publications have addressed key gaps in short-term outcomes following planned modes of birth, highlighting the potential of contemporary data collection, they do not yet provide a comprehensive picture.^34, 47, 55^ The limitations are even more pronounced for long-term outcomes. None of the included studies reported outcomes following planned VB; instead, outcomes were reported following actual VB or compared with any CB, combining planned and emergency procedures. This reduces the precision of comparisons and limits the applicability of findings to decision-making about *planned* MOB. When only outcomes of actual VB are reported (instead of all who planned VB), the outcomes for those who ultimately had unplanned CB are lost from the denominator, concentrating the effect of adverse VB outcomes or diluting the effect of adverse outcomes related to *unplanned* CB. When planned and unplanned/emergency CB (which usually follow a plan for VB) are combined, outcomes of emergency CB may be inappropriately attributed to all CB, including planned procedures, which may be misleading, particularly for outcomes relating to uterine damage or adverse psychological outcomes which are specific to the intrapartum nature of a CB.

We synthesised evidence on outcomes that are widely recognised as important to stakeholders involved in planning the MOB, using the same rigorous methodology as the NG192 guideline. This methodological consistency enabled direct comparison with existing recommendations and strengthened the evidence base supporting informed birth planning. Nevertheless, as with NG192, this review is constrained by the nature of the available evidence. Randomised allocation to planned CB or VB has been largely unsuccessful, meaning that all included studies were observational. Although most studies adjusted for known confounders, residual confounding remains a significant concern. Selection bias is also a possible risk. However, most studies were population-based, selected relatively ‘low risk’ populations and reported routinely collected data. For specific outcomes, interpretation is further limited by heterogeneity. For example, although neonatal unit admission rates were consistent in direction of effect across recent large studies, we recognise that admission thresholds vary substantially by local policy and models of care provision. This limits generalisability, particularly to the UK settings. No new evidence was identified for postpartum haemorrhage, leaving unresolved the conflicting NG192 findings relating to poorly defined ‘obstetric haemorrhage’ and ‘bleeding complications’ following CB.^57^ For several outcomes with mixed findings, meta-analyses would have been desirable but were precluded by heterogeneity observed across study designs.

### Implications for clinicians and policymakers

The reported findings suggest that planned VB offers advantages to women in the short-term, including lower risk of infection and venous thrombosis, and longer-term benefits related to fertility and reduced risk of serious complications in subsequent pregnancies. These associations are biologically plausible given the known links between surgery, scarring and adverse reproductive outcomes.

Meanwhile, planned CB appears to offer advantages in short-term neonatal outcomes, notably reduced neonatal unit admission, with potential implications for less resource use and less family separation. However, as neonatal unit admissions are influenced by gestation at birth and local admission criteria, the applicability of findings will vary by context. Planned CB is usually recommended from 39 completed weeks gestation, but early term CB (at 37-38 weeks gestation) is not uncommon and is linked to higher risk of neonatal respiratory morbidity.^64^ Planned CB was also associated with less maternal pelvic pain and lower long-term risk of incontinence and prolapse, although sexual problems were reported more frequently after any CB than after VB. While most of these findings align with previous research, the increased sexual problems after CB are relatively novel and may reflect causal mechanisms, such as pelvic adhesions, or residual confounding such as women with pre-existing concerns about sexual function being more likely to plan a CB.

In the context of women-centered decision-making, these findings should inform balanced discussion about planned mode of birth options and should inform content of information resources. However, the findings must be translated into accessible information that communicates absolute risks and clearly conveys uncertainty around any causal links.

### Unanswered questions and future research

Substantial gaps remain in understanding the maternal psychological sequelae of planned MOB. Although one meta-analysis found no difference in post-traumatic stress disorder symptoms between CB and VB, planned and unplanned CB were combined, obscuring potentially important distinctions. Psychological outcomes may be strongly influenced by labour events or by not achieving a preferred MOB, but these factors remain underexplored. The journey from planning a MOB to experiencing the outcomes is peppered with predictable and unpredictable events, which may act as confounders. A fundamental challenge in this field is the limited routine recording of the *planned* MOB in population datasets. Most studies rely on proxy measures or inferred intentions to establish the presumed planned MOB. Strengthening the evidence base will require systematic data collection, thereby increasing confidence in whether CB or VB was planned. Finally, safety concerns around planned VB, including the risk of hypoxic ischaemic encephalopathy and cerebral palsy, remain poorly addressed, with no eligible cohort studies reporting on these outcomes. While the populations included in studies assessing planned CB are broadly representative of the UK population, important differences exist across countries, particularly in fertility patterns, sexual health and unplanned CB rates. Greater attention to scrutinising UK data, with detailed information on CB indications and maternal risk factors, is essential to enable more precise and contextually relevant conclusions about the benefits and risks of planned VB and CB.

## Conclusion

This review provides contemporary information that could be used to inform decision-support materials on outcomes of planned CB compared with planned and actual VB for service-users and health professionals. The existing evidence base lacks data on long-term outcomes of planned VB, with studies only reporting outcomes of actual VB.

## Data Availability

All data produced in the present work are contained in the manuscript

## Acknowledgements

Dr Neil Scott provided advice on interpretation of published results and calculation of absolute risk reduction values. Mrs Gillian Taylor provided community midwifery perspective input to project planning and interpretation of findings.

## Supplementary files list

Supplementary file S1 – Literature searches

Supplementary file S2 - Characteristics of included studies (Table S1 and S2)

Supplementary file S3 - The study level summary of the quality assessment of the included studies (Tables S3 and S4)

Supplementary file S4 - Clinical outcomes identified during the review that did not meet NG192 or Delphi inclusion criteria (Tables S5 and S6)

## Footnotes

1 *From:* Page MJ, McKenzie JE, Bossuyt PM, Boutron I, Hoffmann TC, Mulrow CD, et al. The PRISMA 2020 statement: an updated guideline for reporting systematic reviews. BMJ 2021;372:n71. doi: 10.1136/bmj.n71

