## Supplementary file S1 for "Outcomes of planned caesarean birth compared with planned or actual vaginal birth: an update and expansion of the NICE Caesarean Birth Guideline systematic review NG192"

Ovid **MEDLINE**(R) and Epub Ahead of Print, In-Process, In-Data-Review & Other Non-Indexed Citations, Daily and Versions <1946 to December 19, 2022>

1	META-ANALYSIS/	172662

2	META-ANALYSIS AS TOPIC/	21944

3	(meta analy* or metanaly* or metaanaly*).ti,ab.	254746

4	((systematic* or evidence*) adj2 (review* or overview*)).ti,ab.	317730

5	(reference list* or bibliograph* or hand search* or manual search* or relevant journals).ab.	52554

6	(search strategy or search criteria or systematic search or study selection or data extraction).ab.	76988

7	(search* adj4 literature).ab.	91775

8	(medline or pubmed or cochrane or embase or psychlit or psyclit or psychinfo or psycinfo or cinahl or science citation index or bids or cancerlit).ab.	338383

9	cochrane.jw.	16171

10	or/1-9	631907

11	randomized controlled trial.pt.	583048

12	controlled clinical trial.pt.	95138

13	pragmatic clinical trial.pt.	2168

14	randomi#ed.ab.	700009

15	placebo.ab.	234125

16	randomly.ab.	397865

17	CLINICAL TRIALS AS TOPIC/	200659

18	trial.ti.	275852

19	or/11-18	1536317

20	COHORT STUDIES/	322828

21	cohort?.ti,ab.	804816

22	FOLLOW-UP STUDIES/	688855

23	(Follow$ up adj3 (study or studies)).ti,ab.	81194

24	LONGITUDINAL STUDIES/	162033

25	longitudinal$.ti,ab.	327112

26	PROSPECTIVE STUDIES/	646301

27	prospective$.ti,ab.	837291

28	RETROSPECTIVE STUDIES/	1080471

29	retrospective$.ti,ab.	971210

30	OBSERVATIONAL STUDY/	135886

31	observational$.ti,ab.	246840

32	or/20-31	3506343

33	CASE-CONTROL STUDIES/	325149

34	case control$.ti,ab.	152922

35	33 or 34	378080

36	REGISTRIES/	106749

37	(registry or registries).ti,ab.	166551

38	36 or 37	210785

39	CROSS-SECTIONAL STUDIES/	450416

40	cross sectional.ti,ab.	481987

41	39 or 40	619851

42	exp CESAREAN SECTION/	51850

43	(c?esar#an$ or c section$ or csection$ or (deliver$ adj3 abdom$)).ti,ab.	72656

44	42 or 43	85449

45	LABOR, INDUCED/	10085

46	(induc$ adj3 (labo?r$ or birth$ or born or deliver$)).ti,ab.	16800

47	CERVICAL RIPENING/	1289

48	(cervi$ adj3 ripen$).ti,ab.	2119

49	exp EXTRACTION, OBSTETRICAL/	3616

50	((extract$ or vacuum$) adj3 (birth$ or born or deliver$ or obstetric$)).ti,ab.	2626

51	(vacuum$ adj3 extract$).ti,ab.	2082

52	ventouse?.ti,ab.	263

53	OBSTETRICAL FORCEPS/	1710

54	forcep?.ti,ab.	12640

55	(instrument$ adj3 deliver$).ti,ab.	2443

56	NATURAL CHILDBIRTH/	2473

57	((natural$ or unassisted or un-assisted) adj3 (birth$ or born or deliver$)).ti,ab.	2655

58	(spontaneous$ adj3 (birth$ or born or deliver$)).ti,ab.	9066

59	VAGINAL BIRTH AFTER CESAREAN/	1874

60	((vagina$ or cephalic$) adj1 (birth$ or born or deliver$)).ti,ab.	22541

61	VBAC.ti,ab.	800

62	or/45-61	68893

63	*DELIVERY, OBSTETRIC/mt [Methods]	3571

64	(mode? adj3 (birth? or deliver$)).ti,ab.	20054

65	63 or 64	22930

66	((maternal$ or mother$ or wom?n?) adj5 short$ adj5 term adj5 outcome?).ti,ab.	270

67	URINARY BLADDER/in [Injuries]	2342

68	(bladder? adj3 injur$).ti,ab.	2404

69	exp INTESTINE, LARGE/in [Injuries]	4987

70	(bowel? adj3 injur$).ti,ab.	2488

71	URETER/in [Injuries]	2096

72	(ureter$ adj3 injur$).ti,ab.	2375

73	HEMORRHAGE/	80389

74	UTERINE HEMORRHAGE/	10060

75	POSTPARTUM HEMORRHAGE/	8233

76	((major or moderate$ or severe$) adj5 (h?emorrhag$ or (blood adj2 (loss or lose or losing)) or bleed$)).ti,ab.	45860

77	((postpartum or post-partum) adj3 (h?emorrhag$ or (blood adj2 (loss or lose or losing)) or bleed$)).ti,ab.	8907

78	((>1000ml or >1000 ml or >1000millilit$ or >1000 millilit$) adj3 (h?emorrhag$ or (blood adj2 (loss or lose or losing)) or bleed$)).ti,ab.	452

79	MOTHERS/ and PATIENT SATISFACTION/	614

80	MOTHERS/ and "QUALITY OF LIFE"/	838

81	((maternal or mother?) adj5 satisf$).ti,ab.	2068

82	"health related quality of life".ti,ab.	54958

83	HRQOL?.ti,ab.	21261

84	MATERNAL DEATH/	1038

85	MATERNAL MORTALITY/	11223

86	((maternal$ or mother?) adj5 (death? or mortalit$)).ti,ab.	29296

87	PATIENT ADMISSION/ and exp INTENSIVE CARE UNITS/	3132

88	((Intensive Therapy Unit? or ITU? or High Dependency Unit? or HDU? or Intensive care or ICU or PICU or NICU) adj5 admi$).ti,ab.	60271

89	PERIPARTUM PERIOD/ and HYSTERECTOMY/	134

90	PERIPARTUM PERIOD/ and HYSTERECTOMY, VAGINAL/	0

91	((peripart$ or peri-part$) adj3 hysterectom$).ti,ab.	454

92	exp THROMBOSIS/	141393

93	exp THROMBOEMBOLISM/	63111

94	thrombo$.ti,ab.	409312

95	((maternal$ or mother$ or wom?n?) adj5 long$ adj5 term adj5 outcome?).ti,ab.	847

96	PLACENTA ACCRETA/	2814

97	PLACENTA/ab [Abnormalities]	461

98	placenta$ accreta.ti,ab.	2337

99	(morbid$ adj3 adher$ adj3 placenta$).ti,ab.	295

100	(abnormal$ adj3 inva$ adj3 placenta$).ti,ab.	279

101	UTERINE RUPTURE/	4251

102	(uter$ adj3 ruptur$).ti,ab.	5215

103	STILLBIRTH/	6081

104	stillbirth?.ti,ab.	14231

105	ABORTION, SPONTANEOUS/	21711

106	ABORTION, HABITUAL/	7670

107	miscarr$.ti,ab.	17312

108	(abort$ adj3 (spontaneous$ or habitual$)).ti,ab.	13506

109	URINARY INCONTINENCE/	24129

110	URINARY INCONTINENCE, STRESS/	12588

111	((stress$ or mix$ or effort$ or urin$) adj3 incontinen$).ti,ab.	32407

112	FECAL INCONTINENCE/	10730

113	(f?ecal$ adj3 incontinen$).ti,ab.	7664

114	DEPRESSION, POSTPARTUM/	7143

115	(depress$ adj5 (postnatal$ or post-natal$ or postpartum or post-partum)).ti,ab.	9603

116	PND.ti,ab.	5605

117	STRESS DISORDERS, POST-TRAUMATIC/	39973

118	((post-trauma$ or posttrauma$) adj3 stress$ adj3 disorder?).ti,ab.	36714

119	PTSD.ti,ab.	30622

120	((neonat$ or baby or babies or infant?) adj5 short$ adj5 term adj5 outcome?).ti,ab.	666

121	PERINATAL MORTALITY/	2444

122	(perinatal$ adj5 (death? or mortalit$)).ti,ab.	17832

123	((stillbirth or mortalit$) adj5 (one or "1" or two or "2" or three or "3" or four or "4" or five or "5" or six or "6" or seven or "7") adj3 day?).ti,ab.	14214

124	PATIENT ADMISSION/ and INTENSIVE CARE UNITS, NEONATAL/	332

125	((baby or babies or neonat$) adj5 care unit? adj5 admi$).ti,ab.	6140

126	(NICU adj5 admi$).ti,ab.	3435

127	RESPIRATORY DISTRESS SYNDROME, NEWBORN/	13736

128	(respirat$ adj3 distress$ adj3 (baby or babies or neonat$)).ti,ab.	3211

129	(respirat$ adj3 morbidit$).ti,ab.	3073

130	HYPOXIA-ISCHEMIA, BRAIN/	6809

131	(hypoxi$ adj3 ischemi$ adj3 (encephalop$ or brain? or cerebral$)).ti,ab.	7117

132	PERIPHERAL NERVE INJURY/	7773

133	exp BRACHIAL PLEXUS/in [Injuries]	5753

134	PHRENIC NERVE/in [Injuries]	508

135	FACIAL NERVE INJURIES/	2130

136	(nerve? adj3 (injur$ or trauma$)).ti,ab.	34496

137	(brachial plexus adj3 (injur$ or trauma$)).ti,ab.	3424

138	exp INTRACRANIAL HEMORRHAGES/	78812

139	((intracranial or brain or cerebral or subarachnoid) adj3 (h?emorrhag$ or bleed$)).ti,ab.	57353

140	(extracranial adj3 (h?emorrhag$ or bleed$)).ti,ab.	322

141	(cranial adj3 (h?emorrhag$ or bleed$)).ti,ab.	440

142	exp INFANT, NEWBORN/ and INFECTION/	2841

143	(infect$ adj3 morbidit$).ti,ab.	6192

144	((baby or babies or infant? or preschool$ or pre-school$ or child$ or adolescen$ or teenage$) adj5 long$ adj5 term adj5 outcome?).ti,ab.	3130

145	INFANT DEATH/	323

146	INFANT MORTALITY/	29859

147	((infant? or neonat$ or baby or babies) adj5 (death? or mortalit$)).ti,ab.	51518

148	CHILD MORTALITY/	2917

149	(child$ adj5 (death? or mortalit$)).ti,ab.	33899

150	CEREBRAL PALSY/	23187

151	((cerebral or brain or central) adj3 (pals$ or paralys?s or pares?s)).ti,ab.	27123

152	exp NEURODEVELOPMENTAL DISORDERS/	205316

153	(neurodevelopment$ or neuro-development$).ti,ab.	45986

154	((development$ or intellect$ or communicat$ or expressive$ or receptive$ or learning or academic$ or arith$ or numer$ or math$ or read$ or write or writing or litera$ or spell$ or motor skill? or motor function$ or coordination or co-ordination or hyperkinetic$ or hyper-kinetic$ or clumsy child$) adj3 (disab$ or disorder? or difficult$ or impair$ or delay$)).ti,ab.	157857

155	(Asperger? or Kanner? or dyscalculi$ or acalculi$ or dyslexi$ or alexi$ or word blind$).ti,ab.	14873

156	(PDD or PDD-NOS or DCD or SDDMF).ti,ab.	7284

157	COGNITION DISORDERS/	66378

158	(cognit$ adj3 (disab$ or disorder? or difficult$ or impair$ or delay$)).ti,ab.	114691

159	exp COMMUNICATION DISORDERS/	68193

160	((speech or speak$ or language?) adj3 (disab$ or disorder? or difficult$ or impair$ or delay$)).ti,ab.	24803

161	(Dysglossi$ or cluttering? or verbal fluency disorder? or Rhinolali$ or dyslali$ or aprosodi$ or Aphasi$ or Articulation Disorder? or Dysarthri$ or Echolali$ or mute or Mutism? or Stutter$ or Agraphi$ or Anomi$ or Dyslexi$ or Alexi$).ti,ab.	44724

162	exp PSYCHOMOTOR DISORDERS/	14404

163	((Psychomotor or psycho-motor) adj3 (disab$ or disorder? or difficult$ or impair$ or delay$)).ti,ab.	3167

164	(Dyspraxi$ or apraxi$).ti,ab.	4903

165	exp PSYCHOLOGICAL TESTS/ and (neurodevelopment$ or development$ or intellect$ or communicat$ or expressive$ or receptive$ or learning or academic$ or arith$ or numer$ or math$ or read$ or write or writing or litera$ or spell$ or motor skill? or motor function$ or coordination or co-ordination or hyperkinetic$ or hyper-kinetic$ or clumsy child$).ti,ab.	91787

166	exp PSYCHOMOTOR PERFORMANCE/ and (tool? or scale? or index$ or scor$ or system? or test$ or questionnaire? or survey$).ti,ab.	63655

167	(assess$ adj5 (tool? or scale? or index$ or scor$ or system? or test$ or questionnaire? or survey$) adj10 (neurodevelopment$ or development$ or intellect$ or communicat$ or expressive$ or receptive$ or learning or academic$ or arith$ or numer$ or math$ or read$ or write or writing or litera$ or spell$ or motor skill? or motor function$ or coordination or co-ordination or hyperkinetic$ or hyper-kinetic$ or clumsy child$)).ti,ab.	30700

168	bayley$.ti,ab.	4016

169	(mental$ adj3 development$ adj3 index$).ti,ab.	920

170	MDI.ti,ab.	4160

171	((psychomotor or psycho-motor) adj3 development$ adj3 index$).ti,ab.	615

172	PDI.ti,ab.	8394

173	(Ages and stages questionnaire?).ti,ab.	765

174	(Strengths and Difficulties Questionnaire?).ti,ab.	2765

175	PEDIATRIC OBESITY/	13039

176	((p?ediatric? or baby or babies or infan$ or toddler? or child$ or schoolchild$ or preadolescen$ or adolescen$ or teen? or prepubescent or pubescent or offspring) adj10 (obes$ or overweight or over-weight)).ti,ab.	50635

177	(ADOLESCENT/ or MINORS/ or exp CHILD/ or exp INFANT/ or exp PEDIATRICS/) and ASTHMA/	52748

178	((p?ediatric? or baby or babies or infan$ or toddler? or child$ or schoolchild$ or preadolescen$ or adolescen$ or teen? or prepubescent or pubescent or offspring) adj10 asthma$).ti,ab.	37315

179	(ADOLESCENT/ or MINORS/ or exp CHILD/ or exp INFANT/ or exp PEDIATRICS/) and DIABETES MELLITUS, TYPE 1/	30477

180	((p?ediatric? or baby or babies or infan$ or toddler? or child$ or schoolchild$ or preadolescen$ or adolescen$ or teen? or prepubescent or pubescent or offspring) adj10 (type adj1 (one or "1") adj3 diabet$)).ti,ab.	10495

181	((p?ediatric? or baby or babies or infan$ or toddler? or child$ or schoolchild$ or preadolescen$ or adolescen$ or teen? or prepubescent or pubescent or offspring) adj10 T1D).ti,ab.	2327

182	exp AUTISM SPECTRUM DISORDER/	40333

183	(Asperger? or autis$ or Kanner?).ti,ab.	62240

184	ASD.ti,ab.	31813

185	or/66-184	2033391

186	DECISION MAKING/	103666

187	DECISION SUPPORT TECHNIQUES/	22351

188	decision?.ti,ab.	471243

189	186 or 187 or 188	521058

190	exp CESAREAN SECTION/ and (LABOR, INDUCED/ or CERVICAL RIPENING/ or exp EXTRACTION, OBSTETRICAL/ or OBSTETRICAL FORCEPS/ or NATURAL CHILDBIRTH/ or VAGINAL BIRTH AFTER CESAREAN/) and (MOTHERS/ or ADOLESCENT/ or MINORS/ or exp CHILD/ or exp INFANT/ or exp PEDIATRICS/) and (RISK/ or RISK FACTORS/)	437

191	DELIVERY, OBSTETRIC/mt and (MOTHERS/ or ADOLESCENT/ or MINORS/ or exp CHILD/ or exp INFANT/ or exp PEDIATRICS/) and (RISK/ or RISK FACTORS/)	678

192	((c?esar#an$ or c section$ or csection$ or (deliver$ adj3 abdom$)) adj5 ((induc$ adj3 (labo?r$ or birth$ or born or deliver$)) or (cervi$ adj3 ripen$) or ((extract$ or vacuum$) adj3 (birth$ or born or deliver$ or obstetric$)) or (vacuum$ adj3 extract$) or ventouse? or forcep? or (instrument$ adj3 deliver$) or ((natural$ or unassisted or un-assisted) adj3 (birth$ or born or deliver$)) or (spontaneous$ adj3 (birth$ or born or deliver$)) or ((vagina$ or cephalic$) adj1 (birth$ or born or deliver$)) or VBAC) adj5 (maternal$ or mother$ or wom?n? or neonat$ or baby or babies or infant? or preschool$ or pre-school$ or child$ or adolescen$ or teenage$) adj5 risk?).ti,ab.	177

193	(mode? adj3 (birth? or deliver$) adj5 (maternal$ or mother$ or wom?n? or neonat$ or baby or babies or infant? or preschool$ or pre-school$ or child$ or adolescen$ or teenage$) adj5 risk?).ti,ab.	133

194	190 or 191 or 192 or 193	1359

195	((c?esar#an$ or c section$ or csection$ or (deliver$ adj3 abdom$)) adj5 (subsequent$ or prior)).ti,ab.	2471

196	(mode? adj3 (birth? or deliver$) adj5 (subsequent$ or prior)).ti,ab.	219

197	195 or 196	2621

198	exp *CESAREAN SECTION/ and *POSTOPERATIVE COMPLICATIONS/	1051

199	exp *CESAREAN SECTION/ae [Adverse Effects]	4045

200	exp *CESAREAN SECTION/co [Complications]	253

201	44 and 62 and 185	8336

202	65 and 185	5795

203	44 and 62 and 189	1291

204	65 and 189	1331

205	194 or 197 or 198 or 199 or 200 or 201 or 202 or 203 or 204	20723

206	limit 205 to english language	18101

207	LETTER/	1201999

208	EDITORIAL/	630170

209	NEWS/	215966

210	exp HISTORICAL ARTICLE/	409173

211	ANECDOTES AS TOPIC/	4747

212	COMMENT/	989937

213	CASE REPORT/	2307968

214	(letter or comment*).ti.	183273

215	or/207-214	4847192

216	RANDOMIZED CONTROLLED TRIAL/ or random*.ti,ab.	1504034

217	215 not 216	4815858

218	ANIMALS/ not HUMANS/	5041664

219	exp ANIMALS, LABORATORY/	946034

220	exp ANIMAL EXPERIMENTATION/	10266

221	exp MODELS, ANIMAL/	635526

222	exp RODENTIA/	3502059

223	(rat or rats or mouse or mice).ti.	1420352

224	or/217-223	10739827

225	206 not 224	15567

226	10 and 225	1293

227	19 and 225	2690

228	32 and 225	7994

229	35 and 225	938

230	38 and 225	694

231	41 and 225	818

232	or/226-231	10977

233	limit 232 to yr="2020 -Current"	2483

234	(201908* or 201909* or 201910* or 201911* or 201912*).dt.	516428

235	232 and 234	286

236	233 or 235	2656

237	afghanistan/ or africa/ or africa, northern/ or africa, central/ or africa, eastern/ or "africa south of the sahara"/ or africa, southern/ or africa, western/ or albania/ or algeria/ or andorra/ or angola/ or "antigua and barbuda"/ or argentina/ or armenia/ or azerbaijan/ or bahamas/ or bahrain/ or bangladesh/ or barbados/ or belize/ or benin/ or bhutan/ or bolivia/ or borneo/ or "bosnia and herzegovina"/ or botswana/ or brazil/ or brunei/ or bulgaria/ or burkina faso/ or burundi/ or cabo verde/ or cambodia/ or cameroon/ or central african republic/ or chad/ or exp china/ or comoros/ or congo/ or cote d'ivoire/ or croatia/ or cuba/ or "democratic republic of the congo"/ or cyprus/ or djibouti/ or dominica/ or dominican republic/ or ecuador/ or egypt/ or el salvador/ or equatorial guinea/ or eritrea/ or eswatini/ or ethiopia/ or fiji/ or gabon/ or gambia/ or "georgia (republic)"/ or ghana/ or grenada/ or guatemala/ or guinea/ or guinea-bissau/ or guyana/ or haiti/ or honduras/ or independent state of samoa/ or exp india/ or indian ocean islands/ or indochina/ or indonesia/ or iran/ or iraq/ or jamaica/ or jordan/ or kazakhstan/ or kenya/ or kosovo/ or kuwait/ or kyrgyzstan/ or laos/ or lebanon/ or liechtenstein/ or lesotho/ or liberia/ or libya/ or madagascar/ or malaysia/ or malawi/ or mali/ or malta/ or mauritania/ or mauritius/ or mekong valley/ or melanesia/ or micronesia/ or monaco/ or mongolia/ or montenegro/ or morocco/ or mozambique/ or myanmar/ or namibia/ or nepal/ or nicaragua/ or niger/ or nigeria/ or oman/ or pakistan/ or palau/ or exp panama/ or papua new guinea/ or paraguay/ or peru/ or philippines/ or qatar/ or "republic of belarus"/ or "republic of north macedonia"/ or romania/ or exp russia/ or rwanda/ or "saint kitts and nevis"/ or saint lucia/ or "saint vincent and the grenadines"/ or "sao tome and principe"/ or saudi arabia/ or serbia/ or sierra leone/ or senegal/ or seychelles/ or singapore/ or somalia/ or south africa/ or south sudan/ or sri lanka/ or sudan/ or suriname/ or syria/ or taiwan/ or tajikistan/ or tanzania/ or thailand/ or timor-leste/ or togo/ or tonga/ or "trinidad and tobago"/ or tunisia/ or turkmenistan/ or uganda/ or ukraine/ or united arab emirates/ or uruguay/ or uzbekistan/ or vanuatu/ or venezuela/ or vietnam/ or west indies/ or yemen/ or zambia/ or zimbabwe/	1261938

238	"Organisation for Economic Co-Operation and Development"/	498

239	australasia/ or exp australia/ or austria/ or baltic states/ or belgium/ or exp canada/ or chile/ or colombia/ or costa rica/ or czech republic/ or exp denmark/ or estonia/ or europe/ or finland/ or exp france/ or exp germany/ or greece/ or hungary/ or iceland/ or ireland/ or israel/ or exp italy/ or exp japan/ or korea/ or latvia/ or lithuania/ or luxembourg/ or mexico/ or netherlands/ or new zealand/ or north america/ or exp norway/ or poland/ or portugal/ or exp "republic of korea"/ or "scandinavian and nordic countries"/ or slovakia/ or slovenia/ or spain/ or sweden/ or switzerland/ or turkey/ or exp united kingdom/ or exp united states/	3455083

240	European Union/	17457

241	Developed Countries/	21266

242	238 or 239 or 240 or 241	3470782

243	237 not 242	1173338

244	236 not 243	2335

**Embase** <1974 to 2022 Week 50>

1	SYSTEMATIC REVIEW/	384115

2	META-ANALYSIS/	265069

3	(meta analy* or metanaly* or metaanaly*).ti,ab.	325192

4	((systematic or evidence) adj2 (review* or overview*)).ti,ab.	374132

5	(reference list* or bibliograph* or hand search* or manual search* or relevant journals).ab.	64305

6	(search strategy or search criteria or systematic search or study selection or data extraction).ab.	92354

7	(search* adj4 literature).ab.	115314

8	(medline or pubmed or cochrane or embase or psychlit or psyclit or psychinfo or psycinfo or cinahl or science citation index or bids or cancerlit).ab.	412176

9	((pool* or combined) adj2 (data or trials or studies or results)).ab.	87490

10	cochrane.jw.	23898

11	or/1-10	891308

12	random*.ti,ab.	1867644

13	factorial*.ti,ab.	45416

14	(crossover* or cross over*).ti,ab.	121417

15	((doubl* or singl*) adj blind*).ti,ab.	264379

16	(assign* or allocat* or volunteer* or placebo*).ti,ab.	1210531

17	CROSSOVER PROCEDURE/	72352

18	SINGLE BLIND PROCEDURE/	48595

19	RANDOMIZED CONTROLLED TRIAL/	741064

20	DOUBLE BLIND PROCEDURE/	201653

21	or/12-20	2774508

22	COHORT ANALYSIS/	932795

23	cohort?.ti,ab.	1354654

24	FOLLOW UP/	1937920

25	(Follow$ up adj3 (study or studies)).ti,ab.	113413

26	LONGITUDINAL STUDY/	182938

27	longitudinal$.ti,ab.	441420

28	PROSPECTIVE STUDY/	815714

29	prospective$.ti,ab.	1298786

30	RETROSPECTIVE STUDY/	1352644

31	retrospective$.ti,ab.	1606759

32	OBSERVATIONAL STUDY/	299800

33	observational$.ti,ab.	383956

34	or/22-33	5508405

35	exp CASE CONTROL STUDY/	214715

36	case control$.ti,ab.	201166

37	35 or 36	281958

38	REGISTER/	119573

39	(registry or registries).ti,ab.	276619

40	38 or 39	311661

41	CROSS-SECTIONAL STUDY/	523653

42	cross sectional.ti,ab.	627434

43	41 or 42	751517

44	exp CESAREAN SECTION/	116835

45	(c?esar#an$ or c section$ or csection$ or (deliver$ adj3 abdom$)).ti,ab.	106276

46	44 or 45	139476

47	LABOR, INDUCTION/	14925

48	(induc$ adj3 (labo?r$ or birth$ or born or deliver$)).ti,ab.	23224

49	UTERINE CERVIX RIPENING/	3045

50	(cervi$ adj3 ripen$).ti,ab.	2967

51	VACUUM EXTRACTION/	3088

52	((extract$ or vacuum$) adj3 (birth$ or born or deliver$ or obstetric$)).ti,ab.	3567

53	(vacuum$ adj3 extract$).ti,ab.	2176

54	ventouse?.ti,ab.	510

55	FORCEPS DELIVERY/	3461

56	OBSTETRIC FORCEPS/	327

57	forcep?.ti,ab.	19857

58	(instrument$ adj3 deliver$).ti,ab.	4248

59	NATURAL CHILDBIRTH/	2434

60	((natural$ or unassisted or un-assisted) adj3 (birth$ or born or deliver$)).ti,ab.	3586

61	(spontaneous$ adj3 (birth$ or born or deliver$)).ti,ab.	13763

62	VAGINAL DELIVERY/	40608

63	VAGINAL BIRTH AFTER CESAREAN/	799

64	((vagina$ or cephalic$) adj1 (birth$ or born or deliver$)).ti,ab.	35816

65	VBAC.ti,ab.	1396

66	or/47-65	112768

67	(mode? adj3 (birth? or deliver$)).ti,ab.	29677

68	((maternal$ or mother$ or wom?n?) adj5 short$ adj5 term adj5 outcome?).ti,ab.	378

69	URINARY TRACT INJURY/	1187

70	BLADDER INJURY/	5670

71	BLADDER RUPTURE/	1283

72	(bladder? adj3 injur$).ti,ab.	4139

73	INTESTINE INJURY/	12165

74	(bowel? adj3 injur$).ti,ab.	4057

75	URETER INJURY/	4461

76	(ureter$ adj3 injur$).ti,ab.	4056

77	OBSTETRIC HEMORRHAGE/	2684

78	UTERUS BLEEDING/	10577

79	POSTPARTUM HEMORRHAGE/	17287

80	((major or moderate$ or severe$) adj5 (h?emorrhag$ or (blood adj2 (loss or lose or losing)) or bleed$)).ti,ab.	76059

81	((postpartum or post-partum) adj3 (h?emorrhag$ or (blood adj2 (loss or lose or losing)) or bleed$)).ti,ab.	13768

82	((>1000ml or >1000 ml or >1000millilit$ or >1000 millilit$) adj3 (h?emorrhag$ or (blood adj2 (loss or lose or losing)) or bleed$)).ti,ab.	820

83	MOTHER/ and PATIENT SATISFACTION/	596

84	MOTHER/ and "QUALITY OF LIFE"/	1925

85	((maternal or mother?) adj5 satisf$).ti,ab.	2718

86	"health related quality of life".ti,ab.	79490

87	HRQOL?.ti,ab.	34556

88	MATERNAL DEATH/	3961

89	MATERNAL MORTALITY/	23345

90	((maternal$ or mother?) adj5 (death? or mortalit$)).ti,ab.	38342

91	HOSPITAL ADMISSION/ and (INTENSIVE CARE UNIT/ or MEDICAL INTENSIVE CARE UNIT/ or SURGICAL INTENSIVE CARE UNIT/)	28825

92	((Intensive Therapy Unit? or ITU? or High Dependency Unit? or HDU? or Intensive care or ICU or PICU or NICU) adj5 admi$).ti,ab.	111375

93	HYSTERECTOMY/ and (peripart$ or peri-part$).ti,ab.	878

94	VAGINAL HYSTERECTOMY/ and (peripart$ or peri-part$).ti,ab.	14

95	((peripart$ or peri-part$) adj3 hysterectom$).ti,ab.	780

96	exp THROMBOSIS/	415096

97	exp THROMBOEMBOLISM/	603721

98	thrombo$.ti,ab.	603512

99	((maternal$ or mother$ or wom?n?) adj5 long$ adj5 term adj5 outcome?).ti,ab.	1181

100	PLACENTA ACCRETA/	4966

101	placenta$ accreta.ti,ab.	3448

102	(morbid$ adj3 adher$ adj3 placenta$).ti,ab.	583

103	(abnormal$ adj3 inva$ adj3 placenta$).ti,ab.	441

104	UTERUS RUPTURE/	7103

105	(uter$ adj3 ruptur$).ti,ab.	6200

106	STILLBIRTH/	21480

107	stillbirth?.ti,ab.	19596

108	SPONTANEOUS ABORTION/	48114

109	RECURRENT ABORTION/	7282

110	miscarr$.ti,ab.	29895

111	(abort$ adj3 (spontaneous$ or habitual$)).ti,ab.	17592

112	URINE INCONTINENCE/	53653

113	STRESS INCONTINENCE/	25388

114	((stress$ or mix$ or effort$ or urin$) adj3 incontinen$).ti,ab.	51861

115	FECES INCONTINENCE/	23473

116	(f?ecal$ adj3 incontinen$).ti,ab.	12979

117	POSTNATAL DEPRESSION/	6238

118	(depress$ adj5 (postnatal$ or post-natal$ or postpartum or post-partum)).ti,ab.	12836

119	PND.ti,ab.	8008

120	POSTTRAUMATIC STRESS DISORDER/	75168

121	((post-trauma$ or posttrauma$) adj3 stress$ adj3 disorder?).ti,ab.	44569

122	PTSD.ti,ab.	39860

123	((neonat$ or baby or babies or infant?) adj5 short$ adj5 term adj5 outcome?).ti,ab.	948

124	exp PERINATAL MORTALITY/	28372

125	(perinatal$ adj5 (death? or mortalit$)).ti,ab.	22988

126	((stillbirth or mortalit$) adj5 (one or "1" or two or "2" or three or "3" or four or "4" or five or "5" or six or "6" or seven or "7") adj3 day?).ti,ab.	26100

127	HOSPITAL ADMISSION/ and NEONATAL INTENSIVE CARE UNIT/	2433

128	((baby or babies or neonat$) adj5 care unit? adj5 admi$).ti,ab.	8438

129	(NICU adj5 admi$).ti,ab.	7794

130	NEONATAL RESPIRATORY DISTRESS SYNDROME/	8667

131	(respirat$ adj3 distress$ adj3 (baby or babies or neonat$)).ti,ab.	4400

132	(respirat$ adj3 morbidit$).ti,ab.	4554

133	HYPOXIC ISCHEMIC ENCEPHALOPATHY/	10128

134	(hypoxi$ adj3 ischemi$ adj3 (encephalop$ or brain? or cerebral$)).ti,ab.	9755

135	PERIPHERAL NERVE INJURY/	9792

136	BRACHIAL PLEXUS INJURY/	5558

137	PHRENIC NERVE/ and NERVE INJURY/	694

138	FACIAL NERVE INJURY/	2137

139	(nerve? adj3 (injur$ or trauma$)).ti,ab.	43552

140	(brachial plexus adj3 (injur$ or trauma$)).ti,ab.	4144

141	exp BRAIN HEMORRHAGE/	167913

142	((intracranial or brain or cerebral or subarachnoid) adj3 (h?emorrhag$ or bleed$)).ti,ab.	83036

143	(extracranial adj3 (h?emorrhag$ or bleed$)).ti,ab.	551

144	(cranial adj3 (h?emorrhag$ or bleed$)).ti,ab.	859

145	NEWBORN INFECTION/	8367

146	(infect$ adj3 morbidit$).ti,ab.	8734

147	((baby or babies or infant? or preschool$ or pre-school$ or child$ or adolescen$ or teenage$) adj5 long$ adj5 term adj5 outcome?).ti,ab.	4529

148	INFANT MORTALITY/	21421

149	((infant? or neonat$ or baby or babies) adj5 (death? or mortalit$)).ti,ab.	62562

150	CHILDHOOD MORTALITY/	14074

151	exp CHILD DEATH/	29058

152	(child$ adj5 (death? or mortalit$)).ti,ab.	44491

153	CEREBRAL PALSY/	42675

154	((cerebral or brain or central) adj3 (pals$ or paralys?s or pares?s)).ti,ab.	38380

155	DEVELOPMENTAL DISORDER/	37874

156	DEVELOPMENTAL DELAY/	14439

157	(neurodevelopment$ or neuro-development$).ti,ab.	62001

158	((development$ or intellect$ or communicat$ or expressive$ or receptive$ or learning or academic$ or arith$ or numer$ or math$ or read$ or write or writing or litera$ or spell$ or motor skill? or motor function$ or coordination or co-ordination or hyperkinetic$ or hyper-kinetic$ or clumsy child$) adj3 (disab$ or disorder? or difficult$ or impair$ or delay$)).ti,ab.	214279

159	(Asperger? or Kanner? or dyscalculi$ or acalculi$ or dyslexi$ or alexi$ or word blind$).ti,ab.	19798

160	(PDD or PDD-NOS or DCD or SDDMF).ti,ab.	13902

161	COGNITIVE DEFECT/	201516

162	(cognit$ adj3 (disab$ or disorder? or difficult$ or impair$ or delay$)).ti,ab.	173143

163	exp COMMUNICATION DISORDER/	82486

164	((speech or speak$ or language?) adj3 (disab$ or disorder? or difficult$ or impair$ or delay$)).ti,ab.	33842

165	(Dysglossi$ or cluttering? or verbal fluency disorder? or Rhinolali$ or dyslali$ or aprosodi$ or Aphasi$ or Articulation Disorder? or Dysarthri$ or Echolali$ or mute or Mutism? or Stutter$ or Agraphi$ or Anomi$ or Dyslexi$ or Alexi$).ti,ab.	64298

166	exp PSYCHOMOTOR DISORDER/	84890

167	((Psychomotor or psycho-motor) adj3 (disab$ or disorder? or difficult$ or impair$ or delay$)).ti,ab.	4761

168	(Dyspraxi$ or apraxi$).ti,ab.	7167

169	exp NEUROPSYCHOLOGICAL TEST/	134235

170	PSYCHOLOGIC TEST/ and (neurodevelopment$ or development$ or intellect$ or communicat$ or expressive$ or receptive$ or learning or academic$ or arith$ or numer$ or math$ or read$ or write or writing or litera$ or spell$ or motor skill? or motor function$ or coordination or co-ordination or hyperkinetic$ or hyper-kinetic$ or clumsy child$).ti,ab.	8798

171	PSYCHOMOTOR PERFORMANCE/ and (tool? or scale? or index$ or scor$ or system? or test$ or questionnaire? or survey$).ti,ab.	13136

172	(assess$ adj5 (tool? or scale? or index$ or scor$ or system? or test$ or questionnaire? or survey$) adj10 (neurodevelopment$ or development$ or intellect$ or communicat$ or expressive$ or receptive$ or learning or academic$ or arith$ or numer$ or math$ or read$ or write or writing or litera$ or spell$ or motor skill? or motor function$ or coordination or co-ordination or hyperkinetic$ or hyper-kinetic$ or clumsy child$)).ti,ab.	42677

173	bayley$.ti,ab.	5603

174	(mental$ adj3 development$ adj3 index$).ti,ab.	1100

175	MDI.ti,ab.	7234

176	((psychomotor or psycho-motor) adj3 development$ adj3 index$).ti,ab.	744

177	PDI.ti,ab.	10844

178	(Ages and stages questionnaire?).ti,ab.	991

179	(Strengths and Difficulties Questionnaire?).ti,ab.	3411

180	CHILDHOOD OBESITY/	19630

181	((p?ediatric? or baby or babies or infan$ or toddler? or child$ or schoolchild$ or preadolescen$ or adolescen$ or teen? or prepubescent or pubescent or offspring) adj10 (obes$ or overweight or over-weight)).ti,ab.	71962

182	(exp ADOLESCENT/ or exp CHILD/ or exp INFANT/ or exp PEDIATRICS/) and exp ASTHMA/	76138

183	((p?ediatric? or baby or babies or infan$ or toddler? or child$ or schoolchild$ or preadolescen$ or adolescen$ or teen? or prepubescent or pubescent or offspring) adj10 asthma$).ti,ab.	53486

184	(exp ADOLESCENT/ or exp CHILD/ or exp INFANT/ or exp PEDIATRICS/) and INSULIN DEPENDENT DIABETES MELLITUS/	34392

185	((p?ediatric? or baby or babies or infan$ or toddler? or child$ or schoolchild$ or preadolescen$ or adolescen$ or teen? or prepubescent or pubescent or offspring) adj10 (type adj1 (one or "1") adj3 diabet$)).ti,ab.	17723

186	((p?ediatric? or baby or babies or infan$ or toddler? or child$ or schoolchild$ or preadolescen$ or adolescen$ or teen? or prepubescent or pubescent or offspring) adj10 T1D).ti,ab.	5499

187	exp AUTISM/	88430

188	(Asperger? or autis$ or Kanner?).ti,ab.	80124

189	ASD.ti,ab.	44268

190	or/68-189	2933293

191	exp DECISION MAKING/	442451

192	DECISION SUPPORT SYSTEM/	26066

193	decision?.ti,ab.	648246

194	191 or 192 or 193	872574

195	exp CESAREAN SECTION/ and (LABOR, INDUCTION/ or UTERINE CERVIX RIPENING/ or VACUUM EXTRACTION/ or FORCEPS DELIVERY/ or OBSTETRIC FORCEPS/ or NATURAL CHILDBIRTH/ or VAGINAL DELIVERY/ or VAGINAL BIRTH AFTER CESAREAN/) and (MOTHERS/ or exp ADOLESCENT/ or exp CHILD/ or exp INFANT/ or exp PEDIATRICS/) and (RISK/ or RISK FACTOR/)	1909

196	((c?esar#an$ or c section$ or csection$ or (deliver$ adj3 abdom$)) adj5 ((induc$ adj3 (labo?r$ or birth$ or born or deliver$)) or (cervi$ adj3 ripen$) or ((extract$ or vacuum$) adj3 (birth$ or born or deliver$ or obstetric$)) or (vacuum$ adj3 extract$) or ventouse? or forcep? or (instrument$ adj3 deliver$) or ((natural$ or unassisted or un-assisted) adj3 (birth$ or born or deliver$)) or (spontaneous$ adj3 (birth$ or born or deliver$)) or ((vagina$ or cephalic$) adj1 (birth$ or born or deliver$)) or VBAC) adj5 (maternal$ or mother$ or wom?n? or neonat$ or baby or babies or infant? or preschool$ or pre-school$ or child$ or adolescen$ or teenage$) adj5 risk?).ti,ab.	274

197	(mode? adj3 (birth? or deliver$) adj5 (maternal$ or mother$ or wom?n? or neonat$ or baby or babies or infant? or preschool$ or pre-school$ or child$ or adolescen$ or teenage$) adj5 risk?).ti,ab.	203

198	195 or 196 or 197	2330

199	((c?esar#an$ or c section$ or csection$ or (deliver$ adj3 abdom$)) adj5 (subsequent$ or prior)).ti,ab.	4135

200	(mode? adj3 (birth? or deliver$) adj5 (subsequent$ or prior)).ti,ab.	363

201	199 or 200	4397

202	exp CESAREAN SECTION/ and *POSTOPERATIVE COMPLICATION/	634

203	exp CESAREAN SECTION/co [Complication]	1

204	exp CESAREAN SECTION/ and ADVERSE OUTCOME/	2366

205	46 and 66 and 190	18717

206	67 and 190	8659

207	46 and 66 and 194	2836

208	67 and 194	2034

209	198 or 201 or 202 or 203 or 204 or 205 or 206 or 207 or 208	32743

210	limit 209 to english language	30748

211	etter.pt. or LETTER/	1172742

212	note.pt.	918295

213	editorial.pt.	746050

214	CASE REPORT/ or CASE STUDY/	2883358

215	(letter or comment*).ti.	227582

216	or/211-215	5477738

217	RANDOMIZED CONTROLLED TRIAL/ or random*.ti,ab.	1978574

218	216 not 217	5422089

219	ANIMAL/ not HUMAN/	1170193

220	NONHUMAN/	7129021

221	exp ANIMAL EXPERIMENT/	2936341

222	exp EXPERIMENTAL ANIMAL/	785372

223	ANIMAL MODEL/	1615445

224	exp RODENT/	3905303

225	(rat or rats or mouse or mice).ti.	1572907

226	or/218-225	14350499

227	210 not 226	25836

228	11 and 227	2004

229	21 and 227	3835

230	34 and 227	14510

231	37 and 227	1075

232	40 and 227	1019

233	43 and 227	1386

234	or/228-233	18705

235	limit 234 to yr="2020 -Current"	5142

236	(201908* or 201909* or 201910* or 201911* or 201912*).dd,dc.	750169

237	234 and 236	450

238	235 or 237	5561

239	afghanistan/ or africa/ or africa, northern/ or africa, central/ or africa, eastern/ or "africa south of the sahara"/ or africa, southern/ or africa, western/ or albania/ or algeria/ or andorra/ or angola/ or "antigua and barbuda"/ or argentina/ or armenia/ or azerbaijan/ or bahamas/ or bahrain/ or bangladesh/ or barbados/ or belize/ or benin/ or bhutan/ or bolivia/ or borneo/ or "bosnia and herzegovina"/ or botswana/ or brazil/ or brunei/ or bulgaria/ or burkina faso/ or burundi/ or cabo verde/ or cambodia/ or cameroon/ or central african republic/ or chad/ or exp china/ or comoros/ or congo/ or cote d'ivoire/ or croatia/ or cuba/ or "democratic republic of the congo"/ or cyprus/ or djibouti/ or dominica/ or dominican republic/ or ecuador/ or egypt/ or el salvador/ or equatorial guinea/ or eritrea/ or eswatini/ or ethiopia/ or fiji/ or gabon/ or gambia/ or "georgia (republic)"/ or ghana/ or grenada/ or guatemala/ or guinea/ or guinea-bissau/ or guyana/ or haiti/ or honduras/ or independent state of samoa/ or exp india/ or indian ocean islands/ or indochina/ or indonesia/ or iran/ or iraq/ or jamaica/ or jordan/ or kazakhstan/ or kenya/ or kosovo/ or kuwait/ or kyrgyzstan/ or laos/ or lebanon/ or liechtenstein/ or lesotho/ or liberia/ or libya/ or madagascar/ or malaysia/ or malawi/ or mali/ or malta/ or mauritania/ or mauritius/ or mekong valley/ or melanesia/ or micronesia/ or monaco/ or mongolia/ or montenegro/ or morocco/ or mozambique/ or myanmar/ or namibia/ or nepal/ or nicaragua/ or niger/ or nigeria/ or oman/ or pakistan/ or palau/ or exp panama/ or papua new guinea/ or paraguay/ or peru/ or philippines/ or qatar/ or "republic of belarus"/ or "republic of north macedonia"/ or romania/ or exp russia/ or rwanda/ or "saint kitts and nevis"/ or saint lucia/ or "saint vincent and the grenadines"/ or "sao tome and principe"/ or saudi arabia/ or serbia/ or sierra leone/ or senegal/ or seychelles/ or singapore/ or somalia/ or south africa/ or south sudan/ or sri lanka/ or sudan/ or suriname/ or syria/ or taiwan/ or tajikistan/ or tanzania/ or thailand/ or timor-leste/ or togo/ or tonga/ or "trinidad and tobago"/ or tunisia/ or turkmenistan/ or uganda/ or ukraine/ or united arab emirates/ or uruguay/ or uzbekistan/ or vanuatu/ or venezuela/ or vietnam/ or west indies/ or yemen/ or zambia/ or zimbabwe/	1595812

240	"Organisation for Economic Co-Operation and Development"/	2282

241	australasia/ or exp australia/ or austria/ or baltic states/ or belgium/ or exp canada/ or chile/ or colombia/ or costa rica/ or czech republic/ or exp denmark/ or estonia/ or europe/ or finland/ or exp france/ or exp germany/ or greece/ or hungary/ or iceland/ or ireland/ or israel/ or exp italy/ or exp japan/ or korea/ or latvia/ or lithuania/ or luxembourg/ or mexico/ or netherlands/ or new zealand/ or north america/ or exp norway/ or poland/ or portugal/ or exp "republic of korea"/ or "scandinavian and nordic countries"/ or slovakia/ or slovenia/ or spain/ or sweden/ or switzerland/ or turkey/ or exp united kingdom/ or exp united states/	3620643

242	European Union/	30166

243	Developed Countries/	33630

244	240 or 241 or 242 or 243	3651976

245	239 not 244	1453313

246	238 not 245	4868

247	conference abstract.pt.	4623239

248	246 not 247	3698

**Cochrane Library**

#1	MeSH descriptor: [Cesarean Section] explode all trees

#2	(cesarean* or caesarean* or "c section*" or csection* or (deliver* near/3 abdom*)):ti,ab

#3	#1 or #2

#4	MeSH descriptor: [Labor, Induced] this term only

#5	(induc* near/3 (labor* or labour* or birth* or born or deliver*)):ti,ab

#6	MeSH descriptor: [Cervical Ripening] this term only

#7	(cervi* near/3 ripen*):ti,ab

#8	MeSH descriptor: [Extraction, Obstetrical] explode all trees

#9	((extract* or vacuum*) near/3 (birth* or born or deliver* or obstetric*)):ti,ab

#10	(vacuum* near/3 extract*):ti,ab

#11	ventouse*:ti,ab

#12	MeSH descriptor: [Obstetrical Forceps] this term only

#13	forcep*:ti,ab

#14	(instrument* near/3 deliver*):ti,ab

#15	MeSH descriptor: [Natural Childbirth] this term only

#16	((natural* or unassisted or un-assisted) near/3 (birth* or born or deliver*)):ti,ab

#17	(spontaneous* near/3 (birth* or born or deliver*)):ti,ab

#18	MeSH descriptor: [Vaginal Birth after Cesarean] this term only

#19	((vagina* or cephalic*) near/1 (birth* or born or deliver*)):ti,ab

#20	VBAC:ti,ab

#21	#4 or #5 or #6 or #7 or #8 or #9 or #10 or #11 or #12 or #13 or #14 or #15 or #16 or #17 or #18 or #19 or #20

#22	#3 and #21

#23	MeSH descriptor: [Delivery, Obstetric] this term only and with qualifier(s): [methods - MT]

#24	(mode* near/3 (birth* or deliver*)):ti,ab

#25	#22 or #23 or #24

Date limits:

Publication date CDSR 2/8/2019 – 31/12/2022

Date added to CENTRAL 2/8/2019 – 31/12/2022

International **HTA database** (<https://database.inahta.org/>)

(CESAREAN SECTION/)[mh] OR (NATURAL CHILDBIRTH/)[mh] OR (VAGINAL BIRTH AFTER CESAREAN/)[mh] OR (DELIVERY, OBSTETRIC/)[mh]
