## Supplementary file S2 for "Outcomes of planned caesarean birth compared with planned or actual vaginal birth: an update and expansion of the NICE Caesarean Birth Guideline systematic review NG192"

**Supplementary File 2 - Summary study characteristics of the included studies**

*Table S1: Clinical evidence tables for benefits and risks of caesarean birth compared with planned vaginal birth – short-term outcomes*

*Table S2: Clinical evidence tables for benefits and risks of caesarean birth compared with planned vaginal birth – long-term outcomes*

### Table S1. Clinical evidence tables for benefits and risks of caesarean birth compared with planned vaginal birth – short term outcomes

| **Study details** | **Participants** | **Interventions** | **Methods** | **Outcomes** | **Quality assessment** |
| --- | --- | --- | --- | --- | --- |
| **Full citation**  Dahlquist Karin, Stuart Andrea, Källén Karin. Planned caesarean section vs planned vaginal delivery among women without formal medical indication for planned caesarean section: A retrospective cohort study of maternal short-term complications. Acta Obstet Gynecol Scand. 2022;101:1026-1032.  **Ref Id**  Dahlquist 2022  **Country/ies where the study was carried out**  Sweden  **Study type**  Retrospective cohort study  **Aim of the study**  To investigate the rates of short-term complications (infections and thromboembolism within 6 weeks postpartum) with planned caesarean compared with those with planned vaginal delivery in a group with no formal medical indication for planned caesarean.  **Study dates**  2008-2017  **Source of funding**  Stig and Ragna Gorthons fundation | **Sample Size** N= 714,326 (n= 22855 planned caesarean and n= 691471 planned vaginal birth within planned vaginal birth n= 613,335 non-instrumental birth) **Characteristics** \| Planned caesarean (n= 22,855) (%) \| Planned vaginal non- instrumental delivery (n= 612,072) (%); Maternal age (years) <20 20-39 35-39 ≥40 \| 0.6 63.8 27.5 8.0 \| 1.5 79.0 16.2 3.2; Maternal BMI (kg/m2) <18.5 18.5–24.9 25–29.9 30–34.9 ≥35 Not known \| 2.5 56.7 23.8 7.7 3.1 6.3 \| 2.5 58.4 22.9 7.5 2.8 6.0; Parity 1 para 2 para 3+ para \| 32.7 57.3 10.1 \| 44.3 37.8 17.9 **Inclusion criteria** - Low risk cephalic, singleton and term births without formal medical indication for planned CS **Exclusion criteria** - Patients with previous caesarean or medical indication for planned caesarean and high-risk diagnoses/conditions for maternal/foetal morbidity: multiple gestation, non-cephalic presentation, preterm birth (gestational age <37 weeks or >42 weeks), and macrosomia (birth weight >4500g). - Patients with breech deliveries, multiple births, diabetes, gestational diabetes, ablatio placentae, placenta accrete, and pre-eclampsia. | **Interventions**  Planned caesarean birth versus planned vaginal birth | **Details**  Merged data from three Swedish national registries: medical birth register, Swedish national patient register, and Swedish prescribed drug registry.  Results reported as RR using modified Poisson regression models adjusting for maternal age, parity, body mass index, smoking, country of birth, and county | **Included in the decision aid (main paper)**  ***Maternal***  Postpartum infection (any)  Wound infection  Mastitis  Endometritis  Urinary tract infection  Thromboembolic disease  Septicaemia  ***Infant***  None  **Supplementary outcomes**  ***Maternal***  Antibiotic use  ***Infant***  None | **Limitations**  Methodological limitations assessed using the Newcastle Ottawa quality assessment form for cohort studies  **Overall quality:** good |
| **Full citation**  Guo Yanfang, Murphy S.Q Malia, Erwin Erica, Fakhraei Romina, Corsi J Daniel, White Rennicks Ruth, Harvey L.J Alysha, Gaudet M Laura, Walker C. Mark, Wen Wu Shi, El-Chaâr Darine., Birth outcomes following cesarean birth on maternal request: a population-based cohort study, CMAJ 2021 May 3;193:E633-44.  **Ref Id**  Guo 2021  **Country/ies where the study was carried out**  Canada  **Study type**  Population-based retrospective cohort study  **Aim of the study**  To evaluate the trends, determinants and outcomes of Caesarean birth on maternal request (CDMR) using an intent-to-treat approach.  **Study dates**  April 2012- March 2018  **Source of funding**  This study was supported by a Canadian Institutes of Health Research Foundation Grant (FDN 148438) | **Sample Size** N=422,210 (n= 1827 in planned caesarean on maternal demand group and n=420,383 in the planned vaginal birth group) **Characteristics** \| Planned CDMR \| Planned vaginal birth; Maternal age, year, mean ± SD \| 32.5 ± 5.7 \| 29.7 ± 5.2; Pre-pregnancy BMI, kg/m2 ,median (IQR) \| 23.2 (20.7–27.0) \| 23.4 (20.9–27.2); Infant birth weight, grams, mean ± SD \| 3430.2 ± 427.1 \| 3462.5 ± 448.4; Gestational age, week, median (IQR) \| 39.0 (38.7–39.4) \| 39.9 (39.1–40.7); Maternal drug use during pregnancy \| 31 (1.7) \| 8570 (2.0) **Inclusion criteria** - Low risk pregnancies that resulted in birth of a full-term (between ≥ 37 weeks’ and ≤ 42 weeks’ gestation) cephalic and singleton live birth - Intrapartum stillbirth weighing ≥ 500 g **Exclusion criteria** - Pregnancies with medical or pre-labour indications for caesarean birth, as well as records with missing data or data quality issues - Mothers with a caesarean birth in a previous pregnancy - Those who did not meet Health Quality Ontario definition for low-risk pregnancy - Home or birth centre birth | **Interventions**  Planned caesarean birth on maternal request versus planned vaginal birth | **Details**  Data was obtained from Better Outcomes Registry & Network (BORN) Ontario, Linked records with maternal obstetrical discharge abstracts and stillbirth/newborn abstracts from the Canadian Institute for Health Information Discharge Abstract Database.  Results were reported as RR adjusted for confounders: gestational age, maternal age, pre-pregnancy BMI, neighbourhood education quintile, maternal race, parity, conception type, anxiety, maternal substance use during pregnancy | **Included in the decision aid (main paper)**  ***Maternal***  Admission to neonatal unit  Blood transfusion  ***Infant/neonatal***  None  **Supplementary outcomes**  ***Maternal***  Unanticipated operative procedures  Risk of any Adverse Outcome Index (AOI) outcome  Any maternal component  ***Infant/neonatal***  Apgar score  Any neonatal component of Adverse Outcome Index (AOI) | Assessed using the Newcastle Ottawa quality assessment form for cohort studies  **Overall quality:** good |
| **Full citation**  Guo Y, Murphy MS, Dimanlig-Cruz S, Leclerc A, Smith MA, Corsi DJ, White RR, Dingwall-Harvey AL, Harrold J, Walker MC, Wen SW. Infant Infections Following Cesarean Birth on Maternal Request: A Population-Based Cohort Study. Journal of Obstetrics and Gynaecology Canada. 2024 Jun 1;46(6):102455. <https://doi.org/10.1016/j.jogc.2024.102455>  **Ref Id**  Guo 2024  **Country/ies where the study was carried out**  Canada  **Study type**  Population-based retrospective cohort study  **Aim of the study**  To determine the association between caesarean birth on maternal request (CDMR) and infant infection using an intent-to-treat approach  **Study dates**  1^st^ April 2012 to 31^st^ March 2018  **Source of funding**  supported by a Canadian Institutes of Health Research Foundation Grant (FDN 148438) | **Sample Size**  N=422,134 (of which n=1827 were in the planned caesarean on maternal demand group and n=420,307 were in the planned vaginal birth group)    **Characteristics**  See Guo 2021 for details.  **Inclusion criteria**  Not reported  **Exclusion criteria**  Not reported | **Interventions**  Planned caesarean birth on maternal request versus planned vaginal birth | **Details**  Secondary publication of Guo 2021.  The cohort included low-risk singleton pregnancies resulting in a term, cephalic, live birth  from the Better Outcomes Registry  and Network Ontario  Results were reported as RR adjusted for gestational age at birth, maternal age, pre-pregnancy BMI, neighbourhood education quintile, race, parity, conception type, anxiety, maternal substance use during pregnancy (includes self-reported alcohol, smoking and drug use during pregnancy), antenatal health care provider and maternal hospital level of care | **Included in the decision aid (main paper)**  ***Maternal***  None  ***Infant/neonatal***  Infant infections  Infant respiratory tract infections  Infant gastrointestinal infections  Infant otitis media  **Supplementary outcomes**  None | Assessed using the Newcastle Ottawa quality assessment form for cohort studies  **Overall quality:** good |

BMI, body mass index; CB, caesarean birth; g, gram; OR, odds ratio; RR, relative risk; SD, standard deviation; VB, vaginal birth

### Table S2. Clinical evidence tables for benefits and risks of caesarean birth compared with planned vaginal birth - long-term outcomes

| **Study details** | **Participants** | **Interventions** | **Methods** | **Outcomes** | **Quality assessment** |
| --- | --- | --- | --- | --- | --- |
| **Full citation**  Andersen, Vibeke & Möller, Sören & Jensen, Peter & Trier Møller, Frederik & Green, Anders. (2020). Caesarean Birth and Risk of Chronic Inflammatory Diseases (Inflammatory Bowel Disease, Rheumatoid Arthritis, Coeliac Disease, and Diabetes Mellitus): A Population Based Registry Study of 2,699,479 Births in Denmark During 1973–2016. Clinical Epidemiology. Volume 12. 287-293. 10.2147/CLEP.S229056.  **Ref Id**  Andersen 2020  **Country/ies where the study was carried out**  Denmark  **Study type**  Population-based registry study  **Aim of the study**  To evaluate the risk of developing CIDs (including four common hospital-diagnosed childhood CIDs) after being born by caesarean birth, taking advantage of the National Danish registries completeness, size, and long follow-up  **Study dates**  1973-2016 (Limited the analysis to vaginal birth and caesarean)  1991-2016 (comparison of vaginal birth, acute caesarean and planned caesarean)  **Source of funding**  The Danish Rheumatism Association (Gigtforeningen) (R104-A2195-B760). | **Sample Size** N= 2,672,708 (n= 122,419 planned caesareans; n= 163 893 acute caesarean and n= 1,312,522 vaginal birth) **Characteristics** Children live-born from January 1977 through March 2016 \| Caesarean birth \| Vaginal birth; Diabetes mellitus, n (%) \| 2,932 (12.3) \| 20,936 (87.7); IBD (Crohn’s disease + ulcerative colitis), n (%) \| 2,240 (11.5) \| 17,293 (88.5); Crohn’s disease, n (%) \| 1,104 (12.1) \| 8,051 (87.9); Rheumatoid arthritis, n (%) \| 1,395 (13.3) \| 9,075 (86.7); Coeliac disease, n (%) \| 1,047 (14.7) \| 6,085 (85.3); Ulcerative colitis, n (%) \| 1,428 (11.1) \| 11,436 (88.9) **Inclusion criteria** - Children born in Denmark between January 1973 and March 2016, identified through the Danish Medical Birth Register (MBR) and the Danish National Patient Registry (NPR) **Exclusion criteria** - Children with missing birth dates (N=1929) and incomplete parental information in the Danish Civil Registration System (CRS). - Births with an unknown type of birth | **Interventions**  Planned caesarean birth versus vaginal birth | **Details**  Data obtained from Danish Medical Birth Register (MBR) and the Danish National Patient Registry (NPR).  Results were reported as adjusted HR and adjusted for confounders including the decade of birth, child’s sex, parental age at birth, gender of child and mother’s and father’s diabetes mellitus, rheumatoid arthritis, coeliac disease, and inflammatory bowel disease. | **Included in the decision aid (main paper)**  ***Maternal***  None  ***Infant/neonatal***  Type 1 diabetes  **Supplementary outcomes**  ***Maternal***  None  ***Infant/neonatal***  Inflammatory bowel disease  Rheumatoid arthritis  Coeliac disease | Assessed using the Newcastle Ottawa quality assessment form for cohort studies  **Overall quality:** good |
| **Full citation**  Baud, David & Sichitiu, Joanna & Lombardi, Valeria & Rham, Maud & Meyer, Sylvain & Vial, Yvan & Achtari, Chahin. (2020). Comparison of pelvic floor dysfunction 6 years after uncomplicated vaginal versus planned caesarean deliveries: a cross-birthal study. Scientific Reports. 10. 10.1038/s41598-020-78625-3.  **Ref Id**  Baud 2020  **Country/ies where the study was carried out**  Switzerland  **Study type**  Cross-birthal study  **Aim of the study**  To evaluate faecal, urinary and sexual symptoms 6 years postpartum, comparing uncomplicated vaginal birth and planned caesarean birth, and to assess their impact on quality of life  **Study dates**  1996 and 2011  **Source of funding**  Foundation for Research and Development in Gynecology and Obstetrics of Lausanne, Switzerland; Fondation Leenaards” through the “Bourse pour la relève académique”, and by the Devisa Foundation, Switzerland. | **Sample Size** N= 604 (n=208 planned caesarean and n= 309 uncomplicated vaginal birth group returned postal questionnaires) **Characteristics** \| Planned caesarean birth \| Uncomplicated vaginal birth; Maternal age, years mean (SD) \| 37.3 (5.4) \| 36.6 (5.3); Multiparous, % \| 18.5 \| 16.3; Maternal weight, kg, mean (SD) \| 66 (15) \| 64 (12); Maternal smoker (Yes), % \| 15.6 \| 12.7; Health insurance (non-private), % \| 97.2 \| 98.1; Christian, % \| 74.4 \| 82.1 **Inclusion criteria** - Women who delivered 6 years before this study (were chosen randomly from the hospital database). - Women who gave birth either by an planned caesarean birth or a spontaneous vaginal birth with a grade 1 vaginal tear at most **Exclusion criteria** - Women under 18 years - Multiple pregnancies - Instrumental deliveries - Multiparous patients who delivered by both modes of birth | **Interventions**  Planned caesarean birth versus uncomplicated vaginal birth | **Details**  Data was obtained from the obstetrical database at the Maternity Hospital of the Centre Hospitalier Universitaire Vaudois.  Results were reported as RR adjusted for age, ethnicity, parity, weight, smoking, level of education, marital status, religion, type of health insurance. | **Included in the decision aid (main paper)**  ***Maternal***  Pain during or following vaginal penetration  ***Infant/neonatal***  None  **Supplementary outcomes**  ***Maternal***  Urgency urinary incontinence  Stress incontinence  Lower abdominal or genital pain  Wexner incontinence for gas score  Wexner incontinence for liquid stool score  Wexner incontinence for solid stool score  Wexner alteration of lifestyle score  Wexner alteration of sexual life score  Wexner need to wear a pad score  Wexner taking constipating medicine score  Wexner inability to defer defecation for 15 minutes score  ***Infant/neonatal***  None | Assessed using the Newcastle Ottawa quality assessment form for cohort studies  **Overall quality:** good |
| **Full citation**  Bodunde EO, Buckley D, O'Neill E, Al Khalaf S, Maher GM, O'Connor K, McCarthy FP, Kublickiene K, Matvienko‐Sikar K, Khashan AS. Pregnancy and birth complications and long‐term maternal mental health outcomes: A systematic review and meta‐analysis. BJOG: An International Journal of Obstetrics & Gynaecology. 2025 Jan;132(2):131-42. https://doi.org/10.1111/1471-0528.17889  **Ref Id**  Bodunde 2024  **Country/ies where the study was carried out**  Europe (k=14), North America (k=10), Asia (k=4), Australia (k=4), South America (k=1)  **Study type**  Systematic review and meta- analysis  **Aim of the study**  To review the published literature on pregnancy and birth complications and long-term maternal mental health outcomes  **Study dates**  Study published online June 2024. Included studies published between 1996 and 2022  **Source of funding**  Health Research Board (SPHeRE-2018-1) | **Sample Size**  K = 33 studies  Depression: k=1 study; anxiety disorder: k=1 study; PTSD: k=2 studies  **Characteristics**  See review for details  **Inclusion criteria**   - Peer-reviewed observational studies - Women who had at least one pregnancy with any of the following: pregnancy and birth complications as exposures of interest: pre-eclampsia; pregnancy loss (miscarriage or termination of pregnancy); stillbirth; CS (planned and/or unplanned); preterm birth (defined as birth <37 weeks of gestation); third-or fourth-degree perineal laceration; neonatal intensive care unit (NICU) admission for >72 h; major obstetric haemorrhage; and birth injury/trauma. - The comparator was no corresponding pregnancy and/or birth complication(s). - The primary outcome was depression, anxiety disorder, post-traumatic stress disorder (PTSD), substance use disorder, psychosis, schizophrenia or bipolar disorder diagnosed after the first 12 months postpartum. - The outcome of interest was adverse maternal mental health outcomes after the first year following birth.   **Exclusion criteria**   - case reports, editorials, conference abstracts and studies focused on women with a pre-existing mental illness or with mental illness prior to 1 year postpartum. | **Interventions**  Caesarean birth versus vaginal birth. Henderson (2021) and Chen (2017 do not make a distinction between planned and unplanned/medically indicated cesarean births. Hernandez-Martinez (2020) does not specify which type(s) of caesarean birth were considered. | **Details**  Systematic search of Cumulative Index to Nursing and Allied Health Literature (CINAHL), Excerpta Medica Database (Embase), PsycInfoR, PubMedR and Web of Science from inception until August 2022.  Results were reported as OR adjusted following author's definition in individual studies; these studies adjusted for factors including maternal age, smoking, body mass index, pre-existing mental health disorders, income, educational level, parity and other comorbidities. | **Included in the decision aid (main paper)**  ***Maternal***  Depression  Anxiety disorder  PTSD  ***Infant/neonatal***  **Supplementary outcomes**  None | Assessed with the ROBIS checklist  **Risk of bias in the review:** |
| **Full citation**  Cattani, Laura & Neefs, Liesbeth & Verbakel, Jan & Bosteels, Jan & Deprest, Jan. (2021). Obstetric risk factors for anorectal dysfunction after birth: a systematic review and meta-analysis. International Urogynecology Journal. 32. 10.1007/s00192-021-04723-z.  **Ref Id**  Cattani 2021  **Country/ies where the study was carried out**  UK, USA, France, Ireland, Slovenia, Norway, Switzerland, Denmark, Sweden, Czech Republic, Spain, Finland, Italy, China, Australia, Netherlands and Hong Kong  **Study type**  Systematic review and meta- analysis  **Aim of the study**  To assess the effect of obstetric events on anal incontinence and constipation after birth.  **Study dates**  Study published 11 November 2019. Included studies published between 1998 and 2019  **Source of funding**  Not reported | **Sample Size**  K= 47 studies  Anal incontinence: k=8 studies; flatus incontinence: k=4 studies  **Characteristics**  See review for details  **Inclusion criteria**   - Randomised clinical trials, controlled clinical trials, cohort studies and case–control studies, comparing potential obstetric risk factors for developing anorectal dysfunction relating to childbirth. - Studies written in English - Studies that were sufficiently comparable for statistical pooling   **Exclusion criteria**   - Duplicate records, reviews, case reports, incomplete reports, book chapters, conference abstracts, letters to the editor and comments - Cross-birthal studies | **Interventions**  Caesarean birth (any type, including planned and unplanned) versus vaginal birth | **Details**  Searched were conducted in MEDLINE, Embase and CENTRAL from inception until November 2019  Reported the effect measures of all studies as ORs and all included studies controlled for at least one confounding factor: maternal age, BMI and parity.  Included 5 studies from low/middle income countries (i.e., China) | **Included in the decision aid (main paper)**  ***Maternal***  None  ***Infant/neonatal***  None  **Supplementary outcomes**  Anal incontinence  Flatus incontinence | Systematic review assessed with the ROBIS checklist  **Risk of bias in the review:** Low |
| **Full citation**  Chavarro, Jorge & Martín-Calvo, Nerea & Yuan, Changzheng & Arvizu, Mariel & Rich-Edwards, Janet & Michels, Karin & Sun, Qi. (2020). Association of Birth by Cesarean Birth With Obesity and Type 2 Diabetes Among Adult Women. JAMA Network Open. 3. e202605. 10.1001/jamanetworkopen.2020.2605.  **Ref Id**  Chavarro 2020  **Country/ies where the study was carried out**  USA  **Study type**  Prospective cohort study  **Aim of the study**  To evaluate the association of birth by caesarean birth with offspring’s risks of obesity and type 2 diabetes in adulthood.  **Study dates**  Statistical analysis was performed from June 2017 to December 2019.  **Source of funding**  Grants R01-HD093761, U01-HL145386, UM1-CA176726, R01-DK112940, and P30-DK046200 from the National Institutes of Health. | **Sample Size** N= 33226 (n= 1089 caesarean birth caesarean no differentiation over planned and actual mode of birth and n= 32137 vaginal birth) **Characteristics** \| Caesarean birth \| Vaginal birth; Maternal age at birth, years, mean (SD) \| 28.2 (5.6) \| 26.2 (4.9); Pre-pregnancy BMI, mean (SD) \| 21.7 (3.0) \| 21.2 (2.5); Height, cm, mean (SD) \| 160.8 (6.4) \| 163.6 (6.1); Gestational diabetes, n (%) \| 7 (0.6) \| 133 (0.4); Gestational weight gain, n (%) <9.1 kg ≥9.1 kg Missing \| 394 (36.2) 611 (56.1) 84 (7.7) \| 10 526 (32.8) 19 088 (59.4) 2523 (7.9); Offspring gestational age at birth, n (%) <37 wk 37-39 wk 40-42 wk ≥43 wk \| 64 (9.2) 456 (65.7) 109 (15.7) 65 (9.4) \| 757 (4.1) 8498 (46.1) 7111 (38.6) 2071 (11.3) **Inclusion criteria** - Singleton pregnancy - Women participating in the Nurses’ Health Study II who were born between 1946 and 1964 **Exclusion criteria** - Participants who did not provide information on height or weight - Not born of a singleton pregnancy Those whose mothers did not provide information on birth mode | **Interventions**  Caesarean birth versus vaginal birth | **Details**  Included women participating in the Nurses’ Health Study II who were born between 1946 and 1964, with follow-up through the end of the 2013-2015 follow-up cycle.  Results were reported as RR and HR adjusted for maternal age at birth, race/ethnicity, maternal educational level, maternal pre-pregnancy BMI group, gestational weight gain, maternal height, gestational diabetes, preeclampsia, pregnancy-induced hypertension, year of birth, gestational age at birth, birth weight group, smoking during pregnancy, and region of residence at birth. | **Included in the decision aid (main paper)**  **None**  **Supplementary outcomes**  ***Maternal***  None  ***Infant/neonatal***  Adulthood type 2 diabetes  Adulthood obesity | Assessed using the Newcastle Ottawa quality assessment form for cohort studies  **Overall quality:** good |
| **Full citation**  Chen, M., Lin, Y., Yu, C.*et al.*Effect of cesarean birth on the risk of autism spectrum disorders/attention deficit hyperactivity disorder in offspring: a meta-analysis.*Arch Gynecol Obstet* **309**, 439–455 (2024). <https://doi.org/10.1007/s00404-023-07059-9>  **Ref Id**  Chen 2024  **Country/ies where the study was carried out**  The review was conducted in China. The included studies were conducted in Asia-China (*n* = 6), Israel (*n* = 1), Malaysia (*n* = 1), Kingdom of Saudi Arabia (n = 1), Iraq (*n* = 1), Iran (*n* = 1); Europe-Sweden (*n* = 5), Denmark (*n* = 3), Finland (*n* = 2), Spain (*n* = 1), United Kingdom (*n* = 1), Norway (*n* = 1); North America-USA (*n* = 4), Canada (*n* = 2); South America-Brazil (*n* = 1); Oceania-Australia (*n* = 2) and multicenter (*n* = 2)  **Study type**  Systematic review with meta-analysis  **Aim of the study**  To investigate the relationship between caesarean birth offspring and autism spectrum disorders (ASD)/attention deficit hyperactivity disorder (ADHD).  **Study dates**  The literature searches were conducted from inception to the end of August 2022.  **Source of funding**  The authors did not receive support from any organisation for the work. | **Sample Size**  K= 35 ( Autism spectrum condition: k=9 studies  **Characteristics**  See review for details  **Inclusion criteria**   - Cohort studies and case–control studies published in English - The relationship between the mode of obstetric birth (CB and VB) and ASD/ADHD in the offspring was assessed - Participants had a childbirth record - Patients with ASD/ADHD were diagnosed using a clear assessment tool   **Exclusion criteria**   - The article had no extractable relevant data - Only the study protocol was available, or the full text was not available - VB was not used as a control or only the relationship between VB and ASD/ADHD was explored - The outcomes involved in the study were not limited to ASD/ADHD, but were combined outcomes of these two or more neurodevelopmental disorders - Studies used the same group of participants | **Interventions**  Planned caesarean birth versus vaginal birth. | **Details**  Systematic seach of PubMed, Web of Science, Embase, and the Cochrane Library from database establishment to the end of August 2022 for studies on the association between CS  and ASD/ADHD.  Results reported as OR. The review authors report that most included studies adjusted for confounding factors, such as sex, birth weight, gestational age, maternal age, and Apgar score | **Included in the decision aid (main paper)**  ***Maternal***  None  ***Infant/neonatal***  Autism spectrum condition  **Supplementary outcomes**  None | Systematic review assessed with the ROBIS checklist  **Risk of bias in the review:** Low |
| **Full citation**  Chen, Ginden & Chiang, Wan‐Lin & Chiang, Tung‐liang. (2021). Does Cesarean Birth Increase the Occurrence of Neurodevelopmental Disorders in Childhood?. International Journal of Gynecology & Obstetrics. 158. 10.1002/ijgo.14055.  **Ref Id**  Chen 2022  **Country/ies where the study was carried out**  Taiwan  **Study type**  Prospective longitudinal cohort study  **Aim of the study**  This study aimed to investigate whether caesarean birth (CD) is associated with the occurrence of neurodevelopmental disorders (NDDs) at the age of 8 years.  **Study dates**  Not reported (Used children born in 2005 from the National Birth Report Database)  **Source of funding**  Health Promotion Administration, Department of Health and Welfare in and the Ministry of Science and Technology. | **Sample Size** N= 19142 (n= 3649 planned caesarean birth; n= 2467 unplanned caesarean and n= 13026 vaginal birth) **Characteristics** Infant characteristics at age 8 years \| Planned caesarean birth \| Vaginal birth; Neurodevelopmental disorders, n (%) \| 178 (4.8) \| 648 (5.0); Learning disabilities, n (%) \| 74 (2.0) \| 261 (2.0); Developmental delay, n (%) \| 72 (2.0) \| 260 (2.0); ADHD, n (%) \| 77 (2.1) \| 306 (2.4); Sensory integration disorder, n (%) \| 40 (1.1) \| 124 (1.0); Autism, n (%) \| 21 (0.6) \| 66 (0.5); Gender Boy, n (%) \| 1917 (52.5) \| 6720 (51.6); First-born child, n (%) \| 1292 (35.4) \| 6772 (52.0); Maternal preeclampsia / eclampsia, n (%) \| 54 (1.5) \| 30 (0.2) **Inclusion criteria** Not reported **Exclusion criteria** Not reported | **Interventions**  Planned caesarean birth versus vaginal birth | **Details**  Data obtained from the Taiwan birth cohort study database. Results reported as OR adjusted for: caesarean birth, gestational age, maternal age at birth, maternal education level, family income at 8 years of age and maternal diseases in pregnancy | **Included in the decision aid (main paper)**  None  **Supplementary outcomes**  ***Maternal***  None  ***Infant/neonatal***  Neurodevelopmental disorders (at 8 years of age) | Assessed using the Newcastle Ottawa quality assessment form for cohort studies  **Overall quality:** good |
| **Full citation**  Christiansen EL, Thagaard IN, Hedley PL, Lautrup Hansen MJ, Frithioff-Bøjsøe C, Larsen T, Holm JC, Christiansen M, Krebs L. The association between parental, obstetric, socioeconomic, and lifestyle factors and the development of childhood obesity: A Danish Cohort Study. medRxiv. 2023 Nov 24:2023-11. https://doi.org/10.1101/2023.11.23.23298961  **Ref Id**  Christiansen 2023  **Country/ies where the study was carried out**  Denmark  **Study type**  Retrospective cohort  **Aim of the study**  To investigate the association between several parental, obstetric and lifestyle characteristics and childhood overweight and obesity.  **Study dates**  Infants were born between January 1, 2006 and December 31, 2011. Questionnaires were sent between 2012 and 2016  **Source of funding**  Not reported | **Sample Size** N= 1448 children (caesarean before labour n= 159, caesarean during labour n= 159, vaginal birth n= 1130) **Characteristics** \| All children; Maternal age, years, mean (SD) \| 31.32 (4.69); Maternal pre=pregnancy BMI, kg/m^2^, median (IQR) \| 23.44 (5.70); Maternal European ancestry, n (%) \| 1,307 (90.3); Maternal smoking, n (%) \| 133 (9.2); Maternal gestational diabetes, n (%) \| 33 (2.3); Birthweight z score, median (IQR) \| `-0.01 (2.75); Birthweight categories, n (%) Low birth weight (<3000g) Average birth weight (≥ 3,000g, < 4,500g) macrosomia (≥4,500g) \| 165 (11.4) 1,219 (84.2) 64 (4.4); Birthweight relative to gestational age groups, n (%) Small for gestational age Average for gestational age Large for gestational age \| 48 (3.3) 1,323 (91.4) 77 (5.3) **Inclusion criteria** Not reported **Exclusion criteria** - Multifetal pregnancies - Preterm births - Stillborn or neonatal deaths - Children with invalid civil registration numbers - Mismatched mother child pairs - Mothers with no information on pre-pregnancy BMI - Children with missing data on gestational age, height and weight measures, or duration of breastfeeding | **Interventions**  Caesarean birth before labour versus vaginal birth. | **Details**  Clinical data related to pregnancy and birth was retrieved from the Danish Medical Birth Registry and the Astraia Database. Mothers were asked to complete a questionnaire regarding information on child growth, diseases, eating habits, family history and structure, parental anthropometry, socioeconomic status, and use of medication.  Results reported as OR adjusted for Study controls for ancestry, mothers age at birth, parity, smoking cigarettes during pregnancy, hypertensive disorder during pregnancy, diabetes during pregnancy, mode of birth, breastfeeding, allergies in childhood, use of antibiotics in childhood, fastfood habits, consumption of sugar containing drinks, consumption of sugar-free drinks, maternal SES, paternal SES, maternal change in BMI since pregnancy, paternal overweight/obese, and marital status | **Included in the decision aid (main paper)**  ***Maternal***  None  ***Infant/neonatal***  Childhood obesity  **Supplementary outcomes**  None | Assessed using the Newcastle Ottawa quality assessment form for cohort studies  **Overall quality:** good |
| **Full citation**  Currell A., Koplin J.J., Lowe A.J., Perrett K.P., Ponsonby A.L., Tang M.L.K., Dharmage S.C., Peters R.L. Mode of birth is not associated with food allergy risk in infants. J Allergy Clin Immunol Pract2022;10:2135-43. doi: https://doi.org/10.1016/j.jaip.2022.03.031  **Ref Id**  Currell 2022  **Country/ies where the study was carried out**  Australia  **Study type**  Population-based longitudinal cohort study  **Aim of the study**  To assess whether unplanned of planned caesarean, or caesarean birth in the presence or absence of labour initiation is associated with the risk of food allergy  **Study dates**  September 2007 to August 2011  **Source of funding**  The National Health & Medical Research Council (NHMRC) of Australia, the Ilhan Food Allergy Foundation, Anaphy-laxiStop, the Charles and Sylvia Viertel Medical Research Foundation, and the Victorian Government’s Operational Infrastructure Support Program. R.L.Peters, S. C.Dharmage, A.J.Lowe, and J.J. Koplin were supported by NHMRC awards. K. P. Perrett was supported by a Melbourne Children’s Clinician-Scientist Fellowship. | **Sample Size** N= 2045 (vaginal birth n=1428/2042; unplanned caesarean birth n=304/2042; planned caesarean birth n=310/2042) **Characteristics** \| Total N=2045; Maternal age, years, mean (SD) \| 33.1 (4.6); Mother’s country of birth, n (%) Australian Far east Asian UK/Europe Other \| 1526/2004 (76.2) 172/2004 (8.6) 153/2004 (7.6) 153/2004 (7.6); Socioeconomic status (1= most disadvantaged), n (%) 1 2 3 4 5 \| 397/2042 (19.4) 389/2042 (19.1) 483/2042 (23.7) 399/2042 (19.5) 374/2042 (18.3); Family history of allergy, n (%) \| 1492/2045 (73.0); Any food allergy at 12 months, N (%) \| 253/1944 (13.0); Any food sensitisation at 12 months, n (%) \| 367/1965 (18.7); Weight for gestational age (HealthNuts data), n (%) Normal Small Large \| 1721/2045 (84.2) 150/2045 (7.3) 174/2045 (8.5); Weight for gestational age (VPDC data), n (%) Normal Small Large \| 1704/2045 (83.3) 128/2045 (6.3) 213/2045 (10.4) **Inclusion criteria** Not reported **Exclusion criteria** Not reported | **Interventions**  Vaginal birth, planned and unplanned caesarean birth | **Details**  Data from the HealthNuts study of 5276 12 month-old infants who underwent testing for food allergies were link to additional birth data from the Victorian Perinatal Data Collection (VPDC).  Results were reported as OR adjusted for maternal country of birth, maternal age at birth, maternal smoking during pregnancy, prematurity (≤36 weeks of gestational age), socioeconomic index for areas and weight for gestational age. | **Included in the decision aid (main paper)**  None  **Supplementary outcomes**  ***Maternal***  None  ***Infant/neonatal***  Any food allergy  Egg allergy  Peanut allergy | Assessed using the Newcastle Ottawa quality assessment form for cohort studies    **Overall quality:** good |
| **Full citation**  Dachew, Berihun & Tessema, Gizachew & Alati, Rosa. (2022). Association between obstetric mode of birth and emotional and behavioural problems in children and adolescents: the children of the 90s health study. Social Psychiatry and Psychiatric Epidemiology. 58. 1-12. 10.1007/s00127-022-02374-z.  **Ref Id**  Dachew 2023  **Country/ies where the study was carried out**  UK  **Study type**  Population-based longitudinal birth cohort study  **Aim of the study**  To examine the association between obstetric mode of birth and emotional and behavioural problems in offspring aged 3–16 years.  **Study dates**  Used participants in the Avon Longitudinal Study of Parents and Children (ALSPAC) cohort, with women who had an expected birth date between 1st April 1991 and 31st December 1992. Followed participants up to 16 years. Didn’t report anything further.  **Source of funding**  CAUL and its Member Institutions. The UK Medical Research Council and Wellcome and the University of Bristol provide core support for ALSPAC. | **Sample Size** N= 13,280 children at baseline; 7074 at age 3; 6114 at age 7; 5639 at age 9; 5211 at age 11 and 4071 at age 16. (n= 1397 with 3.7% planned caesarean and 6.8% unplanned caesarean. n=11883 total vaginal birth with n= 10175 spontaneous vaginal birth and n= 1708 assisted birth **Characteristics** \| Caesarean birth \| Spontaneous vaginal birth; Maternal age at birth, years, mean (SD) \| 28.6 (5.0) \| 27.6 (4.9); Gestational age at birth, weeks, mean (SD) \| 38.6 (2.6) \| 39.6 (1.6); Birth weight, kg, mean (SD) \| 3.2 (0.7) \| 3.4 (0.5); Multiparous, n (%) \| 47.1% \| 62.1% **Inclusion criteria** - Women from the former county of Avon, UK, with expected birth dates between April 1, 1991, and December 31, 1992. - Women with singleton and live-born infants - Women had complete data on both the exposure (obstetric mode of birth) and outcomes (child emotional and behavioural problem measures). **Exclusion criteria** - Pre-term infants - Incomplete data | **Interventions**  Planned caesarean versus vaginal birth | **Details**  The sample consisted of individuals from the Avon Longitudinal Study of Parents and Children (ALSPAC) cohort.  Results were reported as OR adjusted for: maternal age, educational status, ethnicity, parity, pre-pregnancy body mass index, pregnancy diabetes, infection during pregnancy, hypertensive disorders during pregnancy, alcohol consumption during pregnancy, smoking during pregnancy, maternal antenatal depression and anxiety and offspring sex and gestational age at birth | **Included in the decision aid (main paper)**  None  **Supplementary outcomes**  ***Maternal***  None  ***Infant/neonatal***  Total behavioural difficulties (at age 3, 7, 9, 11, 16 years)  Emotional symptoms (at age 3, 7, 9, 11, 16 years)  Peer relationship problems (at age 7, 9, 11, 16 years)  Hyperactivity/inattention problems (at age 3, 7, 9, 11, 16 years)  Conduct problems (at age 3, 7, 9, 11, 16 years)  Prosocial behaviours (at age 3, 7, 9, 11, 16 years) | Assessed using the Newcastle Ottawa quality assessment form for cohort studies  **Overall quality:** good |
| **Full citation**  Einum A, Harmon QE, Sørbye LM, Nilsen RM, Morken NH. Associations between term cesarean birth in the first pregnancy and second‐pregnancy preterm birth. Acta Obstetricia et Gynecologica Scandinavica. 2025 Jan;104(1):68-76. <https://doi.org/10.1111/aogs.14996>  **Ref Id**  Einum 2024  **Country/ies where the study was carried out**  Norway  **Study type**  Retrospective cohort  **Aim of the study**  To explore the association between term caesarean birth in the first pregnancy and second-pregnancy preterm birth  **Study dates**  1999 to 2020  **Source of funding**  Odd Fellow Medical Fund; Gidske  and Peter Jacob Sørensens Fund;  Helse Vest, Grant/Award Number: F-12156/4800003677; National Institute of Environmental Health Sciences, Grant/Award Number: Z01ES103333 | **Sample Size** N= 298901 (n= 3604 planned caesarean birth in first pregnancy n= 29,907 unplanned caesarean birth in first pregnancy, n=265,390 vaginal birth in first pregnancy) **Characteristics** \| Planned caesarean birth \| Vaginal birth; Maternal age, years, n (%) <25 25-29 30-34 ≥35 \| 730 (20.2) 1192 (33.1) 1156 (32.1) 526 (14.6) \| 79710 (30.0) 110795 (41.8) 62 210 (23.4) 12 675 (4.8); Maternal smoking, n (%) Yes No Missing \| 395 (11.0) 2510 (69.6) 699 (19.4) \| 29 411 (11.1) 196 692 (74.1) 39 287 (14.8); Chronic maternal disease, n (%) Yes No \| 90 (2.5) 3514 (97.5) \| 3140 (1.2) 262 250 (98.8); Maternal placental disease, n (%) Yes No \| 283 (7.9) 3321 (92.1) \| 19 685 (7.4) 245 705 (92.6) **Inclusion criteria** Not reported **Exclusion criteria** Not reported | **Interventions**  Planned caesarean birth versus vaginal birth. | **Details**  298 901 mothers with first and second singleton deliveries from 1999 to 2020 were investigated using data from the Medical Birth Registry of Norway linked with Statistics Norway.  Results were reported as RR adjusted for maternal country of birth, year of first birth, maternal age at first birth,  smoking at the beginning of first pregnancy, chronic maternal disease (diabetes mellitus type 1 or  2, chronic hypertension, or kidney disease in the first pregnancy), gestational diabetes mellitus  in first pregnancy, and placental disease in first pregnancy (preeclampsia, eclampsia, HELLP-syndrome,  placental abruption, or fetal growth restriction (<fifth percentile of birthweight by  gestational age)). | **Included in the decision aid (main paper)**  ***Maternal***  Pre-term birth in the second pregnancy  ***Infant/neonatal***  None  **Supplementary outcomes**  None | Assessed using the Newcastle Ottawa quality assessment form for cohort studies    **Overall quality:** good |
| **Full citation**  Frijmersum ZZ, Van der Meij E, Bakker PC, De Vries R, Anema JR, Huirne JA. What birth related factors affect postpartum recovery? A systematic review. AJOG Global Reports. 2025 Apr 5:100496. https://doi.org/10.1016/j.xagr.2025.100496  **Ref Id**  Frijmersum 2025  **Country/ies where the study was carried out**  The review authors were based in The Netherlands. The authors do not report details of the countries where the included studies were conducted  **Study type**  Systematic review  **Aim of the study**  Toidentifythebirth-relatedfactorsthataffectpostpartumrecovery.  **Study dates**  Literature searches were conducted from inception to April 2024  **Source of funding**  The study received no funding | **Sample size**  K=38  Postpartum pelvic pain: k=1 study; pelvic pain at 12 months: k=1 study; upper back pain at 12 months: k=1 study; lower back pain at 12 months: k=1 study  **Inclusion criteria**  Cohort studies and RCTs of woman aged ≥18 years with a live born child reporting factors influencing post-partum recovery with follow up of at least 6 weeks after childbirth.  **Exclusion criteria**  Studies on personal and social factors. Outcomes regarding anatomic changes were not considered for inclusion in women who did not experience morbidity from these anatomic changes. | Caesarean birth, versus vaginal birth. Studies included in the review were of planned and unplanned caesarean and included assisted vaginal birth | PubMed, Embase, and Web of Science (core collection) were searched from inception to April 2024 | **Included in the decision aid (main paper)**  ***Maternal***  Pelvic pain, postpartum and at 12 months  Lower and upper back pain at 12 months  ***Infant/neonatal***  None  **Supplementary outcomes**  None | Assessed with the ROBIS checklist  **Risk of bias in the review:** Low |
| **Full citation**  Hagen S, Sellers C, Elders A, Glazener C, MacArthur C, Toozs‐Hobson P, Hemming C, Herbison P, Wilson D. Urinary incontinence, faecal incontinence and pelvic organ prolapse symptoms 20–26 years after childbirth: A longitudinal cohort study. BJOG: An International Journal of Obstetrics & Gynaecology. 2024 Dec;131(13):1815-23.  <https://doi.org/10.1111/1471-0528.17913>  **Ref Id**  Hagen 2024  **Country/ies where the study was carried out**  United Kingdom and New Zealand  **Study type**  Retrospective Cohort  **Aim of the study**  To investigate pelvic floor dysfunction (urinary incontinence,  faecal incontinence and prolapse) ≥20 years after childbirth and their association with birth mode history and demographic characteristics.  **Study dates**  2013 to 2014 New Zealand; 2019 to 2020 UK  **Source of funding**  Chief Scientist Office, Grant/Award Number: HIPS/17/09; University of Otago; Department of Women's and Children's Health (University of Otago); New Zealand Continence Association | **Sample Size** N= 2270 (n= 364 caesarean birth; n=1868 vaginal birth: n=1526 spontaneous, n=224 forceps, n=118 vacuum; missing n=38) **Characteristics** \| Total; Maternal age, years, median (IQR) \| 30.1 (6.2); Perineal trauma (vaginal births only) n/N (%) Intact Laceration Episiotomy Unknown \| 487/1819 (26.8) 857/1819 (47.1) 475/1819 (26.1) 87; Urinary incontinence at 3 months postpartum, n (%) Incontinent Uknown \| 796 (35.1) 0; Faecal incontinence, n (%) Incontinent Unknown \| 186 (8.8) 153 **Inclusion criteria** Not reported **Exclusion criteria** Not reported | Casearean birth (it is not reported whether the Caeseran births were planned or unplanned CB) versus vaginal birth. | **Details**  Women who gave birth in 1993-1994, who had not died, withdrawn or been lost to follow-up, were invited to take part in a postal survey. In New Zealand, this was carried out 20 years after the index birth, while in the United Kingdom, it was carried out at 26 years.  Results were reported as OR adjusted for age at first birth, number of births and current BMI, ethnic group | **Included in the decision aid (main paper)**  ***Maternal***  urinary incontinence,  faecal incontinence and prolapse  ***Infant/neonatal***  None  **Supplementary outcomes**  None | Assessed using the Newcastle Ottawa quality assessment form for cohort studies    **Overall quality:** good |
| **Full citation**  Hartley, Maria & Woolcott, Christy & Langley, Joanne & Brown, Mary & Ashley-Martin, Jillian & Kuhle, Stefan. (2020). Birth by Caesarean birth and otitis media in childhood: a retrospective cohort study. Scientific Reports. 10. 10.1038/s41598-020-62229-y.  **Ref Id**  Hartley 2020  **Country/ies where the study was carried out**  Canada  **Study type**  Retrospective Cohort Study  **Aim of the study**  To examine the association between birth by caesarean birth and health care use for Otitis Media in childhood in a large population-based sample in the Canadian province of Nova Scotia  **Study dates**  2003 to 2007 and followed through to 2014  **Source of funding**  Supported by a Nova Scotia/Canadian Institutes of Health Research Regional Partnership Program Operating Grant (FRN #134531) with matching funds from the Dalhousie Medical Research Foundation, the IWK Health Centre Foundation, the Department of Obstetrics and Gynaecology Atlee Endowment Fund, and the Department of Pediatrics, IWK Health Centre | **Sample Size** N= 36318 (n= 9729 caesarean didn’t report differentiation over planned and actual mode of birth and n= 26589 vaginal birth) **Characteristics** \| Caesarean birth \| Vaginal birth; Maternal age, years, mean (SD) \| 30 (5.4) \| 28 (5.6); Male offspring sex (%) \| 53 \| 50; Gestational diabetes mellitus (%) \| 5.3 \| 2.7; Parity (%) 0 1 2 ≥ 3 \| 48 37 11 3.5 \| 44 36 14 6.4; Birth weight for gestational age (%) Appropriate (10th–90th percentile) Small (<10th percentile) Large (>90th percentile) \| 72 6.9 21 \| 79 7.1 14 **Inclusion criteria** - Children born at 37 weeks’ gestation or later and had at least two months of follow-up in the administrative health databases. **Exclusion criteria** - Multiple births - Children with major congenital anomalies - Children born preterm - Less than two months of follow-up due to death or migration out of the province | **Interventions**  Caesarean versus vaginal birth  (only reported caesarean however stated planned CB was included in secondary analysis) | **Details**  The cohort was derived through a linkage of the Nova Scotia Atlee Perinatal Database with provincial administrative health data.  Results were reported as HR adjusted for maternal pre-pregnancy weight (not overweight or obese, overweight, obese), birth weight for gestational age (small, appropriate, large), maternal smoking during pregnancy, area-level income quintile, area of residence (rural versus urban), sex, parity (0, 1, 2, ≥3 excluding current pregnancy), maternal age at birth. | **Included in the decision aid (main paper)**  None  **Supplementary outcomes**  ***Maternal***  None  ***Infant/neonatal***  Risk of otitis media in childhood  Number of otitis media episodes  Otitis media episodes >95th percentile | Assessed using the Newcastle Ottawa quality assessment form for cohort studies  **Overall quality:** good |
| **Full citation**  Hjorth S, Kirkegaard H, Olsen J, et al. Mode of birth and long-term sexual health: a follow-up study of mothers in the Danish National Birth Cohort. BMJ Open 2019;9:e029517. doi: 10.1136/bmjopen-2019-029517  **Ref Id**  Hjorth 2019  **Country/ies where the study was carried out**  Denmark  **Study type**  Follow-up of the Danish National birth cohort (1996-2002)  **Aim of the study**  To investigate the relationship between mode of birth and women’s long-term sexual health  **Study dates**  December 2013 to December 2014  **Source of funding**  The Danish National Birth Cohort was established with a grant from the Danish National Research Foundation. Additional support was obtained from the Danish Regional Committees, the Pharmacy Foundation, the Egmont Foundation, the March of Dimes Birth Defects Foundation, the Health Foundation and other minor grants. The Danish Council for Independent Research supported the maternal follow-up. | **Sample Size** N= 37,417 (only spontaneous vaginal births n=23,608; instrumental vaginal birth (n=5003, only caesarean birth n=3244; spontaneous VBAC n=2038; instrumental VBAC n=457; caesarean birth after vaginal birth n=3067) **Characteristics** \| Only caesarean birth \| Spontaneous vaginal birth; Maternal age at first birth, years, n (%) <25 25-29 30-34 ≥35 \| 363 (11) 1433 (44) 1024 (32) 424 (13) \| 4864 (21) 12,758 (54) 5063 (21) 623 (4); Socio-occupational status, n (%) Low Middle High Missing \| 202 (7) 1056 (35) 1779 (59) 207 \| 1393 (6) 7444 (34) 13,318 (60) 1453; Pre-pregnant BMI, n (%) <18.5 18.5-24.9 25.0-29.9 ≥30 Missing \| 87 (3) 1843 (62) 697 (23) 355 (12) 262 \| 906 (4) 15,991 (73) 3748 (17) 1249 (6) 1714 **Inclusion criteria** - Women who participated in the follow-up and answered at least one sexual health question - Women with male and/or female partners **Exclusion criteria** - Women who did not have a partner | **Interventions**  only spontaneous vaginal births; instrumental vaginal birth; spontaneous VBAC; instrumental VBAC; caesarean birth after vaginal birth (no distinction is made between planned and non-planned) caesarean birth | **Details**  Follow-up questionnaire on physical, mental and sexual health of women enrolled into the Danish National birth cohort between 1996 and 2002.  Results were reported as OR adjusted for maternal age at first birth, calendar year at first birth, prepregnant body mass index, socio-occupational  status, self-assessed  health, disease, exercise in pregnancy and smoking in  pregnancy. | **Included in the decision aid (main paper)**  ***Maternal***  One or more sexual problems  Reduced desire  Difficulty in obtaining orgasm  Insufficient lubrication  Dyspareunia  Entry dyspareunia  Deep dyspareunia  ***Infant/neonatal***  None  **Supplementary outcomes**  None | Assessed using the Newcastle Ottawa quality assessment form for cohort studies    **Overall quality:** Good |
| **Full citation**  Legro Nicole R., Lehman Erik B., Kjerulff Kristen H., Mode of first birth and postpartum weight retention at 1 year. Obes Res Clin Pract. 2020 May-Jun;14(3):241-248. doi: 10.1016/j.orcp.2020.04.009. Epub 2020 May 23. PMID: 32456882.  **Ref Id**  Legro 2020  **Country/ies where the study was carried out**  USA  **Study type**  Prospective cohort study – Secondary data analysis study  **Aim of the study**  To analyse the association between mode of birth and postpartum weight retention at 12 months after birth  **Study dates**  Only reported dates from used First Baby Study (FBS) data  **Source of funding**  Eunice Kennedy Shriver National Institute of Child Health and Human Development, National Institutes of Health | **Sample Size** N= 2500 (n= 796 caesarean and n= 1951 vaginal birth) **Characteristics** \| Caesarean Birth \| Vaginal birth; Maternal age, years, n (%) 18-24 25-29 30-35 \| 135 (18.5) 295 (40.5) 298 (40.9) \| 472 (26.6) 724 (40.9) 579 (32.5); Pre-pregnancy BMI, ≥30 kg/m^2^, n (%) \| 220 (30.2) \| 294 (16.6); Gestational weight gain more than recommended by IOM guidelines, n (%) \| 467 (64.3) \| 887 (50.1); Postpartum weight retention at 12 month - Retained 10 pounds or more, n (%) \| 203 (27.9) \| 394 (22.2); BMI ≥30 kg/m^2^ at 12 months postpartum, n (%) \| 228 (31.3) \| 334 (18.8) **Inclusion criteria** - Nulliparous mothers, aged 18–35 at the time of recruitment with singleton pregnancies - English or Spanish speaking, and willing to be in a 3-year telephone interview study **Exclusion criteria** - Women who had a prior pregnancy of 20 weeks’ gestation or longer, planning to have a tubal ligation at the time of birth, surrogate pregnancy and planning to have the child adopted. - Women who delivered before 34 weeks’ gestation were also excluded - Women over 35 years old | **Interventions**  Caesarean birth (no distinction is made between planned and non-planned) versus vaginal birth | **Details**  Data obtained from the First Baby Study, a prospective cohort study of women who delivered their first child in Pennsylvania between 2009 and 2011, followed for three years after childbirth.  Results were reported as OR adjusted for pre-pregnancy body mass index, gestational weight gain, age, education, poverty status, smoking, race/ethnicity, gestational age, pregnancy complications, breastfeeding and exercise habits during pregnancy and in the first year after birth. | **Included in the decision aid (main paper)**  ***Maternal***  Postpartum weight retention at 1 year postpartum (retained 10 pounds or more)  ***Infant/neonatal***  None  **Supplementary outcomes**  None | Assessed using the Newcastle Ottawa quality assessment form for cohort studies    **Overall quality:** good |
| **Full citation**  Maher, Gillian & Khashan, Ali & McCarthy, Fergus. (2022). Obstetrical mode of birth and behavioural outcomes in childhood and adolescence: findings from the Millennium Cohort Study. Social Psychiatry and Psychiatric Epidemiology. 57. 1-13. 10.1007/s00127-022-02233-x.  **Ref Id**  Maher 2022  **Country/ies where the study was carried out**  UK  **Study type**  Population-based cohort study  **Aim of the study**  To examine the association between mode of birth (in particular caesarean birth) and behavioural outcomes in offspring at six time-points between age 3 and 17 years.  **Study dates**  The Millennium Cohort Study included children born between 2000 and 2002, study followed up till 17 years.  **Source of funding**  The IReL Consortium and the Health Research Board (HRB), Ireland. | **Sample Size** N= 18213 at baseline; 13,600 at age 3; 13,831 at age 5; 12,687 at age 7; 11,055 at age 11; 10,745 at age 14 and 8839 at age 17 (n= 1440 planned caesarean birth; n=1253 unplanned caesarean birth and n= 1180 CS after induction. n= 14340 total vaginal birth including n=8866 spontaneous VD; n= 1735 assisted VD and n= 3739 induced VD) **Characteristics** \| Planned caesarean birth \| Spontaneous vaginal birth; Maternal age, years, mean (SD) \| 30.75 (5.34) \| 27.92 (5.89); Maternal smoking status, n (%) Smoked during pregnancy \| 19.44% \| 23.89%; Maternal alcohol consumption during pregnancy, n (%) - Yes \| 30.00% \| 29.47%; Parity (first born child), n (%) \| 24.31% \| 34.37%; Total SDQ, mean (SD) Age 3 years (N=13,600) Age 5 years (N=13,831) Age 7 years (N=12,687) Age 11 years (N=11,055) Age 14 years (N=10,745) Age 17 years (N=8839) \| 9.12 (5.17) 6.88 (4.72) 7.14 (5.12) 7.14 (5.43) 7.74 (5.85) 7.55 (6.17) \| 9.63 (5.30) 7.37 (4.98) 7.44 (5.43) 7.37 (5.72) 8.17 (6.01) 7.90 (6.19) **Inclusion criteria** - Singleton mother–child pairs - Those who remained living in the (UK) at the time of sampling. **Exclusion criteria** - Preterm births (gestational age <37 weeks’) Multiple births | **Interventions**  Planned caesarean vs vaginal birth | **Details**  The Millennium Cohort Study (MCS) follows children born between 2000 and 2002 in 398 areas across the UK.  Results were reported as OR adjusted for: maternal age, maternal education, maternal smoking status, maternal alcohol consumption during pregnancy, pre-pregnancy body mass index (BMI), household income, small for gestational age (SGA), infant sex, parity, hypertensive disorders of pregnancy (including raised blood pressure, eclampsia/ preeclampsia or toxaemia) and maternal depression/serious anxiety. | **Included in the decision aid (main paper)**    None  **Supplementary outcomes**  ***Maternal***  None  ***Infant/neonatal***  Emotional difficulties (at age 3, 5, 7, 11, 14, 16, 17 years)  Conduct difficulties (at age, 3, 5, 7, 11, 14, 16, 17 years)  Hyperactivity (at age 3, 5, 7, 11, 14, 16, 17 years)  Peer problems (at age 3, 5, 7, 11, 14, 16, 17 years)  Prosocial behaviour (at age 3, 5, 7, 11, 14, 16, 17 years) | Assessed using the Newcastle Ottawa quality assessment form for cohort studies  **Overall quality:** good |
| **Full citation**  Matsumoto, N., Mitsui, T., Tamai, K.*et al.* Cesarean birth on child health and development in Japanese nationwide birth cohort. *Sci Rep*15, 2485 (2025). <https://doi.org/10.1038/s41598-025-87043-2>  **Ref Id**  Matsumoto 2025  **Country/ies where the study was carried out**  Japan  **Study type**  Prospective population-based cohort  **Aim of the study**  To investigate the long-term effects of caesarean birth on child health and development  **Study dates**  The 21st Century Longitudinal Survey of Newborns, encompasses all 43,767 infants born in Japan between May 10 and 24, 2010  **Source of funding**  supported by JSPS KAKENHI Grant Number JP23K16329 | **Sample Size** N= 2114 infants (n= 763 caesarean births, and n= 1351 vaginal births) **Characteristics** \| Caesarean birth \| Vaginal birth; Preterm birth, n (%) \| 196 (25.7%) \| 119 (8.8%); Birthweight, g, median (IQR) \| 2,670 (2,255-3,005) \| 2,988 (2,710-3,240); Low birth weight < 2500 g, n (%) \| 279 (36.6%) \| 192 (14.2%); Maternal age at birth, years, n (%) <30 30-34 ≥35 \| 180 (23.6%) 274 (35.9%) 309 (40.5%) \| 454 (33.6%) 476 (35.2%) 421 (31.2%); Maternal smoking during pregnancy, n (%) \| 31 (5.3%) \| 50 (5.0%); Maternal alcohol consumption during pregnancy, n (%) \| 12 (2.1%) \| 49 (5.0%) **Inclusion criteria** Not reported **Exclusion criteria** 26 children with ambiguous birth mode responses (e.g., “other”) were excluded. | **Interventions**  Caesarean birth versus vaginal birth. No distinction is made between planned and unplanned caesarean birth. | **Details**  Analysis of data from 2,114 children in a nationwide Japanese birth cohort, linking the 21st Century Longitudinal Survey of Newborns with the Perinatal Research Network database. Questionnaires are sent annually to guardians until infants are aged 5.5 years, with further follow-up at ages 7, 8 and 9 years. Data from this cohort were linked with the Perinatal Research Network (PRN) database that contains clinical information regarding maternal characteristics, preexisting medical conditions, pregnancy complications, birth details, neonatal transport, and other relevant factors.  Results are reported as RR adjusted for maternal age at birth, multiple births, multiparity, fetal presentation, presence of fetal anomalies, maternal transport, pre-existing maternal medical conditions, pregnancy complications, maternal smoking during pregnancy, maternal alcohol consumption during pregnancy, maternal education attainment, paternal age at birth, paternal education attainment, and place of residence at birth. In the obesity outcome model, maternal pre-pregnancy BMI (continuous) was also added as an adjustment variable. | **Included in the decision aid (main paper)**  ***Maternal***  None  ***Infant/neonatal***  Overweight/obesity at ages 5.5 and 9 years  Motor milestones not attained by the expected time (2.5 years)  Language milestones not attained by the expected time (2.5 years)  Cognitive milestones not attained by the expected time (5.5 years)  **Supplementary outcomes**  None | Assessed using the Newcastle Ottawa quality assessment form for cohort studies  **Overall quality:** good |
| **Full citation**  Miller, Jessica & Goldacre, Raphael & Id, Hannah & Zeltzer, Justin & Knight, Marian & Morris, Carole & Nowell, Sian & Id, Rachael & Carter, Kim & Id, Parveen & Id, Nicholas & Strunk, Tobias & Li, Jiong & Nassar, Natasha & Id, Lars & Id, David. (2020). Mode of birth and risk of infection-related hospitalisation in childhood: A population cohort study of 7.17 million births from 4 high-income countries. PLoS Medicine. 17. 10.1371/journal.pmed.1003429.  **Ref Id**  Miller 2020  **Country/ies where the study was carried out**  Denmark, Scotland, England and Australia  **Study type**  Multi-country Population-based cohort study  **Aim of the study**  Investigated the relationship between mode of birth and childhood infection-related hospitalisation in high-income countries with varying caesarean rates  **Study dates**  2001 to 2010  **Source of funding**  National Health and Medical Research Council; DHB Foundation; Health Research Fund of Central Denmark Region; Novo Nordisk Foundation; Danish Council for Independent Research; Financial Markets Foundation for Children; Raine Foundation Clinician Research Fellowship; Li Ka Shing Foundation; Robertson Foundation; Medical Research Council; British Heart Foundation and the NIHR Oxford Biomedical Research Centre | **Sample Size**  N= 7,174,787 (n= 727,755 planned caesarean including Denmark n= 56567; Scotland n= 74841; England n= 375864, New South Wales n= 150049 and Western Australia n= 70434. n= 691471 planned vaginal birth including Denmark n= 649955; Scotland n= 537816; England n= 3312535; New South Wales n= 683940 and Western Australia n= 308575)  **Characteristics**  Rich data provided from each country, for further detail please the original manuscript (Miller 2020)  **Inclusion criteria**   - All recorded singleton live births from January 1, 1996 to December 31, 2015 - Gestational age range was 24 to 43 weeks, with the exception of England where gestational age data for 42 weeks were not deemed reliable and excluded   **Exclusion criteria**   - Children with congenital malformations - England gestational age data for <30 weeks were not deemed reliable and excluded | **Interventions**  Planned caesarean birth versus vaginal birth | **Details**  Data were sourced from population-level databases in Denmark, Scotland, England, and Australia (New South Wales and Western Australia), including linked administrative and hospital data. Children were followed from birth-related hospital discharge until a maximum age of 5 years.  Results were reported as HR adjusted for smoking during pregnancy, maternal age at birth, parity, gestational age, birth weight, sex, birth year, season of birth, socioeconomic status and recorded hypertensive disorders or diabetes mellitus | **Included in the decision aid (main paper)**  None  **Supplementary outcomes**  ***Maternal***  None  ***Infant/neonatal***  Infant hospital-related infection by age of first occurrence (0-3 months, 4-6 months, 7-12 months, 1-2 years, 2-5 years)  At least one infection-related hospitalisation until 5 years old  Infection-related hospitalisation: invasive bacterial; skin and soft tissue; genitourinary; lower respiratory tract; upper respiratory tract; viral; gastrointestinal | Assessed using the Newcastle Ottawa quality assessment form for cohort studies  **Overall quality:** good |
| **Full citation**  Mitselou, Niki & Hallberg, Jenny & Stephansson, Olof & Almqvist Malmros, Catarina & Melen, Erik & Ludvigsson, Jonas. (2020). Adverse pregnancy outcomes and risk of later allergic rhinitis – Nationwide Swedish cohort study. Pediatric Allergy and Immunology. 31. 10.1111/pai.13230.  **Ref Id**  Mitselou 2020  **Country/ies where the study was carried out**  Sweden  **Study type**  Population- based prospective cohort study  **Aim of the study**  To examine pregnancy outcome (caesarean birth, preterm birth, low birthweight) and offspring allergic rhinitis as defined by national registers.  **Study dates**  2001-2013  **Source of funding**  The Swedish Research Council, Stockholm County Council (ALF), Swedish Lung-Heart Foundation and Swedish Research Council, Örebro County Council (ALF) | **Sample Size** N= 1,059,600 (n= 83923 planned caesarean birth; n= 86576 unplanned caesarean birth and n= 889101 vaginal birth) **Characteristics** \| Children without allergic rhinitis N (%) \| Children with allergic rhinitis N (%; Total births \| 1 037 214 (97.9) \| 22 386 (2.1); Caesarean birth \| N/A \| 2.34%; Vaginal birth \| N/A \| 2.10%; Maternal asthma/pulmonary disease (No) \| 92.5% \| 84.8% **Inclusion criteria** - Singleton livebirths, aged range of 0 months to 13 years who had complete data on all variables at the end of follow-up - Individuals born in Sweden between 2001 – 2012 **Exclusion criteria** - Not reported | **Interventions** Planned caesarean birth versus vaginal birth | **Details**  Data between the Medical Birth Register and National Patient Register were linked through the unique personal identity number assigned to all Swedish residents.  Results were reported as HR adjusted for infant sex and maternal factors (age at birth, country of birth, parity, early-pregnancy smoking, BMI, asthma/pulmonary disease) | **Included in the decision aid (main paper)**  None  **Supplementary outcomes**  ***Maternal***  None  ***Infant/neonatal***  Allergic rhinitis | Assessed using the Newcastle Ottawa quality assessment form for cohort studies  **Overall quality:** good  **Other information**  No reports on the number of pre-term births |
| **Full citation**  Mueller, Noel & Zhang, Mingyu & Hoyo, Cathrine & Østbye, Truls & Benjamin Neelon, Sara. (2019). Does Cesarean birth impact infant weight gain and adiposity over the first year of life?. International Journal of Obesity. 43. 10.1038/s41366-018-0239-2.  **Ref Id**  Mueller 2019  **Country/ies where the study was carried out**  USA  **Study type**  Prospective cohort study  **Aim of the study**  To examine differences in infant weight and adiposity trajectories from birth to 12 months by birth mode.  **Study dates**  2013-2015  **Source of funding**  National Institutes of Health (R01DK094841). NTM is supported by the National Heart, Lung, and Blood Institute of the National Institutes of Health under award number K01HL141589, and by grants from the MidAtlantic Nutrition Obesity Research Center (P30DK072488) and the Foundation for Gender Specific Medicine. | **Sample Size** N= 563 infants (n= 179 caesarean no differentiation over planned and actual mode of birth and n= 384 vaginal births including n=17 operative and n= 367 spontaneous) **Characteristics** \| Caesarean birth \| Vaginal birth; Maternal pre-pregnancy BMI, kg/m^2^, mean (SD) \| 33.97 (10.77) \| 28.25 (7.81); Age, years, mean (SD) \| 28.38 (5.35) \| 27.09 (5.95); Any breastfeeding (ever), n (%) \| 146 (81.6) \| 329 (85.7); Gestational age, week, mean (SD) \| 38.35 (1.71) \| 38.70 (1.51); Birth weight, kg, mean (SD) \| 3.24 (0.57) \| 3.19 (0.50); Infant birth weight for gestational age z-score, mean (SD) \| 0.25 (1.08) \| 0.00 (0.99) **Inclusion criteria** - Infant had to be a singleton born after 28 weeks’ gestation with no congenital abnormalities. **Exclusion criteria** - Observations with missing continuous variables in the models. - Underweight maternal pre-pregnancy BMI | **Interventions**  Caesarean birth versus vaginal birth | **Details**  Women were recruited from a private prenatal clinic and the county health department prenatal clinic in Durham, North Carolina, USA.  Results were reported as adjusted mean differences. Adjusted for maternal age at birth, race, household income, educational level, marital status, smoking status, infant birth weight, maternal pre-pregnancy body mass index | **Included in the decision aid (main paper)**  None  **Supplementary outcomes**  ***Maternal***  None  ***Infant/neonatal***  Weight-for-length at 12 months  Abdominal skinfold thickness at 12 months 9mm)  Subscapular skinfold thickness at 12 months (mm)  Triceps skinfold thickness at 12 months (mm)  Subscapular and triceps skinfold thickness at 12 months (mm) | Assessed using the Newcastle Ottawa quality assessment form for cohort studies  **Overall quality:** good |
| **Full citation**  Ning J, Deng J, Li S, Lu C, Zeng P. Meta-analysis of association between caesarean birth and postpartum depression risk. Frontiers in Psychiatry. 2024 Mar 28;15:1361604. <https://doi.org/10.3389/fpsyt.2024.1361604>  **Ref Id**  Ning 2024  **Country/ies where the study was carried out**  The review authors are based in Naning, China. The included studies were conducted in: Armenia, China, Denmark, Finland, India, Iran, Japan, Norway, Republic of Korea, Saudi Arabia, Sweden, USA  **Study type**  Systematic review and meta-analysis of 14 cohort studies and 4 case-control reports  **Aim of the study**  To explore the relationship between caesarean birth and the risk of postpartum depression (PPD).  **Study dates**  The initial literature search covered inception to December 2022. An updated literature search was conducted on 27^th^ February 2024.  **Source of funding**  Innovation Project of Guangxi Graduate Education (JYG2023082) and the Study on the construction and ability improvement of the Integrated Eldercare Services with Medical Care in Nanning from the perspective of supply side (RH2300019361). | **Sample size**  K=18 studies  Postpartum depression: k=3 studies  **Characteristics**  The youngest reported maternal age was 16 years and the oldest was 39 years. PPD assessment was conducted between 3 days and 12 months after birth. See review for additional details.  **Inclusion criteria**  Cohort and case control studies of parturients with PPD, regardless of race, region, whether they were primiparae and multiparae, or history of depression reporting PPD scores (either self-report or clinician administered measures).  **Exclusion criteria**  Studies in which PPD was managed using alternative therapies or treatments, such as diet, behavioural therapy, or antidepressants; meta-analyses, clinical trials, reviews, editorials, preprints, animal studies, and other types of literature; studies with ambiguous or incomplete data, which were not obtainable after contacting the authors, or studies in which data could not be transformed or combined; studies with a quality assessment score of ≤ 4 on the Newcastle-Ottawa Scale. | **Interventions**  The exposure group were parturients undergoing CB (including planned and unplanned CB). The comparison group were parturients undergoing vaginal birth  (including natural and assisted vaginal birth). | **Details**  A systematic literature search of PubMed, Web of Science, Cochrane  Library, and EMBASE was conducted up to 27 February 2024.  The meta-analysis results are reported as adjusted ORs. The included studies adjusted for various confounders. | **Included in the decision aid (main paper)**  ***Maternal***  Postpartum depression  ***Infant/neonatal***  None  **Supplementary outcomes**  None | Assessed with the ROBIS checklist  **Risk of bias in the review:**  Low |
| **Full citation**  Salem Yasmin, Oestreich MA, Oestreich Marc-Alexander, Fuchs Oliver Usemann Jakob, Frey Urs, Surbek Daniel, Amylidi-Mohr Sofia, Latzin Philipp, Ramsey Kathryn, Yammine Sophie. Are children born by cesarean birth at higher risk for respiratory sequalae? Am J Obstet Gynecol 2022;226:257.e1-11.  **Ref Id**  Salem 2022  **Country/ies where the study was carried out**  Switzerland  **Study type**  Prospective cohort study  **Aim of the study**  To assess whether the mode of birth is associated with changes in respiratory and atopic outcomes during infancy and at school age.  **Study dates**  April 1999  **Source of funding**  Supported by the Swiss National Science Foundation | **Sample Size** N= 578 (n= 47 planned caesarean and n= 464 vaginal birth) **Characteristics** \| Planned caesarean birth \| Vaginal birth; Gestational age, weeks, mean (SD) \| 38.6 (0.8) \| 39.8 (1.1); Maternal age, years, mean (SD) \| 36 (4) \| 33 (4); Transient signs of respiratory distress at birth (yes), n (%) \| 7 (15) \| 70 (15); Maternal smoking during pregnancy (yes), n (%) \| 6 (13) \| 79 (17); Parental smoking (yes), n (%) \| 5 (11) \| 58 (13) **Inclusion criteria** - Children born between April 1999 and May 2019 at or after 37 weeks of gestation. - Infants assessed for at least 40 weeks. **Exclusion criteria** - Those who were premature (birth at <37 weeks’ gestation), had congenital malformation and substantial perinatal disease. - Women who had severe maternal health problems, and maternal drug abuse excluding smoking. - Later excluded, children with transient signs of respiratory distress and children with antepartum administration of maternal antibiotics | **Interventions**  Planned caesarean birth versus vaginal birth | **Details**  Data obtained from Bern Basel Infant Lung Development birth cohort.  Results were reported as OR adjusted for sex, gestational age, birthweight, maternal age at birth, duration of exclusive breastfeeding, maternal smoking during pregnancy, older siblings, attendance of childcare, atopy  status of the mother, and current parental smoking | **Included in the decision aid (main paper)**  ***Maternal***  None  ***Infant/neonatal***  Asthma  **Supplementary outcomes**  ***Maternal***  None  ***Infant/neonatal***  Weeks with any respiratory symptom  Ever wheezing at early-school age  Allergic rhinoconjunctivitis  Atopic dermatitis | Assessed using the Newcastle Ottawa quality assessment form for cohort studies  **Overall quality:** good |
| **Full citation**  Sima YT, Magnus MC, Kvalvik LG, Morken NH, Klungsøyr K, Skjærven R, Sørbye LM. The relationship between cesarean birth and fecundability: a population-based cohort study. American Journal of Obstetrics and Gynecology. 2024 Jun 1;230(6):667-e1**.** [https://doi.org/10.1016/j.ajog.2023.10.02](https://doi.org/10.1016/j.ajog.2023.10.029)9  **Ref Id**  Sima 2024  **Country/ies where the study was carried out**  Norway  **Study type**  Population-based cohort study  **Aim of the study**  To assess the bidirectional relationship between caesarean birth and fecundability  **Study dates**  Pregnant women were recruited to the MoBa cohort study throughout Norway at the time of routine second-trimester ultrasound screening between 1999 and 2008. Data files released in 2019 were used as the basis for the study.  **Source of funding**  supported by the Norwegian Ministry of Health and Care Services and the Ministry of Education and Research | **Sample Size** N= 42,379 (n= 5153 (12.2%) planned and unplanned caesarean births and n= 37,226 (87.8%) vaginal births **Characteristics** \| Total n=42,379; Maternal age, years, n (%) <25 25-34 ≥35 \| 9128 (21.7) 31,416 (74.1) 1835 (4.3); Maternal BMI, kg/m^2^, n (%) <18.5 18.5-24.9 25-29.9 ≥30 Missing \| 1052 (2.5) 26,265 (62.0) 9882 (23.3) 4226 (10.0) 954 (2.3); Maternal smoking, n (%) Non-smoker Quit smoking during current pregnancy Current smoker Missing \| 31,154 (73.5) 7275 (17.2) 3198 (7.6) 752 (1.8); Self-reported chronic conditions, n (%) None One or more \| 34,947 (82.5) 7432 (17.5) **Inclusion criteria** Women with at least 1 recorded pregnancy in MoBa **Exclusion criteria** Women who did not complete the recruitment questionnaire, women with incomplete time to pregnancy data, women without a registered birth before the index pregnancy and those with in vitro fertilization in their previous pregnancy | **Interventions**  Planned caesarean birth versus vaginal birth (the authors report a sub-analysis of planned caesarean versus vaginal birth but do not report the number of women who had planned caesarean births) | **Details**  This was a prospective cohort study based on data from the Norwegian Mother, Father, and Child Cohort (MoBa) study linked with the Medical Birth Registry of Norway (MBRN), and analysed self-reported questionnaires completed at 15 to 18 weeks’ gestation.  Results were reported as RR adjusted for maternal age and pregnancy complications in the previous birth, maternal education, smoking, prepregnancy body mass index and chronic conditions,  and accounting for women who participated with several pregnancies. | **Included in the decision aid (main paper)**  ***Maternal***  Infertility  ***Infant/neonatal***  None  **Supplementary outcomes**  None | Assessed using the Newcastle Ottawa quality assessment form for cohort studies  **Overall quality:** good |
| **Full citation**  Tanoey, Justine & Gulati, Amit & Patterson, Chris & Becher, Heiko. (2019). Risk of Type 1 Diabetes in the Offspring Born through Planned or Non-planned Caesarean Birth in Comparison to Vaginal Birth: a Meta-Analysis of Observational Studies. Current Diabetes Reports. 19. 10.1007/s11892-019-1253-z.  **Ref Id**  Tanoey 2019  **Country/ies where the study was carried out**  UK, Denmark, Sweden and Australia  **Study type**  Systematic review and meta-analysis  **Aim of the study**  To summarise the effects of planned and non-planned CS on Type 1 Diabetes risk in the offspring  **Study dates**  Study published 11 November 2019. Included studies follow up between 1975 to 2012.  **Source of funding**  The authors JT and HB were supported by the German Federal Ministry of Education and Research (Grant Number 01ER1306 PERGOLA).v | **Sample Size**  K=9 studies  Type 1 diabetes: k=9 studies  **Characteristics**  See review for details  **Inclusion criteria**   - Original studies that reported effect sizes or adequate data for calculation of type 1 diabetes risk in children or young adults associated with planned CS and non-planned CS   **Exclusion criteria**   - Studies which only reported CS without specifying whether planned or non-planned (unplanned) - Studies which investigated only children with high-risk HLA genotype | **Interventions**  Planned caesarean versus vaginal birth | **Details**  Searches were conducted in Medline, Web of Science and Cumulative Index to Nursing and Allied Health Literature (CINAHL) databases for articles published before 23rd April 2018.  Included studies adjusted for various confounders, mainly maternal diabetes, maternal age, and gestational age and controlled for siblings. | **Included in the decision aid (main paper)**  ***Maternal***  None  ***Infant/neonatal***  Type 1 diabetes  **Supplementary outcomes**  None | Systematic review assessed with the ROBIS checklist  **Risk of bias in the review:** Low |
| **Full citation**  Vaajala M, Liukkonen R, Mattila VM, Kekki M, Kuitunen I. Birth mode and subsequent birth rate: A nationwide register‐based analysis in Finland. International Journal of Gynecology & Obstetrics. 2025 Mar;168(3):1161-70. <https://doi.org/10.1002/ijgo.15982>  **Ref Id**  Vaajala 2024  **Country/ies where the study was carried out**  Finland  **Study type**  Retrospective cohort  **Aim of the study**  To calculate the subsequent birth rate for different birth modes, comparing them with spontaneous vaginal deliveries, using a comprehensive nationwide high-quality registry  **Study dates**  First births occurred between 2004 to 2016. Second pregnancies between 2004 to 2018 were retrieved and combined with the data of the first births  **Source of funding**  The study did not receive funding | **Sample Size** N=375,619 (n= 22021 planned caesarean births, 50426 unplanned caesarean births, n= 252593 spontaneous vaginal births, n=50579 assisted vaginal births) **Characteristics** \| Planned caesarean birth \| Spontaneous vaginal birth; Maternal age, years, mean (SD) \| 30.4 (5.6) \| 27.8 (5.2); Maternal age, years, n (%) <20 20-24 25-29 30-34 35-39 ≥40 \| 508 (2.3) 2888 (13.1) 6401 (29.1) 6997 (31.8) 4029 (18.3) 1198 (5.4) \| 12,207 (4.8) 58,166 (23.0) 90 420 (35.8) 64,914 (26.1) 22 744 (9.0) 4127 1.6; Maternal BMI, kg/m^2^, mean (SD) \| 24.7 (5.1) \| 23.7 (4.5); Maternal smoking, n (%) \| 3509 (15.9) \| 45,618 (18.1) **Inclusion criteria** First pregnancy was a singleton pregnancy, pregnancies ending in birth after gestational week 21 + 6. data on pregnancies ending in miscarriage or induced abortions were not available in our data; however, stillbirths were included. **Exclusion criteria** Not reported | **Interventions**  Planned caesarean birth versus spontaneous vaginal birth | **Details**  Data from the National Medical Birth Register (MBR) were used to evaluate  the birth rate after different birth modes.    Results were reported as HR adjusted for categorized maternal age and body mass index, year of pregnancy, gestational diabetes, socioeconomic status, smoking status, prior miscarriages, and prior induced abortions | **Included in the decision aid (main paper)**  ***Maternal***  Became pregnant again after their first birth  ***Infant/neonatal***  None  **Supplementary outcomes**  None | Assessed using the Newcastle Ottawa quality assessment form for cohort studies  **Overall quality:** good |
| **Full citation**  Yoshida, T., Matsumura, K., Hatakeyama, T. *et al.* Association between Cesarean birth and neurodevelopmental disorders in a Japanese birth cohort: the Japan Environment and Children’s Study. *BMC Pediatr* 23, 306 (2023). <https://doi.org/10.1186/s12887-023-04128-5>  **Ref Id**  Yoshida 2024  **Country/ies where the study was carried out**  Japan  **Study type**  Retrospective cohort study  **Aim of the study**  To investigate the possible associations between mode of birth and presence of neurodevelopmental disorders at age 3 years.  **Study dates**  Pregnant women were recruited between 2011 and 2014. Datasets were released in April 2021.  **Source of funding**  The Ministry of the Environment, Japan | **Sample Size** N= 65,701 mother–toddler pairs (n = 12,174 caesarean births, n = 53,527 vaginal births) **Characteristics** \| Caesarean births \| Vaginal births; Maternal age, years, mean (SD) \| 32.7 (4.9) \| 31.1 (4.8); Pre-pregnancy BMI kg/m^2^, mean (SD) \| 21.9 (3.8) \| 21.0 (3.0); Maternal smoking, n (%) Never Former Current \| 7,072 (58.1) 4,624 (38.0) 478 (3.9) \| 32,518 (60.8) 19,158 (35.8) 1,851 (3.5); Maternal history of depression, anxiety disorder, dysautonomia, or schizophrenia, n (%) No Yes \| 10,346 (85.0) 1,828 (15.0) \| 46,198 (86.3) 7,329 (13.7); Infant birthweight, g, mean (SD) \| 2,875.8 (500.4) \| 3,062.9 (376.3); Infant major congenital abnormality, n (%) No Yes \| 11,751 (96.5) 423 (3.5) \| 52,467 (98.0) 1,060 (2.0) **Inclusion criteria** Not reported **Exclusion criteria** Multiple participations, multiple births, and miscarriages or stillbirths, missing information about a history of caesarean birth or missing answers about neurodevelopmental disorders | **Interventions**  Caesarean birth versus vaginal birth. No distinction is made between planned and unplanned caesarean births. | **Details**  This study analysed data obtained from the Japan Environment and Children’s Study (JECS), an ongoing birth cohort study.  Results were reported as OR adjusted for maternal age, pre-pregnancy body mass index, parity, history of depression, anxiety disorder, dysautonomia, or schizophrenia, history of physical disease, pregnancy complication, marital status, employed, highest education level, annual household income, alcohol intake, smoking status, negative attitude toward pregnancy, any major congenital anomaly, gestational age, and birth weight | **Included in the decision aid (main paper)**  ***Maternal***  None  ***Infant/neonatal***  Motor delay  Intellectual disability (including language delay)  Autism spectrum disorder  **Supplementary outcomes**  None | Assessed using the Newcastle Ottawa quality assessment form for cohort studies  **Overall quality:** Good |
| **Full citation**  Zhang, Tianyang & Sidorchuk, Anna & Sevilla-Cermeño, Laura & Vilaplana-Pérez, Alba & Chang, Zheng & Larsson, Henrik & Mataix-Cols, David & Fernández de la Cruz, Lorena. (2019). Association of Cesarean Birth With Risk of Neurodevelopmental and Psychiatric Disorders in the Offspring: A Systematic Review and Meta-analysis. JAMA Network Open. 2. e1910236. 10.1001/jamanetworkopen.2019.10236.  **Ref Id**  Zhang 2019  **Country/ies where the study was carried out**  Japan, Poland, Lebanon, USA, Canada, Denmark, Taiwan, Sweden, Norway, Finland, China, Australia, Egypt, Iraq, Turkey, UK, Brazil, South Korea, Iran, Spain and Ireland  **Study type**  Systematic review and meta-analysis  **Aim of the study**  To evaluate the association between caesarean birth and risk of neurodevelopmental and psychiatric disorders in the offspring.  **Study dates**  Study was published in August 28, 2019. Include studies had study periods between 1966 to 2016.  **Source of funding**  China Scholarship Council (Ms Zhang) | **Sample Size**  K=61 studies  Autism spectrum condition: k=8 studies  **Characteristics**  See review for details  **Inclusion criteria**   - Observational studies that allowed estimation of the associations between obstetric mode of birth (caesarean vs vaginal birth) and neurodevelopmental and psychiatric disorders in the offspring. - Studies reporting the method of exposure ascertainment (i.e., self-report or birth records) - Studies with the outcome diagnoses assessed through structured interviews or using standardised diagnostic criteria (e.g., International Classification of Diseases, DSM, or equivalent).   **Exclusion criteria**   - Review articles, book chapters, conference abstracts, and dissertations. - Self-reported or caregiver reported outcomes - Studies with lack of healthy comparison group - Studies which did not report caesarean birth | **Interventions**  Caesarean birth (any type, including planned and unplanned – data were extracted for elective caesarean as reported in the review’s supplementary material) versus vaginal birth (including assisted vaginal birth [use of vacuum or forceps] and unassisted vaginal birth | **Details**  Searches were conducted in Ovid MEDLINE, Embase, Web of Science, and PsycINFO from inception to December 19, 2018.  Included seven low/middle income countries: China, Brazil, Egypt, Iran, Iraq, Lebanon, Turkey  Results were reported as OR adjusted for sample size, caesarean birth type (planned/unplanned) or vaginal birth type (assisted/unassisted), participants in each group, and risk estimates (ORs, HRs) | **Included in the decision aid (main paper)**  ***Maternal***  None  ***Infant/neonatal***  Autism Spectrum condition  **Supplementary outcomes**  ***Maternal***  None  ***Infant/neonatal***  Tic disorder  Affective and non-affective psychoses  Childhood ADHD | Systematic review assessed with the ROBIS checklist  **Risk of bias in the review:** Low |
| **Full citation**  Zhang, Tianyang & Brander, Gustaf & Mantel, Ängla & Kuja-Halkola, Ralf & Stephansson, Olof & Chang, Zheng & Larsson, Henrik & Mataix-Cols, David & Fernández de la Cruz, Lorena. (2021). Assessment of Cesarean Birth and Neurodevelopmental and Psychiatric Disorders in the Children of a Population-Based Swedish Birth Cohort. JAMA network open. 4. e210837. 10.1001/jamanetworkopen.2021.0837.  **Ref Id**  Zhang 2021  **Country/ies where the study was carried out**  Sweden  **Study type**  Population-based cohort study  **Aim of the study**  To examine the association between caesarean birth (planned and intrapartum) and neurodevelopmental and psychiatric disorders in children.  **Study dates**  Statistical analyses were performed from September 26, 2019, to January 16, 2021.  **Source of funding**  China Scholarship Council (Ms Zhang) and the ìShizu Matsumuraîs Donation (Dr Fernández de la Cruz) | **Sample Size** N= 1179341 (n= 59514 planned caesarean birth and 1048838 vaginal birth) **Characteristics** \| Planned caesarean birth \| Vaginal birth; Maternal age, years, n (%) <20 20-29 30-39 40 \| 511 (0.9) 2222 (37.3) 33468 (56.2) 3315 (5.6) \| 23693 (2.3) 578341 (55.1) 426603 (40.7) 20201 (1.9); Infant age at follow-up, years, mean (SD) \| 16.6 (4.2) \| 17.7 (4.1); Breech presentation, n (%) \| 13623 (22.9) \| 7298 (0.7); Male children, n (%) \| 30138 (50.6) \| 533140 (50.8); Gestational age, week, n (%) 37 38 39 40 >40 \| 3439 (5.8) 8019 (11.5) 32894 (55.2) 11496 (19.3) 3666 (6.2) \| 333517 (32) 46371 (4.4) 120847 (1.5) 262507 (25.0) 285596 (27.1) **Inclusion criteria** - Singleton live births registered in Sweden in 1990-2003 **Exclusion criteria** - Preterm (<37 weeks) birth - Congenital malformation - Multiple gestations - Immigration - Mother’s ID missing - Child’s sex missing - Mode of birth missing | **Interventions**  Planned caesarean versus vaginal birth | **Details**  Swedish register-based cohort study of  term-birth singletons born between January 1, 1990, and December 31, 2003, and followed to December 31, 2013.  .  Results were reported as HR. Model 1 adjusted for child sex and birth year; model 2 further adjusted for parental characteristics, parental history of psychiatric disorders and gestational age; model 3 additionally adjusted for specific indications for planned or intrapartum CD | **Included in the decision aid (main paper)**  ***Maternal***  None  ***Infant/neonatal***  Autism Spectrum condition  **Supplementary outcomes**  ***Maternal***  None  ***Infant/neonatal***  Tic disorder  Obsessive compulsive disorder  Eating disorder  Any psychiatric disorder  Anxiety and stress-related disorder  Depression and other mood disorder  Bipolar disorder  Schizophrenia and other psychotic disorder  Childhood ADHD  Intellectual disability  Communication disorders  Learning disorders | Assessed using the Newcastle Ottawa quality assessment form for cohort studies  **Overall quality:** good |
| **Full citation**  Zweygberg AL, Martin FZ, Brynedal B, Lindholm ES, Kosidou K, Ahlqvist VH, Magnusson C. Mode of birth and subsequent self-perceived sexual life satisfaction: a population-based cohort study. American Journal of Obstetrics and Gynecology. 2024 Jul 1;231(1):107-e1. <https://doi.org/10.1016/j.ajog.2024.02.015>  **Ref Id**  Zweygberg 2024  **Country/ies where the study was carried out**  Sweden  **Study type**  Population-based cohort study  **Aim of the study**  To investigate if there was any association between mode of birth and subsequent sexual life satisfaction of  the birthing parent.  **Study dates**  The study sample included people who were recruited to the SPHC in 2006, 2010, and 2014.  **Source of funding**  Region Stockholm. Award number: RS2020-0731. | **Sample Size** N= 22,3688 participants and 42,684 births (n=4250 caesarean participants, n= 18,580 vaginal participants, n= 3212 instrumental vaginal participants; n= 5894 caesarean deliveries, n= 33,397 vaginal deliveries, n= 3393 instrumental vaginal deliveries) **Characteristics** \| Caesarean participants \| Vaginal participants; Paternal age at birth, years, mean (SD) \| 31.6 (5.2) \| 29.3 (5.0); Age at SPHC, years, mean (SD) \| 46.8 (10.3) \| 48.4 (10.4); Years since birth at time of SPHC, mean (SD) \| 15.2 (10.5) \| 18.6 (10.7); Infant birthweight class, n (%) Normal Small for gestational age Large for gestational age \| 5239 (88.9) 364 (6.2) 291 (4.9) \| 31,773 (95.1) 782 (2.3) 842 (2.5) **Inclusion criteria** Not reported **Exclusion criteria** Not reported | **Interventions**  Caesarean versus vaginal birth. No distinction is made between planned and unplanned caesarean birth. It is not specified whether the vaginal births were planned. | **Details**  This study was based on the Stockholm Public Health Cohort (SPHC). Self-reported information was supplemented with national health data and data from administrative registers, including the Swedish Medical Birth Register (MBR) and the Longitudinal Integrated Database for Health Insurance and Labor Market Studies (LISA), using the Swedish personal identity numbers for record linkage.  Results were reported as OR adjusted for birth year, parental age at time of birth, non-Swedish-born participants, parity, income, living with partner, education level, gestational age, other than normal labour presentation, birthweight class, and multiple pregnancy | **Included in the decision aid (main paper)**  ***Maternal***  Satisfaction with sex life  ***Infant/neonatal***  None  **Supplementary outcomes**  None | Assessed using the Newcastle Ottawa quality assessment form for cohort studies  **Overall quality:** good |

ADHD, attention deficit hyperactivity disorder; ASD, autism spectrum disorder; BMI, body mass index; CB, caesarean birth; cm, centimetre; g, gram; HR, hazard ratio; IBD, inflammatory bowel disease; IQR, interquartile range; kg, kilogram; NNU, neonatal unit; OR, odds ratio; PPD, postnatal psychological distress; RR, relative risk; SD, standard deviation; VB, vaginal birth; VBAC, vaginal birth after caesarean
