## Supplementary file S3 for "Outcomes of planned caesarean birth compared with planned or actual vaginal birth: an update and expansion of the NICE Caesarean Birth Guideline systematic review NG192"

Supplementary File 3 Study level quality assessment summary tables

Table S3 Study level Newcastle Ottawa Scale (NOS) assessments

| **Study ID** | **Newcastle Ottawa Scale for cohort studies** | | | | | | | | |
| --- | --- | --- | --- | --- | --- | --- | --- | --- | --- |
|  | ***Representativeness of the exposed cohort*** | ***Selection of the non-exposed cohort*** | ***Ascertainment of exposure*** | ***Demonstration that outcome of interest was not present at start of study*** | ***Compare ability of cohorts on the basis of the design or analysis*** | ***Assessment of outcome*** | ***Was follow up long enough for outcomes to occur*** | ***Adequacy of follow up of cohorts*** | ***Overall quality*** |
| Andersen 2020 | Truly representative | Drawn from the same community as the exposed cohort | Secure record | Yes | study controls for other factors (decade of birth, child’s sex, parental age at birth, gender of child and mother’s and father’s diabetes mellitus, rheumatoid arthritis, coeliac disease, and inflammatory bowel disease) | Record linkage | Yes | No statement | Good |
| Baud 2020 | Truly representative | Drawn from the same community as the exposed cohort | Secure record | Yes | study controls for other factors (age, ethnicity, parity, weight, smoking, level of education, marital status, religion, type of health insurance) | Self-report | Yes | Subjects lost to follow up unlikely to introduce bias - small number lost (the response rate was 49%) | Good |
| Chavarro 2020 | Truly representative | Drawn from the same community as the exposed cohort | Self-report | Yes | study controls for other factors (maternal age at delivery, race/ethnicity, maternal educational level, maternal pre-pregnancy BMI group, gestational weight gain, maternal height, gestational diabetes, preeclampsia, pregnancy-induced hypertension, year of birth, gestational age at delivery, birth weight group, smoking during pregnancy, and region of residence at birth) | Self-report | Yes | Complete follow up – all subjects accounted for | Good |
| Chen 2022 | Truly representative | Drawn from the same community as the exposed cohort | Secure record | Yes | study controls for other factors (caesarean delivery, gestational age, maternal age at birth, maternal education level, family income at 8 years of age and maternal diseases in pregnancy) | Record linkage | Yes | Complete follow up – all subjects accounted for | Good |
| Christiansen 2023 | Truly representative | Drawn from the same community as the exposed cohort | Self-report | Yes | Study controls for ancestry, mothers age at delivery, parity, smoking cigarettes during pregnancy, hypertensive disorder during pregnancy, diabetes during pregnancy, mode of birth, breastfeeding, allergies in childhood, use of antibiotics in childhood, fastfood habits, consumption of sugar containing drinks, consumption of sugar-free drinks, maternal SES, paternal SES, maternal change in BMI since pregnancy, paternal overweight/obese, and marital status | Self-report | Yes | Participants were retrospectively identified. 20% of those invited to participate returned questionnaires. N=891 (38.1%) were subsequently excluded from the analysis. The number excluded due to missing data is not reported. | Good |
| Currell 2022 | Somewhat representative | Drawn from the same community as the exposed cohort | Skin prick testing | Yes | study controls for maternal country of birth, maternal age at birth, maternal smoking during pregnancy, prematurity (≤36 weeks of gestational age), Socio-Economic Indexes for Areas (SEIFA) and weight for gestational age | Record linkage | Yes | Good, most subjects accounted for – very little missing data | Good |
| Dachew 2023 | Somewhat representative | Drawn from the same community as the exposed cohort | Self-report | Yes | study controls for other factors (maternal age, educational status, ethnicity, parity, pre-pregnancy body mass index, pregnancy diabetes, infection during pregnancy, hypertensive disorders during pregnancy, alcohol consumption during pregnancy, smoking during pregnancy, maternal antenatal depression and anxiety and offspring sex and gestational age at delivery) | Self-report | Yes | 69.29% of participants lost from baseline till 16 years. The final analyses were conducted on children who had complete data on exposure, outcome and covariates | Good |
| Dahlquist 2022 | Truly representative | Drawn from the same community as the exposed cohort | Secure record | Yes | study controls for other factors (maternal age, parity, body mass index, smoking, country of birth, and county) | Record linkage | Yes | Complete follow up – all subjects accounted for | Good |
| Einum 2024 | Truly representative | Drawn from the same community as the exposed cohort | Secure record | Yes | Study controls for maternal country of birth, year of first delivery, maternal age at first delivery,  smoking at the beginning of first pregnancy, chronic maternal disease (diabetes mellitus type 1 or  2, chronic hypertension, or kidney disease in the first pregnancy), gestational diabetes mellitus  in first pregnancy, and placental disease in first pregnancy (preeclampsia, eclampsia, HELLP-syndrome,  placental abruption, or fetal growth restriction (<fifth percentile of birthweight by  gestational age)) | Record linkage | Yes | Participants were retrospectively identified. No evidence that there is substantial missing data | Good |
| Guo 2021 | Truly representative | Drawn from the same community as the exposed cohort | Secure record | Yes | study controls for other factors (gestational age, maternal age, pre-pregnancy BMI, neighbourhood education quintile, maternal race, parity, conception type, anxiety, maternal substance use during pregnancy) | Record linkage | Yes | No statement | Good |
| Hagen 2024 | Truly representative | Drawn from the same community as the exposed cohort | Secure record | Yes | Study controls for age at first birth, number of births and current BMI, ethnic group | Self-report | Yes | Adequate follow-up | Good |
| Hartley 2020 | Truly representative | Drawn from the same community as the exposed cohort | Secure record | Yes | study controls for other factors (maternal pre-pregnancy weight [not overweight or obese, overweight, obese], birth weight for gestational age [small, appropriate, large], maternal smoking during pregnancy, area-level income quintile, area of residence [rural vs. urban], sex, parity [0, 1, 2, ≥3 excluding current pregnancy], maternal age at delivery) | Record linkage | Yes | Complete follow up – all subjects accounted for | Good |
| Hjorth 2019 | Somewhat representative | Drawn from the same community as the exposed cohort | Questionnaire | No | Study controls for maternal age at first birth, calendar year at first birth, pre-pregnant BMI, socio-occupational status, self-assessed health, disease, exercise in pregnancy and smoking in pregnancy | Self-report | Yes | 35710/37417 (4.56%) answered question one or more sexual problems | Good |
| Legro 2020 | Somewhat representative | Drawn from the same community as the exposed cohort | Structured interview | Yes | study controls for other factors (pre-pregnancy body mass index, gestational weight gain, age, education, poverty status, smoking, race/ethnicity, gestational age, pregnancy complications, breastfeeding and exercise habits during pregnancy and in the first year after delivery) | Self-report | Yes | Subjects lost to follow up unlikely to introduce bias – small number lost (complete data from 83.1%) | Good |
| Maher 2022 | Truly representative | Drawn from the same community as the exposed cohort | Secure record | Yes | study controls for other factors (maternal age, maternal education, maternal smoking status, maternal alcohol consumption during pregnancy, pre-pregnancy body mass index [BMI], household income, small for gestational age [SGA], infant sex, parity, hypertensive disorders of pregnancy [including raised blood pressure, eclampsia/ preeclampsia or toxaemia] and maternal depression/serious anxiety) | Record linkage | Yes | subjects lost to follow up unlikely to introduce bias – 51.45% lost from baseline to 17 years. | Good |
| Matsumoto 2025 | Truly representative | Drawn from the same community as the exposed cohort | Secure record | Yes | study controls for maternal age at delivery, multiple births, multiparity, fetal presentation, presence of fetal anomalies, maternal transport, pre-existing maternal medical conditions, pregnancy complications, maternal smoking during pregnancy, maternal alcohol consumption during pregnancy, maternal education attainment, paternal age at delivery, paternal education attainment, and place of residence at birth. In the obesity outcome model, maternal pre-pregnancy BMI (continuous) was also added as an adjustment variable. | Record linkage | Yes | 26/2114 (1.2%) infants were excluded due to missing mode of birth data | Good |
| Miller 2020 | Truly representative | Drawn from the same community as the exposed cohort | Secure record | Yes | study controls for other factors (Smoking during pregnancy, maternal age at birth, parity, gestational age, birth weight, sex, birth year, season of birth, socioeconomic status and recorded hypertensive disorders or diabetes mellitus) | Record linkage | Yes | Complete follow up – all subjects accounted for | Good |
| Mitselou 2020 | Truly representative | Drawn from the same community as the exposed cohort | Secure record | Yes | study controls for other factors (infant sex and maternal factors including age at delivery, country of birth, parity, early-pregnancy smoking, BMI, asthma/pulmonary disease) | Record linkage | Yes | No statement | Good |
| Mueller 2019 | Truly representative | Drawn from the same community as the exposed cohort | Secure record | Yes | study controls for other factors (maternal age, race, marital status, education level, household income, smoking status, maternal pre-pregnancy body mass index, and infant birth weight) | Record linkage | Yes | subjects lost to follow up unlikely to introduce bias (n=367 [65.2%] completed all four visits) | Good |
| Salem 2022 | Somewhat representative | Drawn from the same community as the exposed cohort | Structured interview | Yes | Study controls for other factors (sex, gestational age, birthweight, maternal age at birth, duration of exclusive breastfeeding, maternal smoking during pregnancy, older siblings, attendance of childcare, atopy status of the mother, parental smoking, age, and body length at study visit) | Self-report | Yes | subjects lost to follow up unlikely to introduce bias - small number lost - >14% follow up. The study reports There was no clinically relevant difference between the children included in the follow-ups and those who dropped out | Good |
| Sima 2024 | Truly representative | Drawn from the same community as the exposed cohort | Secure record | Yes |  | Record linkage and self-report | Yes | 71% of the cohort had complete time to pregnancy data (mode of birth is not reported for those with incomplete data). There does not appear to be large amounts of missing data from those with complete time to pregnancy data. | Good |
| Vaajala 2024 | Truly representative | Drawn from the same community as the exposed cohort | Secure record | Yes | Study controls for categorized maternal age and body mass index, year of pregnancy, gestational diabetes, socioeconomic status, smoking status, prior miscarriages, and prior induced abortions | Record linkage | Yes | Retrospective analysis. Doesn’t appear to be missing data from those participant that met the study inclusion criteria. | Good |
| Yoshida 2023 | Truly representative | Drawn from the same community as the exposed cohort | Secure record | Yes | Study controls for maternal age, pre-pregnancy body mass index, parity, history of depression, anxiety disorder, dysautonomia, or schizophrenia, history of physical disease, pregnancy complication, marital status, employed, highest education level, annual household income, alcohol intake, smoking status, negative attitude toward pregnancy, any major congenital anomaly, gestational age, and birth weight | Record linkage and self-report | Yes | Retrospective cohort. Appears to be complete data for the selected participant. | Good |
| Zhang 2021 | Truly representative | Drawn from the same community as the exposed cohort | Secure record | Yes | study controls for other factors (Included 3 levels of confounding adjustment: Model 1 adjusted for child sex and birth year; Model 2 further adjusted for parental characteristics, parental history of psychiatric disorders and gestational age; Model 3 additionally adjusted for specific indications for planned or intrapartum CD | Record linkage | Yes | No statement | Good |
| Zweyberg 2024 | Truly representative | Drawn from the same community as the exposed cohort | Secure record | Yes | study controls for delivery year, parental age at time of delivery, non-Swedish-born participants, parity, income, living with partner, education level, gestational age, other than normal labour presentation, birthweight class, and multiple pregnancy | Record linkage and self-report | Yes | 7.4% missing data from the full cohort (4.3% missing births and 3.1% missing other covariates. These participants were excluded from the analysis. | Good |

Table S4 Review level ROBIS assessments

| **Study ID** | **Identifying concerns in the review process** | | | | **Risk of bias in the review** | | | **Risk of bias in the review** |
| --- | --- | --- | --- | --- | --- | --- | --- | --- |
|  | ***Domain 1: concerns regarding specification of study eligibility criteria*** | ***Domain 2: concerns regarding methods used to identify and/or select studies*** | ***Domain 3: concerns regarding methods used to collect data and appraise studies*** | ***Domain 4: concerns regarding the synthesis and findings*** | ***Did the interpretation of findings address all of the concerns identified in Domains 1 to 4?*** | ***Was the relevance of identified studies to the review's research questions appropriately considered?*** | ***Did the reviewers avoid emphasizing results on the basis of their statistical significance?*** |  |
| Bodunde 2024 | Low | Low | Low | Low | Yes | Yes | Yes | Low |
| Cattani 2021 | Low | Low | Low | Low | Yes | Yes | Yes | Low |
| Chen 2024 | Low | Low | Low | Low | Yes | Yes | Yes | Low |
| Frijmersum 2025 | Low | Low | Low | Low | Yes | Yes | Yes | Low |
| Ning 2024 | Low | Low | Low | Low | Yes | Yes | Yes | Low |
| Tanoey 2019 | Low | Low | Low | Low | Yes | Yes | Yes | Low |
| Zhang 2019 | Low | Low | Low | Low | Yes | Yes | Yes | Low |
