## Supplementary file S4 for "Outcomes of planned caesarean birth compared with planned or actual vaginal birth: an update and expansion of the NICE Caesarean Birth Guideline systematic review NG192"

**Supplementary file 2**

**Table S5. Short-term clinical outcomes of planned caesarean versus planned vaginal birth which did not meet inclusion criteria for review**

| **Study ID** | **n/N (%) participants with event** | | **Effect** |
| --- | --- | --- | --- |
| **NEW MATERNAL SHORT-TERM OUTCOMES IDENTIFIED** | | | |
|  | **Planned caesarean birth** | **Planned vaginal birth** | **Measure of effect (95%CI)** |
| **Infant antibiotic use** | | | |
| Dahlquist 2022 | 0-14 days: 1758/22,855 (7.7)  0-42 days: 2937/22,855 (12.9) | 0-14 days: 3698/691,471 (0.5)  0-42 days: 62,631/691,471 (9.1) | Adjusted RR  0-14 days: 1.3 (1.3, 1.4)  0-42 days: 1.4 (1.4, 1.4) |
| Yang 2022 | Multiparous and no previous CB: 455/2588 (17.6)  Multiparous and previous CB: 3080/16571 (18.6) | Multiparous and no previous CB: 18009/72417 (24.9)  Multiparous and previous CB: 2851/9125 (31.2) | Multiparous and no previous CB: OR 0.78 (0.60, 1.01):  Multiparous and previous CB: OR 0.33 (0.27, 0.40) |
| **Unanticipated operative procedures** | | | |
| Guo 2021 | 21/1827 (1.2) | 2212/420,383 (0.5) | Adjusted RR 2.00 (1.30, 3.08) |
| **Risk of any Adverse Outcome Index outcome** | | | |
| Guo 2021 | 70/1827 (3.8) | 34,999/420,383 (8.3) | RR 0.42 (0.33, 0.54) |
| **Any maternal component of the Adverse Outcome Index** | | | |
| Guo 2021 | 37/1827 (2.0) | 18,336/420,383 (4.4) | RR 0.41 (0.30, 0.57) |
| **NEW INFANT SHORT-TERM OUTCOMES IDENTIFIED** | | | |
| **Apgar score** | | | |
| Guo 2021  Score <7 at 5min | Suppressed for small cell sizes (n<6) | 5353/420,383 (1.3) | Suppressed for small cell sizes (n<6) |
| Yang 2022  0-3 scores at 5min | Multiparous and no previous CB: 17/2918 (0.6)  Multiparous and previous CB: 36/18380 (0.2) | Multiparous and no previous CB:699/76963 (0.9)  Multiparous and previous CB: 175/9614 (1.8) | Multiparous and no previous CB: OR 0.76 (0.20, 2.94)  Multiparous and previous CB: OR 0.14 (0.04, 0.44) |
| Yang 2022  4-6 scores at 5min | Multiparous and no previous CB: 49/2918 (1.7)  Multiparous and previous CB: 100/18380 (0.5) | Multiparous and no previous CB: 769/76963 (1.0)  Multiparous and previous CB: 189/9614 (2.0) | Multiparous and no previous CB: OR 0.47 (0.24, 0.93)  Multiparous and previous CB: OR 0.30 (0.17, 0.54) |
| **Any neonatal component of Adverse Outcome Index (AOI)** | | | |
| Guo 2021 | 34/1827 (1.9) | 17,899/420,383 (4.3) | RR 0.42 (0.30, 0.58) |

**CB=caesarean birth; RR=relative risk; OR=odds ratio**

**Table S6. Long-term clinical outcomes of caesarean versus vaginal birth which did not meet inclusion criteria for review**

| **Study ID** | **n/N (%) participants with event** | | **Effect** |
| --- | --- | --- | --- |
|  | **Caesarean birth (P=planned, M= mixed planned and unplanned)** | **Vaginal birth** | **Relative (95%CI)** |
| **LONG-TERM MATERNAL OUTCOMES** | | | |
| **Urgency urinary incontinence** | | | |
| Baud 2020 | 46/208 (22.1) (P) | 102/309 (33.0) | RR 0.55 (0.34, 0.88) |
| **Stress incontinence** | | | |
| Baud 2020 | 75/208 (36.1)  (P) | 156/309 (50.5) | RR 0.53 (0.35, 0.80) |
| **Lower abdominal or genital pain** | | | |
| Baud 2020 | 62/208 (29.8) (P) | 65/309 (21.0) | RR 1.58 (1.01, 2.49) |
| **NEW MATERNAL LONG-TERM OUTCOMES IDENTIFIED** | | | |
| **Postpartum weight retention at 1 year postpartum (retained 10 pounds or more)** | | | |
| Legro 2020 | 203 (27.9) (M) | 394 (22.2) | OR 1.30 (1.04, 1.62) |
| **Depression** | | | |
| Bodunde 2024 | NR (M) | NR | OR 1.32 (0.93, 1.87 ) |
| **Anxiety** | | | |
| Bodunde 2024 | NR (M) | NR | OR 1.14 (0.95, 1.37 ) |
| **Anal incontinence** | | | |
| Cattani 2021 (MA of 8 studies) | NR (M) | NR | OR 1.22 (1.08, 1.38) |
| **Flatus incontinence** | | | |
| Cattani 2021 (MA of 4 studies) | NR (M) | NR | OR 1.20 (1.05, 1.37) |
| **Wexner faecal incontinence subscale scores** | | | |
| ***Wexner incontinence for gas score*** | | | |
| Baud 2020 | NR/208 (38.6) (P) | NR/309 (39.6) | RR 0.95 (0.63, 1.42) |
| ***Wexner incontinence for liquid stool score*** | | | |
| Baud 2020 | NR/208 (11.4) (P) | NR/309 (10.0) | RR 1.10 (0.57, 2.08) |
| ***Wexner incontinence for solid stool score*** | | | |
| Baud 2020 | NR/208 (1.9) (P) | NR/309 (2.9) | RR 0.58 (0.12, 2.70) |
| ***Wexner alteration of lifestyle score*** | | | |
| Baud 2020 | NR/208 (7.1) (P) | NR/309 (5.5) | RR 1.04 (0.43, 2.54) |
| ***Wexner alteration of sexual life score*** | | | |
| Baud 2020 | NR/208 (4.6) (P) | NR/309 (1.4) | RR 1.72 (1.13, 2.63) |
| ***Wexner need to wear a pad score*** | | | |
| Baud 2020 | NR/208 (6.8) (P) | NR/309 (6.2) | RR 0.97 (0.43, 2.15) |
| ***Wexner taking constipating medicine score*** | | | |
| Baud 2020 | NR/208 (0.5) (P) | NR/309 (0.3) | RR 3.23 (0.16, 66.1) |
| ***Wexner inability to defer defecation for 15 minutes score*** | | | |
| Baud 2020 | NR/208 (13.5) (P) | NR/309 (9.8) | RR 1.66 (0.88, 3.12) |
| **Lower back pain at 12 months** | | | |
| Frijmersum 2025 (Woolhouse 2012) | NR (M) | NR | OR 1.22 (0.92, 1.61) |
| **LONG-TERM INFANT OUTCOMES** | | | |
| **NEW INFANT LONG-TERM OUTCOMES IDENTIFIED** | | | |
| **Infant hospital-related infection by age of first occurrence: 0-3 months** | | | |
| Miller 2020 | 23,760/590,769 (4.0)^a^ (P) | NR | I-V HR 1.16 (1.15, 1.18)  D+L HR 1.16 (1.14, 1.19) |
| **Infant hospital-related infection by age of first occurrence: 4-6 months** | | | |
| Miller 2020 | 13,671/563,043 (2.4)^a^ (P) | NR | I-V HR 1.21 (1.18, 1.23)  D+L HR 1.19 (1.15, 1.24) |
| **Infant hospital-related infection by age of first occurrence: 7-12 months** | | | |
| Miller 2020 | 24,894/552,431 (4.5)^a^ (P) | NR | I-V HR 1.15 (1.13, 1.16)  D+L HR 1.14 (1.11, 1.17) |
| **Infant hospital-related infection by age of first occurrence: 1-2 years** | | | |
| Miller 2020 | 36,488/465,105 (7.8)^a^ (P) | NR | I-V HR 1.14 (1.13, 1.15)  D+L HR 1.14 (1.11, 1.16) |
| **At least one infection-related hospitalisation until 5 years old** | | | |
| Miller 2020 | NR (P) | NR | HR 1.13 (1.12, 1.13) |
| **Infant hospital-related infection by age of first occurrence: 2-5 years** | | | |
| Miller 2020 | 38,800/467,571 (8.3)^a^ (P) | NR | I-V HR 1.12 (1.11, 1.13)  D+L HR 1.11 (1.08, 1.15) |
| **Infection-related hospitalisation: invasive bacterial** | | | |
| Miller 2020 | 2,408/456,489 (0.5)^a^ (P) | NR | I-V HR 1.05 (1.00, 1.09)  D+L HR 1.00 (0.91, 1.09) |
| **Infection-related hospitalisation: skin and soft tissue** | | | |
| Miller 2020 | 6,307/460,532 (1.4)^a^ (P) | NR | I-V HR 1.04 (1.01, 1.07)  D+L HR 1.02 (0.97, 1.08) |
| **Infection-related hospitalisation: genitourinary** | | | |
| Miller 2020 | 6,889/461,590 (1.5)^a^ (P) | NR | I-V HR 1.14 (1.11, 1.17)  D+L HR 1.13 (1.09, 1.17) |
| **Infection-related hospitalisation: lower respiratory tract** | | | |
| Miller 2020 | 48,699/503,198 (9.7)^a^ (P) | NR | I-V HR 1.16 (1.15, 1.17)  D+L HR 1.15 (1.12, 1.18) |
| **Infection-related hospitalisation: upper respiratory tract** | | | |
| Miller 2020 | 58,914/514,253 (11.5) ^a^ (P) | NR | I-V HR 1.19 (1.18, 1.20)  D+L HR 1.18 (1.16, 1.20) |
| **Infection-related hospitalisation: viral** | | | |
| Miller 2020 | 34,721/489,545 (7.1) ^a^ (P) | NR | I-V HR 1.17 (1.16, 1.19)  D+L HR 1.17 (1.13, 1.20) |
| **Infection-related hospitalisation: gastrointestinal** | | | |
| Miller 2020 | 22,429/477,287 (4.7) ^a^ (P) | NR | I-V HR 1.21 (1.19, 1.23)  D+L HR 1.18 (1.13, 1.23) |
| **Allergic rhinitis** | | | |
| Mitselou 2020 | 1907/86,576 (2.2) (P) | NR | HR 1.10 (1.05, 1.15) |
| **Allergic rhinoconjunctivitis** | | | |
| Salem 2022 | 0/24 (0.0) (P) | 15/306 (4.9) | OR 0.6 (0.0, 11.9) |
| **Weeks with any respiratory symptom** | | | |
| Salem 2022 | Number of weeks, median (IQR)  3 (6) (P) | Number of weeks, median (IQR)  5 (7) | IRR 0.8 (0.6, 1.1) |
| **Ever wheezing at early-school age** | | | |
| Salem 2022 | 5/24 (20.8) (P) | 64/306 (20.9) | OR 1.1 (0.4, 3.4) |
| **Atopic dermatitis** | | | |
| Salem 2022 | 1/24 (4.2) (P) | 27/306 (8.8) | OR 0.3 (0.0, 2.4) |
| **Coeliac disease** | | | |
| Andersen 2020 | 248/122,419 (P) | 2677/1,312,522 | HR 1.04 (0.92, 1.19) |
| **Risk of otitis media in childhood** | | | |
| Hartley 2020 | NR (M) | NR (ref) | HR 1.06 (1.03, 1.09) |
| **Number of otitis media episodes** | | | |
| Hartley 2020 | NR (M) | NR (ref) | IRR 1.04 (1.01, 1.07) |
| **Otitis media episodes >95^th^ percentile** | | | |
| Hartley | NR (M) | NR (ref) | OR 1.10 (0.00, 1.23) |
| **Infant weight gain and adiposity at 12 months** | | | |
| ***Weight-for-length at 12 months*** | | | |
| Mueller 2019 | NR (M) | NR | Adjusted mean difference between caesarean and vaginal births:  z score (95%CI) 0.26 (0.05, 0.47) |
| ***Abdominal skinfold thickness at 12 months (mm)*** | | | |
| Mueller 2019 | NR (M) | NR | Adjusted mean difference between caesarean and vaginal births (95% CI): 0.27 (-0.17, 0.71) |
| ***Subscapular skinfold thickness at 12 months (mm*)** | | | |
| Mueller 2019 | NR (M) | NR | Adjusted mean difference between caesarean and vaginal births (95% CI): 0.42 (0.12, 0.73) |
| ***Triceps skinfold thickness at 12 months (mm)*** | | | |
| Mueller 2019 | NR (M) | NR | Adjusted mean difference between caesarean and vaginal births (95% CI): 0.52 (0.09, 0.95) |
| ***Subscapular and triceps skinfold thickness at 12 months*** | | | |
| Mueller 2019 | NR (M) | NR | Adjusted mean difference between caesarean and vaginal births: mm (95%CI) 0.95 (0.30, 1.60) |
| **Any food allergy** | | | |
| Currell 2022 | 37/277 (13.4) (M) | 333/2989 (11.1) | OR 1.05 (0.71, 1.55) |
| **Egg allergy** | | | |
| Currell 2022 | 30/279 (10.8) (M) | 301/3035 (9.9) | OR 0.92 (0.60, 1.41) |
| **Peanut allergy** | | | |
| Currell 2022 | 12/281 (4.3) (M) | 92/2972 (3.1) | OR 1.10 (0.56, 2.14) |
| **Inflammatory bowel disease** | | | |
| Andersen 2020 | 3735/122,419 (3.1) (P) | 316/1,312,522 (0.1) | HR 1.16 (1.03, 1.30) |
| **Rheumatoid arthritis** | | | |
| Andersen 2020 | 353/122,419 (P) | 3776/1,312,522 | HR 1.14 (1.02, 1.27) |
| **Adulthood DM2** | | | |
| Chavarro 2020 | 87 (61,876 person-years) (M) | 1927 (1,852,102 person-years) (ref) | HR 1.46 (1.18, 1.81) |
| **Adulthood obesity** | | | |
| Chavarro 2020 | 434/1089 (39.9) (M) | 11,722/32,137 (36.5) (ref) | RR 1.11 (1.03, 1.19) |
| **Tic disorder** | | | |
| Zhang 2019 (MA of 2 studies) | 939/336,675 (0.3) (P) | 5242/2,614,522 (0.2) | OR 1.23 (0.73, 2.07) |
| **Obsessive-compulsive disorder** | | | |
| Zhang 2021 | 299/59,514 (0.5) (P) | 5035/1,048,838 (0.5) | HR 1.08 (0.92, 1.28) |
| **Eating disorder** | | | |
| Zhang 2021 | 435/59,514 (0.7) (P) | 8745/1,048,838 (0.8) | 0.89 (0.78, 1.01) |
| **Affective and non-affective psychoses** | | | |
| Zhang 2019 (MA of 4 studies) | NR (P) | NR | OR 1.21 (0.86, 1.71) |
| **Any psychiatric disorder** | | | |
| Zhang 2021 | NR (P) | NR | HR 1.04 (0.99, 1.10) |
| **Anxiety and stress-related disorder** | | | |
| Zhang 2021 | 2034/59,514 (3.4) (P) | 37,593/1,048,838 (3.6) | HR 1.07 (1.00, 1.14) |
| **Depression and other mood disorder** | | | |
| Zhang 2021 | 1465/59,514 (2.5) (P) | 29,205/1,048,838 (2.8) | HR 1.03 (0.96, 1.10) |
| **Bipolar disorder** | | | |
| Zhang 2021 | 158/59,514 (0.3) (P) | 3084/1,048,838 (0.3) | HR 1.03 (0.83, 1.27) |
| **Schizophrenia and other psychotic disorder** | | | |
| Zhang 2021 | 100/59,514 (0.2) (P) | 1677/1,048,838 (0.2) | HR 1.15 (0.88, 1.51) |
| **Total behavioural difficulties at 3 years** | | | |
| Dachew 2023 | NR (P) | NR | OR 1.04 (0.84, 1.28) |
| **Total behavioural difficulties at 7 years** | | | |
| Dachew 2023 | NR (P) | NR | OR 0.98 (0.62, 1.55) |
| **Total behavioural difficulties at 9 years** | | | |
| Dachew 2023 | NR (P) | NR | OR 0.90 (0.54, 1.51) |
| **Total behavioural difficulties at 11 years** | | | |
| Dachew 2023 | NR (P) | NR | OR 0.80 (0.45, 1.40) |
| **Total behavioural difficulties at 16 years** | | | |
| Dachew 2023 | NR (P) | NR | OR 0.65 (0.30, 1.44) |
| **Emotional symptoms at 3 years** | | | |
| Dachew 2023 | NR (P) | NR | OR 0.89 (0.66, 1.21) |
| **Emotional symptoms at 7 years** | | | |
| Dachew 2023 | NR (P) | NR | OR 1.26 (0.85, 1.85) |
| **Emotional symptoms at 9 years** | | | |
| Dachew 2023 | NR (P) | NR | OR 1.29 (0.86, 1.91) |
| **Emotional symptoms at 11 years** | | | |
| Dachew 2023 | NR (P) | NR | OR 1.39 (0.91, 2.10) |
| **Emotional symptoms at 16 years** | | | |
| Dachew 2023 | NR (P) | NR | OR 0.83 (0.48, 1.43) |
| **Peer relationship problems at 7 years** | | | |
| Dachew 2023 | NR (P) | NR | OR 1.43 (1.01, 2.04) |
| **Peer relationship problems at 9 years** | | | |
| Dachew 2023 | NR (P) | NR | OR 1.19 (0.82, 1.74) |
| **Peer relationship problems at 11 years** | | | |
| Dachew 2023 | NR (P) | NR | OR 0.82 (0.53, 1.28) |
| **Peer relationship problems at 16 years** | | | |
| Dachew 2023 | NR (P) | NR | OR 0.70 (0.50, 1.21) |
| **Hyperactivity/inattention problems at 3 years** | | | |
| Dachew 2023 | NR (P) | NR | OR 0.88 (0.53, 1.44) |
| **Hyperactivity/inattention problems at 7 years** | | | |
| Dachew 2023 | NR (P) | NR | OR 1.22 (0.87, 1.70) |
| **Hyperactivity/inattention problems at 9 years** | | | |
| Dachew 2023 | NR (P) | NR | OR 0.88 (0.56, 1.39) |
| **Hyperactivity/inattention problems at 11 years** | | | |
| Dachew 2023 | NR (P) | NR | OR 0.97 (0.60, 1.56) |
| **Hyperactivity/inattention problems at 16 years** | | | |
| Dachew 2023 | NR (P) | NR | OR 0.74 (0.38, 1.45) |
| **Conduct problems at 3 years** | | | |
| Dachew 2023 | NR (P) | NR | OR 0.91 (0.70, 1.18) |
| **Conduct problems at 7 years** | | | |
| Dachew 2023 | NR (P) | NR | OR 1.03 (0.75, 1.41) |
| **Conduct problems at 9 years** | | | |
| Dachew 2023 | NR (P) | NR | OR 1.02 (0.69, 1.51) |
| **Conduct problems at 11 years** | | | |
| Dachew 2023 | NR (P) | NR | OR 0.75 (0.47, 1.19) |
| **Conduct problems at 16 years** | | | |
| Dachew 2023 | NR (P) | NR | OR 1.01 (0.59, 1.74) |
| **Prosocial behaviours at 7 years** | | | |
| Dachew 2023 | NR (P) | NR | OR 1.32 (0.89, 1.97) |
| **Prosocial behaviours at 9 years** | | | |
| Dachew 2023 | NR (P) | NR | OR 1.15 (0.70, 1.92) |
| **Prosocial behaviours at 11 years** | | | |
| Dachew 2023 | NR (P) | NR | OR 1.36 (0.57, 1.44) |
| **Prosocial behaviours at 16 years** | | | |
| Dachew 2023 | NR (P) | NR | OR 0.45 (0.21, 0.98) |
| **Emotional difficulties at 3 years** | | | |
| Maher 2022 | NR (P) | NR | OR 0.99 (0.70, 1.39) |
| **Emotional difficulties at 5 years** | | | |
| Maher 2022 | NR (P) | NR | OR 0.93 (0.64, 1.35) |
| **Emotional difficulties at 7 years** | | | |
| Maher 2022 | NR (P) | NR | OR 0.99 (0.71, 1.38) |
| **Emotional difficulties at 11 years** | | | |
| Maher 2022 | NR (P) | NR | OR 1.16 (0.89, 1.52) |
| **Emotional difficulties at 14 years** | | | |
| Maher 2022 | NR (P) | NR | OR 0.92 (0.69, 1.21) |
| **Emotional difficulties at 17 years** | | | |
| Maher 2022 | NR (P) | NR | OR 1.27 (0.97, 1.66) |
| **Conduct difficulties at 3 years** | | | |
| Maher 2022 | NR (P) | NR | OR 0.94 (0.79, 1.11) |
| **Conduct difficulties at 5 years** | | | |
| Maher 2022 | NR (P) | NR | OR 0.63 (0.46, 0.85) |
| **Conduct difficulties at 7 years** | | | |
| Maher 2022 | NR (P) | NR | OR 1.02 (0.77, 1.35) |
| **Conduct difficulties at 11 years** | | | |
| Maher 2022 | NR (P) | NR | OR 0.86 (0.63, 1.18) |
| **Conduct difficulties at 14 years** | | | |
| Maher 2022 | NR (P) | NR | OR 1.09 (0.80, 1.48) |
| **Conduct difficulties at 17 years** |  |  |  |
| Maher 2022 | NR (P) | NR | OR 1.22 (0.86, 1.73) |
| **Hyperactivity at 3 years** | | | |
| Maher 2022 | NR (P) | NR | OR 1.01 (0.81) |
| **Hyperactivity at 5 years** | | | |
| Maher 2022 | NR (P) | NR | OR 0.91 (0.69, 1.21) |
| **Hyperactivity at 7 years** | | | |
| Maher 2022 | NR (P) | NR | OR 0.94 (0.72, 1.21) |
| **Hyperactivity at 11 years** | | | |
| Maher 2022 | NR (P) | NR | OR 0.93 (0.69, 1.25) |
| **Hyperactivity at 14 years** | | | |
| Maher 2022 | NR (P) | NR | OR 0.93 (0.67, 1.29) |
| **Hyperactivity at 17 years** |  |  |  |
| Maher 2022 | NR (P) | NR | OR 1.00 (0.65, 1.53) |
| **Peer problems at 3 years** | | | |
| Maher 2022 | NR (P) | NR | OR 0.85 (0.66, 1.09) |
| **Peer problems at 5 years** | | | |
| Maher 2022 | NR (P) | NR | OR 0.93 (0.68, 1.28) |
| **Peer problems at 7 years** | | | |
| Maher 2022 | NR (P) | NR | OR 0.87 (0.64, 1.18) |
| **Peer problems at 11 years** | | | |
| Maher 2022 | NR (P) | NR | OR 0.90 (0.66, 1.21) |
| **Peer problems at 14 years** | | | |
| Maher 2022 | NR (P) | NR | OR 0.85 (0.66, 1.11) |
| **Peer problems at 17 years** | | | |
| Maher 2022 | NR (P) | NR | OR 1.04 (0.80, 1.36) |
| **Prosocial behaviour at 3 years** | | | |
| Maher 2022 | NR (P) | NR | OR 0.67 (0.48, 0.94) |
| **Prosocial behaviour at 5 years** | | | |
| Maher 2022 | NR (P) | NR | OR 0.87 (0.51, 1.47) |
| **Prosocial behaviour at 7 years** | | | |
| Maher 2022 | NR (P) | NR | OR 0.56 (0.28, 1.12) |
| **Prosocial behaviour at 11 years** | | | |
| Maher 2022 | NR (P) | NR | OR 0.79 (0.36, 1.75) |
| **Prosocial behaviour at 14 years** | | | |
| Maher 2022 | NR (P) | NR | OR 0.85 (0.52, 1.36) |
| **Prosocial behaviour at 17 years** | | | |
| Maher 2022 | NR (P) | NR | OR 0.84 (0.46, 1.53) |
| **Neurodevelopmental disorders at 8 years of age** | | | |
| Chen 2022 | 341/6116 (5.6) (P) | 648/13,026 (5.0) (ref) | OR 0.98 (0.82, 1.18) |
| Zhang 2021 (ref ID 3658) | 2997/59,514 (5.0) (P) | 45,692/1,048,838 (4.4) | HR 1.17 (1.12, 1.23) |
| **Childhood ADHD OR data** | | | |
| Zhang 2019 (MA of 4 studies) | NR (P) | NR | OR 1.09 (1.05, 1.13) |
| **Intellectual Disability** | | | |
| Zhang 2021 | 693/59,514 (1.2) (P) | 8783/1,048,838 (0.8) | HR 1.26 (1.14, 1.39) |
| **Communication disorders** | | | |
| Zhang 2021 | 482/59,514 (0.8) (P) | 6234/1,048,838 (0.6) | HR 1.14 (1.02, 1.28) |
| **Learning disorders** | | | |
| Zhang 2021 | 377/59,514 (0.6) (P) | 5686/1,048,838 (0.5) | HR 1.15 (1.01, 1.30) |

a. Manually calculated using the author reported % Cases and Total N for the individual countries’ datasets (Denmark, Scotland, England, New South Wales, Western Australia)

**D+L= DerSimonian and Laird random effects model; DM2=type 2 diabetes; I-V = inverse-variance weighted fixed effects model; MA = meta-analysis; RR=relative risk; OR=odds ratio; HR=hazard ratio; NR=not reported; ADHD = attention deficit and hyperactivity disorder**
